# Deep untargeted wastewater metagenomic sequencing from sewersheds across the United States

**DOI:** 10.64898/2026.03.05.26345726

**Authors:** Lennart J. Justen, Clayton Rushford, Olivia S. Hershey, Róisín Floyd-O’Sullivan, Simon L. Grimm, William J. Bradshaw, Harmon Bhasin, Daniel P. Rice, Katherine Stansifer, Jo D. Faraguna, Michael R. McLaren, Alessandro Zulli, Alejandro Tovar-Mendez, Emma E. Copen, Kristen K. Shelton, Ayaaz Amirali, Sherin Kannoly, Sofia Pesantez, Aiden Stanciu, Iñigo Caballero Quiroga, Leopolda Silvera, Nicole Greenwood, Barbra Bongiovi, Austin Walkins, Ryan Love, Scott Lening, Kaylyn Patterson, Theresa Johnston, Sandra Hernandez, Aymara Benitez, Billie Jo McCarley, Samantha Engelage, Suguna Pillay, Cindy Calender, Brent Herring, Carey Robinson, Monett Wastewater Treatment Plant, Columbia Missouri Wastewater Treatment Plant, Daniel Cunningham-Bryant, Gordon Adams, Jillian Paull, Jamie Devlin, Vamsi Thiriveedhi, Sarah E. Turbett, Jacob E. Lemieux, Rose S. Kantor, David H. O’Connor, John J. Dennehy, Rachel Poretsky, Jason A. Rothman, Helena M. Solo-Gabriele, Jason R. Vogel, Pardis C. Sabeti, Jeff Kaufman, Marc C. Johnson

**Author notes:** These authors contributed equally to this work (Lennart J. Justen, Clayton Rushford).

## Abstract

Wastewater monitoring enables non-invasive, population-scale tracking of community infections independent of healthcare-seeking behavior and clinical diagnosis. Metagenomic sequencing extends this capability by enabling broad, pathogen-agnostic detection of hundreds of viral taxa, including emerging threats. Here, we present data from CASPER (the Coalition for Agnostic Sequencing of Pathogens from Environmental Reservoirs), a U.S.-based wastewater metagenomic sequencing network designed for deep, untargeted pathogen monitoring at national scale. This release includes 1,206 samples collected between December 2023 and December 2025 from 27 sites across nine states, covering 13 million people. Deep sequencing (∼1 billion read pairs per sample) generated 1.2 trillion read pairs (358 terabases), capturing diverse human, animal, and plant viruses including rare taxa. CASPER data tracks wastewater digital PCR and clinical abundance trends for SARS-CoV-2, influenza A, and respiratory syncytial virus, and recovers national-scale variant dynamics for SARS-CoV-2 and norovirus. As the largest publicly available untargeted wastewater sequencing dataset to date, CASPER provides a shared and growing resource for pathogen surveillance and microbial ecology.

## Main

Wastewater monitoring has emerged as a powerful tool for providing early warning of pathogen outbreaks and tracking community-level infections independent of health care-seeking behavior (Diamond et al., 2022; Hellmér et al., 2014; Karthikeyan et al., 2022; Kirby et al., 2022; Peccia et al., 2020; Rebecca Falender et al., 2026). A single wastewater sample can capture pathogens shed by millions of individuals, including those with asymptomatic or pre-symptomatic infections, providing a comparatively unbiased snapshot of pathogen circulation at scale (Committee on Community Wastewater-based Infectious Disease Surveillance et al., 2024; Corrin et al., 2024; Wolfe et al., 2021).

In practice, current wastewater monitoring programs rely on targeted detection approaches such as quantitative PCR (qPCR), digital PCR (dPCR), or amplicon sequencing to detect predefined pathogen targets (Adams, Bias, et al., 2024; Boehm et al., 2024; Duvallet et al., 2022; Morfino et al., 2023). These approaches are sensitive, cost-effective, and valuable for public health, but face limitations: they cannot detect novel pathogens outside their targets, require sometimes lengthy assay design and validation for each new target, often during critical periods of an outbreak (Adams, Kirby, et al., 2024; Wolfe et al., 2022), and may lose sensitivity as mutations occur in primer binding sites (Kidd et al., 2021; Lambisia et al., 2022; Public Health England, 2020; Schuele et al., 2024; Wu et al., 2023). In one example, a widely used measles wastewater dPCR assay failed to detect an internationally circulating genotype B3 variant owing to a mutation in the probe binding region, while untargeted sequencing from CASPER (not in this release) confirmed the virus was present in the same wastewater (Pray et al., 2026). Hybrid-capture sequencing is a promising semi-targeted alternative detection platform (Tisza et al., 2023; Wolfe et al., 2026; Zulli et al., 2025), using probe panels to enrich for sequences similar to known pathogens and tolerating greater sequence divergence (up to 20–30%) than amplicon-based methods (Kantor & Jiang, 2024).

Unlike these approaches, wastewater metagenomic sequencing (WW-MGS) is untargeted, using no pathogen-specific primers or probes, enabling pathogen-agnostic detection of known and emerging pathogens across the microbial community (Wongprommoon et al., 2024). WW-MGS, including from CASPER, has already demonstrated its utility for large-scale microbial ecology surveys (Edgar et al., 2022; Fiamenghi et al., 2025; Kantor et al., 2026; Neri et al., 2022; Worp et al., 2025) and has shown the ability to detect and track a broad range of human and animal pathogens (Justen, Zulli, et al., 2026; Rothman et al., 2021; Rushford et al., 2025; Spurbeck et al., 2023), including emerging threats such as influenza H5N1 (Rushford et al., 2025) and measles virus (Poretsky et al., 2026).

Despite this potential, routine untargeted WW-MGS has seen limited adoption as a biosecurity or public health tool. Vertebrate-infecting viruses constitute a small fraction of total nucleic acids in wastewater, requiring deep and costly sequencing for sensitive detection (Grimm, Kaufman, et al., 2025); computational infrastructure must scale to hundreds of terabases; and interpretation of detections is challenged by the diverse sources of nucleic acids in wastewater (Grassly et al., 2025). However, continued declines in sequencing costs and the expansion of wastewater monitoring infrastructure have made routine deep, untargeted WW-MGS increasingly feasible.

We established CASPER (Coalition for Agnostic Sequencing of Pathogens from Environmental Reservoirs) to enable deep, untargeted wastewater metagenomic sequencing at national scale. CASPER is a collaboration of U.S.-based partners, including academic institutions, national laboratories, municipal wastewater utilities, and hospitals, advancing pathogen monitoring through untargeted sequencing. Launched through a partnership between SecureBio (SB) and the University of Missouri (MU), the network has since expanded across the country and continues to grow, with data released periodically on the NCBI Sequence Read Archive (SRA). Data is shared with consortium members in near real time, supporting various independent analyses by partners, including public dashboards maintained by the University of Wisconsin-Madison (O’Connor et al., 2025). Detections of immediate concern are reported to relevant public health and biosecurity authorities. CASPER protocols primarily capture RNA, covering an important diversity of plant-, bacteria-, and vertebrate-infecting viruses, including likely sources of future human-infecting pandemic pathogens (Adalja et al., 2018), though some DNA and DNA viruses are also captured. To enable sensitive detection of rare taxa, we sequence to a depth of approximately one billion read pairs per sample—far deeper than most previous efforts.

Here, we report the public release of CASPER data from 27 sites approved for sharing. The release comprises 1.2 trillion read pairs (358 terabases (Tb)) from samples collected between December 2023 and December 2025, spanning nine U.S. states and covering over 13 million people. A previous, smaller CASPER release comprising 13 Tb from a Los Angeles pilot study is also publicly available (Grimm, Rothman, et al., 2025; BioProject PRJNA1198001). Together, CASPER constitutes the largest public repository of WW-MGS data to date. Beyond pathogen surveillance, this data provides a shared resource for research in microbial ecology, viral discovery, antimicrobial resistance, and methodological development. All data is publicly available (BioProject PRJNA1247874). We invite the research and public health communities to use and build upon this growing resource.

## Results

### Sampling collection, processing, and sequencing

CASPER sampling sites in this data release span nine U.S. states and encompass diverse geographic regions and major population centers (Figure 1a). This release includes 27 sites (Table 1). Using sewershed population estimates from the National Wastewater Surveillance System (NWSS) (CDC, 2025c) and other sources, we estimate that these sites collectively serve approximately 13 million people (Table 1). This figure counts each physical catchment once and excludes hospital and airport sites, which lack reliable population estimates, as well as one site that opted not to disclose.

**Table 1.**
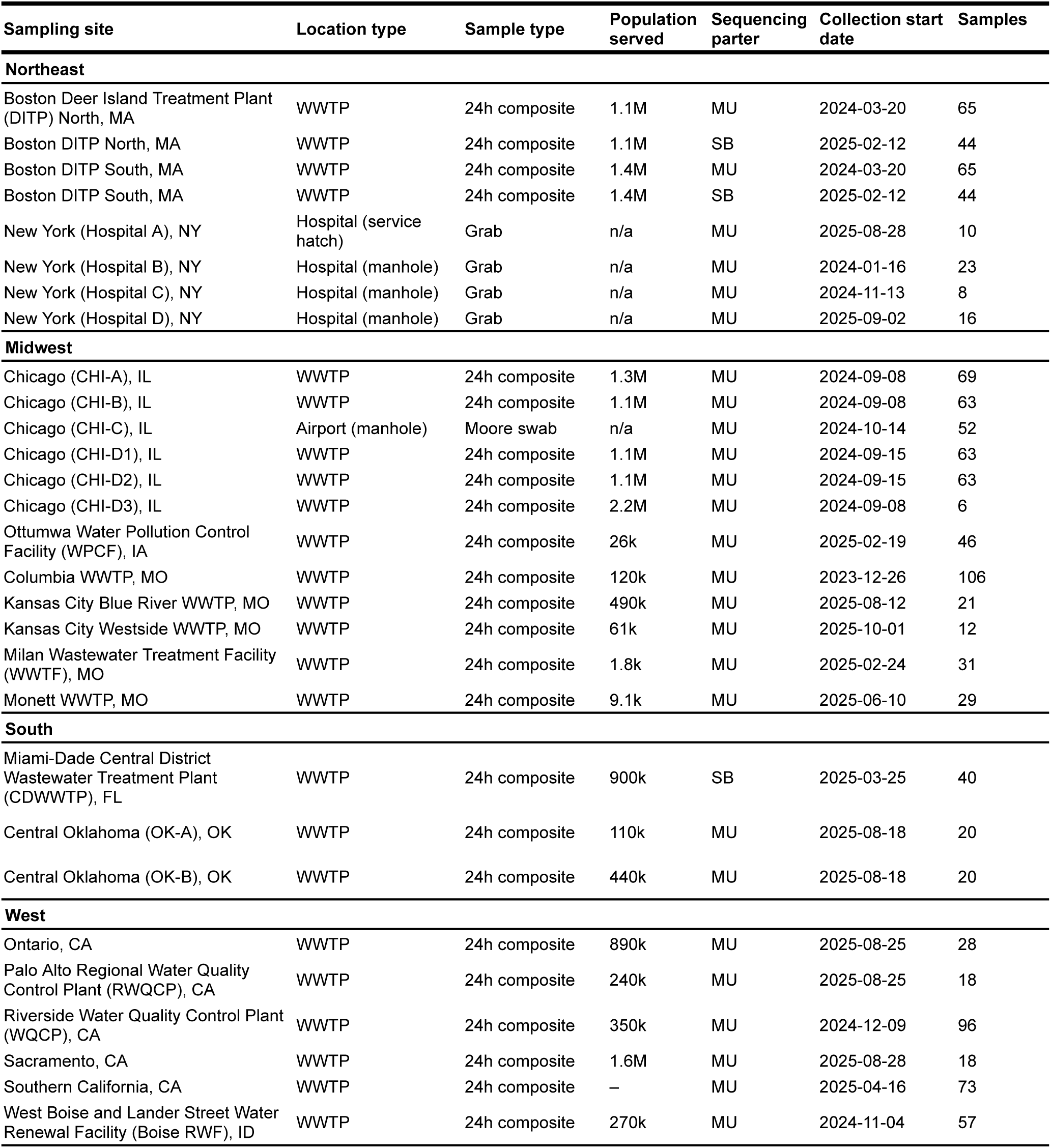
Summary of CASPER sampling sites included in this data release. The table lists site location, location type, sample type, estimated population served, sequencing partner, collection start date, and samples in this data release. Population estimates are approximate and based on sewershed coverage data from the National Wastewater Surveillance System (NWSS) where available. "n/a" indicates non-wastewater treatment plant (WWTP) sites for which contributing population estimates are difficult to determine, and "–" indicates a site that opted not to disclose its population estimate. MU, University of Missouri; SB, SecureBio.

| Sampling site | Location type | Sample type | Population served | Sequencing partner | Collection start date | Samples |
| --- | --- | --- | --- | --- | --- | --- |
| **Northeast** |  |  |  |  |  |  |
| Boston Deer Island Treatment Plant (DITP) North, MA | WWTP | 24h composite | 1.1M | MU | 2024-03-20 | 65 |
| Boston DITP North, MA | WWTP | 24h composite | 1.1M | SB | 2025-02-12 | 44 |
| Boston DITP South, MA | WWTP | 24h composite | 1.4M | MU | 2024-03-20 | 65 |
| Boston DITP South, MA | WWTP | 24h composite | 1.4M | SB | 2025-02-12 | 44 |
| New York (Hospital A), NY | Hospital (service hatch) | Grab | n/a | MU | 2025-08-28 | 10 |
| New York (Hospital B), NY | Hospital (manhole) | Grab | n/a | MU | 2024-01-16 | 23 |
| New York (Hospital C), NY | Hospital (manhole) | Grab | n/a | MU | 2024-11-13 | 8 |
| New York (Hospital D), NY | Hospital (manhole) | Grab | n/a | MU | 2025-09-02 | 16 |
| **Midwest** |  |  |  |  |  |  |
| Chicago (CHI-A), IL | WWTP | 24h composite | 1.3M | MU | 2024-09-08 | 69 |
| Chicago (CHI-B), IL | WWTP | 24h composite | 1.1M | MU | 2024-09-08 | 63 |
| Chicago (CHI-C), IL | Airport (manhole) | Moore swab | n/a | MU | 2024-10-14 | 52 |
| Chicago (CHI-D1), IL | WWTP | 24h composite | 1.1M | MU | 2024-09-15 | 63 |
| Chicago (CHI-D2), IL | WWTP | 24h composite | 1.1M | MU | 2024-09-15 | 63 |
| Chicago (CHI-D3), IL | WWTP | 24h composite | 2.2M | MU | 2024-09-08 | 6 |
| Ottumwa Water Pollution Control Facility (WPCF), IA | WWTP | 24h composite | 26k | MU | 2025-02-19 | 46 |
| Columbia WWTP, MO | WWTP | 24h composite | 120k | MU | 2023-12-26 | 106 |
| Kansas City Blue River WWTP, MO | WWTP | 24h composite | 490k | MU | 2025-08-12 | 21 |
| Kansas City Westside WWTP, MO | WWTP | 24h composite | 61k | MU | 2025-10-01 | 12 |
| Milan Wastewater Treatment Facility (WWTF), MO | WWTP | 24h composite | 1.8k | MU | 2025-02-24 | 31 |
| Monett WWTP, MO | WWTP | 24h composite | 9.1k | MU | 2025-06-10 | 29 |
| **South** |  |  |  |  |  |  |
| Miami-Dade Central District Wastewater Treatment Plant (CDWWTP), FL | WWTP | 24h composite | 900k | SB | 2025-03-25 | 40 |
| Central Oklahoma (OK-A), OK | WWTP | 24h composite | 110k | MU | 2025-08-18 | 20 |
| Central Oklahoma (OK-B), OK | WWTP | 24h composite | 440k | MU | 2025-08-18 | 20 |
| **West** |  |  |  |  |  |  |
| Ontario, CA | WWTP | 24h composite | 890k | MU | 2025-08-25 | 28 |
| Palo Alto Regional Water Quality Control Plant (RWQCP), CA | WWTP | 24h composite | 240k | MU | 2025-08-25 | 18 |
| Riverside Water Quality Control Plant (WQCP), CA | WWTP | 24h composite | 350k | MU | 2024-12-09 | 96 |
| Sacramento, CA | WWTP | 24h composite | 1.6M | MU | 2025-08-28 | 18 |
| Southern California, CA | WWTP | 24h composite | - | MU | 2025-04-16 | 73 |
| West Boise and Lander Street Water Renewal Facility (Boise RWF), ID | WWTP | 24h composite | 270k | MU | 2024-11-04 | 57 |

**Figure 1.**
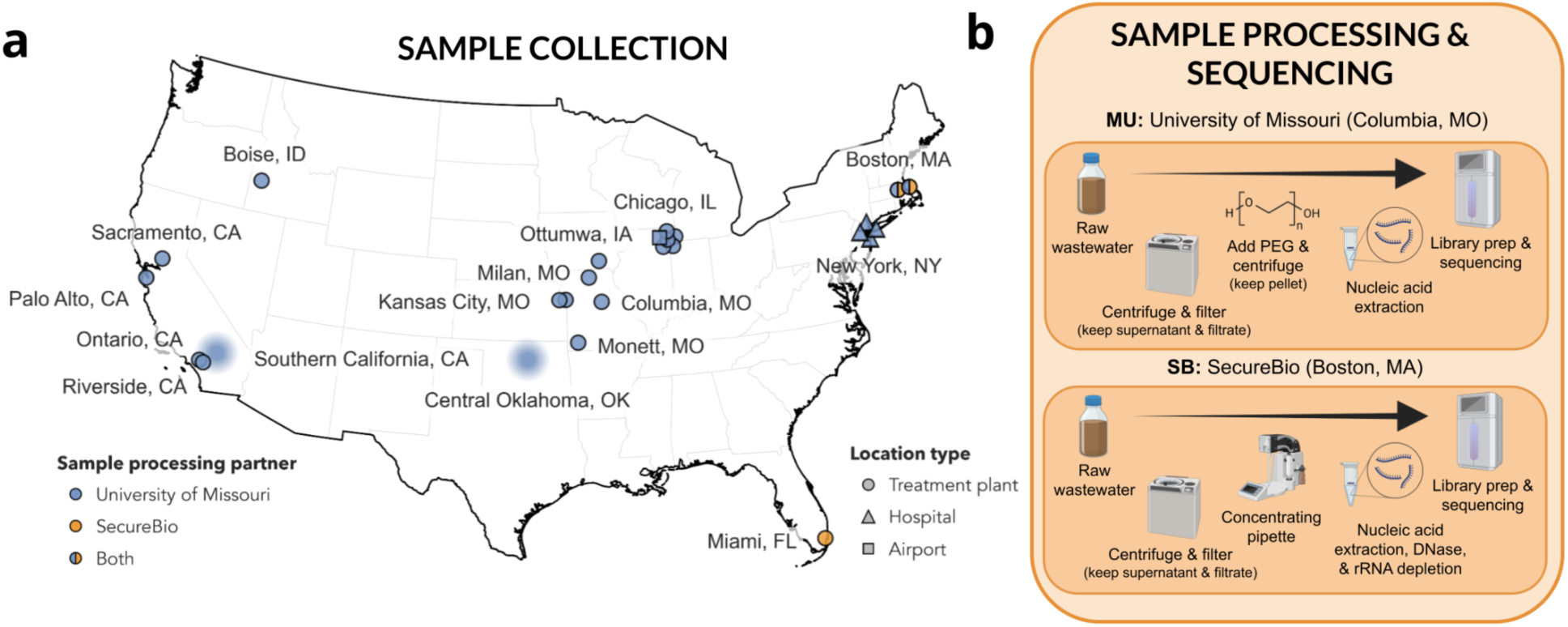
Overview of CASPER sampling network and laboratory workflows. (a) Geographic distribution of CASPER sampling sites included in this data release. The 27 sites span nine U.S. states; some metropolitan areas (Chicago, Kansas City, Boston, and New York City) include multiple sites. Marker locations for Southern California and Central Oklahoma are approximate to preserve site anonymity. Wastewater treatment plants labeled with a particular city may serve surrounding municipalities and communities. (b) High-level sample processing and sequencing workflows at the University of Missouri (MU) and SecureBio (SB) (see Methods for details).

After collection, local partners ship samples on ice packs to one of two sample processing and sequencing laboratories: MU or SB (Figure 1b). Each laboratory applies a distinct sample processing workflow optimized for viral enrichment and RNA sequencing (details in the Methods). In both workflows, teams perform viral concentration, nucleic acid extraction, cDNA synthesis, and Illumina library preparation. MU processed the majority of samples in this release (1,078 samples from 26 sites), while SB processed samples from Boston and Miami (128 samples from 3 sites) (Table 1; Figure 2a). Both laboratories independently process the same physical samples from Boston’s two Deer Island Treatment Plant (DITP) sites, enabling direct comparison of protocols at a single site.Most samples consist of 24-hour composite influent collected from municipal wastewater treatment plants (WWTP) at approximately weekly intervals, although sampling frequency varies by site (Figure 2a). A small number of sites, including Riverside WQCP, CA and Ontario, CA, collect samples twice weekly, whereas most others collect weekly. Samples from New York City (NYC) represent grab samples from hospital sewage, with one site collected from an indoor service hatch within the facility and three others from external manholes serving the hospitals exclusively. NYC samples were collected at more irregular intervals and often stored at 4°C for several months prior to sequencing. Samples from the CHI-C site consist of Moore swabs (Sikorski & Levine, 2020) collected from a manhole serving an airport. We selected sites based on established partnerships and to maximize geographic and population coverage across U.S. regions. Some sites, including Milan WWTF, MO and Ottumwa WPCF, IA receive industrial wastewater from livestock processing facilities, enabling monitoring of animal disease signals. Site names reflect preferences of local partners, and some sites opted not to disclose treatment plant names, locations, or other metadata.

**Figure 2.**
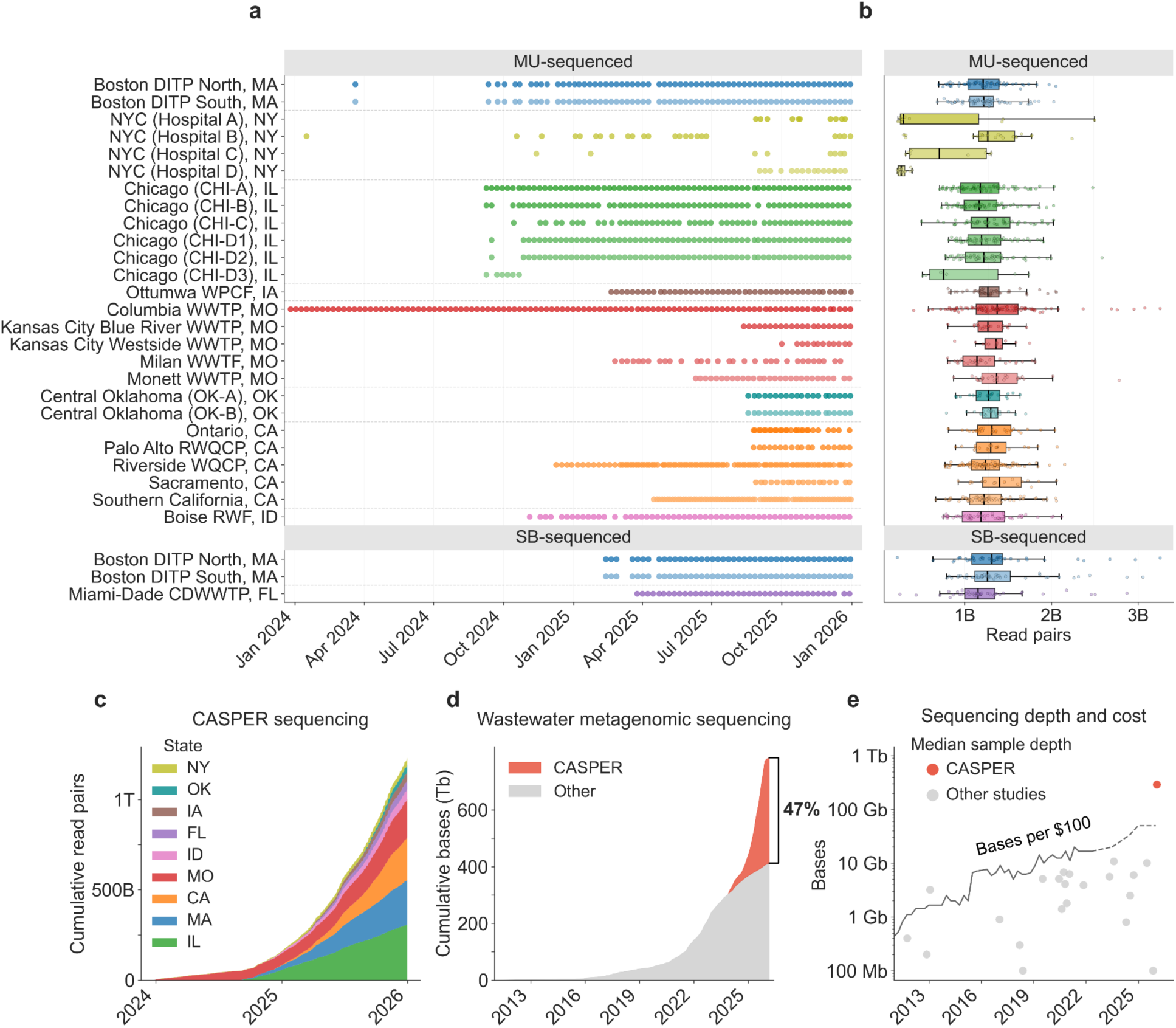
CASPER sequencing effort and scale. (a) Sample collection dates by site. (b) Sequencing depth per sample by site. (c) Cumulative read pairs generated by CASPER in this release over time. (d) CASPER contribution to all wastewater metagenomic sequencing data deposited on the NCBI Sequence Read Archive (SRA). Includes data from both the current release (PRJNA1247874; 358 Tb) and the previous Los Angeles data release (PRJNA1198001; 13 Tb). (e) Median sequencing depth (bases per sample) across studies with publicly available untargeted wastewater metagenomic sequencing data, compared to CASPER samples in this release, alongside the number of bases purchasable for $100 USD over time (line). Sequencing cost data through 2022 are from NHGRI (Wetterstrand, 2019; solid line); costs from 2023-present are estimated from observed sequencing output and pricing (Kaufman, 2025; dashed line).

All samples undergo deep, untargeted RNA sequencing on Illumina NovaSeq instruments, generating 2×150 bp paired-end reads and targeting approximately one billion read pairs per sample. NYC hospital samples sequenced December 2025 are the exception, targeting approximately 150 million read pairs per sample. Across the 1,206 samples included in this release, median sequencing depth is 986 million read pairs (range: 127M–3.25B; Figure 2b; Supplementary Figure S1), corresponding to a median of 290 gigabases (Gb) per sample (Figure 2e) and totaling 1.2 trillion read pairs (358 terabases) (Figure 2c, Supplementary Figure S2). This depth substantially exceeds that of other publicly available untargeted WW-MGS datasets, with median bases per sample approximately 27-fold higher than the next-deepest study identified in our literature survey (Figure 2e; Supplementary Table S1). Rapid declines in sequencing costs over the past decade have made deep sequencing at this scale feasible. Sequencing a typical CASPER sample would have cost approximately $4,000 USD in 2016, compared to approximately $500 USD in 2026 based on National Human Genome Research Institute (NHGRI) cost tracking data and internal estimates (Kaufman, 2025; Wetterstrand, 2019) (Figure 2e; Supplementary Table S2).

Median raw read quality score is Q39 (range: Q35–Q39) for both MU- and SB-processed samples. Median turnaround time from sample collection to delivery of sequencing data is 19 days (range: 9–657 days), varying by site and sequencing laboratory (Supplementary Figure S3. The longest turnaround times reflect NYC hospital grab samples collected during early pilot sampling and banked at 4°C prior to integration into routine processing; turnaround for these sites has since improved (Methods).

Following sequencing, data is transferred to Amazon Web Services (AWS) cloud storage and processed by the SB MGS analysis pipeline (Bradshaw et al., 2026). Partner reports and raw sequencing data are made available immediately to CASPER members, enabling them to run their own analyses. Sequencing data is periodically deposited in the NCBI SRA following masking of human-derived reads (Figure 1b). As of 2026-03-31, CASPER data account for approximately 47% of all wastewater metagenomic sequencing data on the SRA, including amplicon- and hybrid-capture-based datasets (Figure 2d; Supplementary Figure S2).

### Taxonomic composition of sequencing data

To rapidly characterize the high-level composition of CASPER sequencing data (Figure 3), we perform shallow taxonomic profiling on a subset of reads from each sequencing library. For each library, we randomly sample one million read pairs per sequencing lane, resulting in approximately one million read pairs for SB-processed samples and eight million for MU-processed samples. Reads are quality filtered, partitioned into ribosomal and non-ribosomal fractions, and classified using Kraken2 with the Standard reference database (Methods).

**Figure 3.**
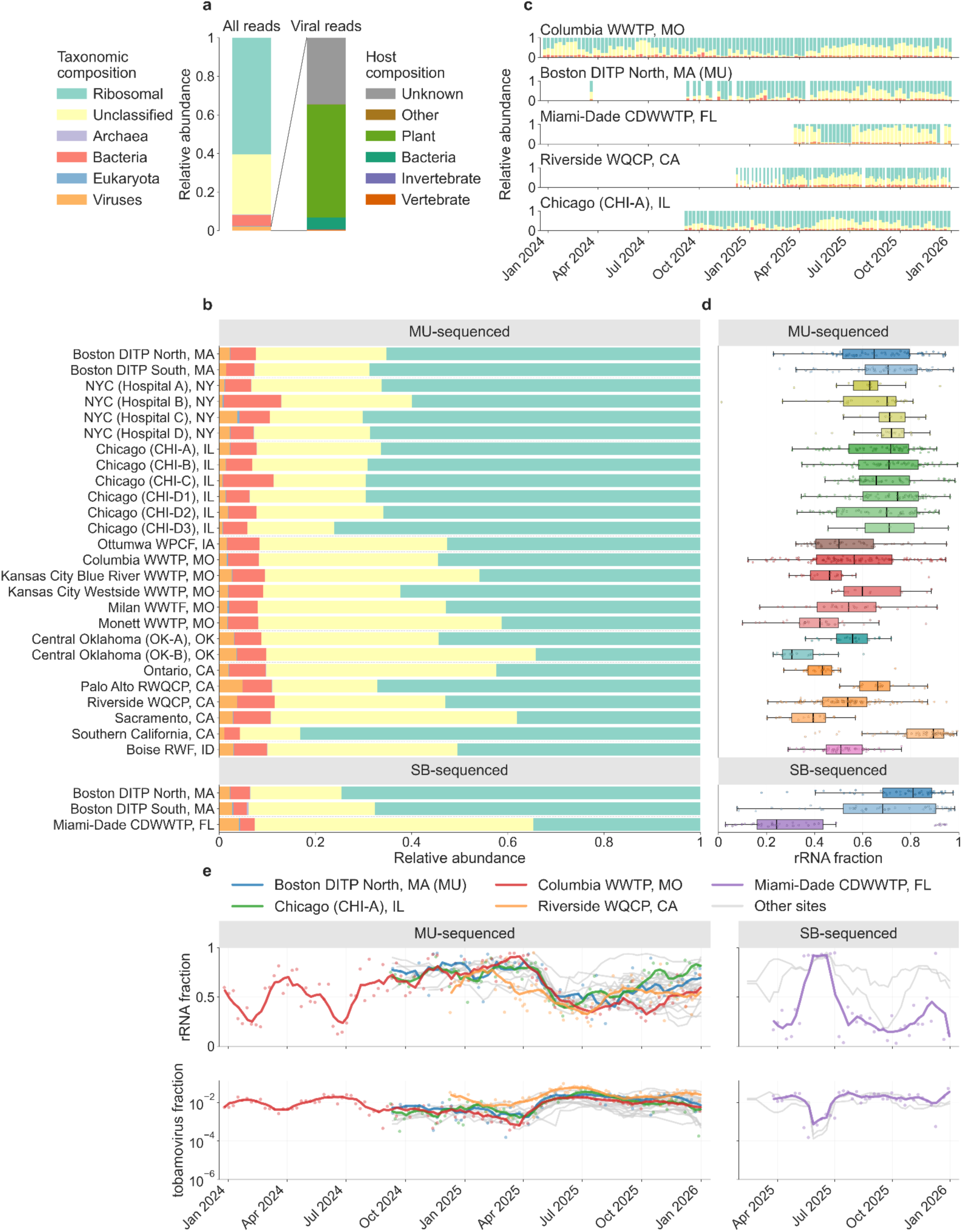
Taxonomic composition of CASPER sequencing data. (a) Aggregate composition across all samples, showing read classification by major taxonomic categories (left) and viral reads stratified by host category (right). (b) Aggregate composition by site and sequencing partner. (c) Temporal variation in taxonomic composition for selected sites. (d) Distribution of ribosomal RNA (rRNA) fraction by site. (e) rRNA fraction (top) and tobamovirus relative abundance (bottom) over time, with select sites highlighted; points show individual samples and lines show a centered 5-week moving average (Methods). The mid-2025 increase in rRNA fraction among SecureBio-processed samples reflects a temporary omission of ribodepletion reagent (see Methods). Tobamovirus is used in Figures 4c and 5 to normalize virus abundance trends in wastewater metagenomic sequencing data (Methods). Data in all panels derive from shallow taxonomic profiling of subsampled reads with Kraken2 against the Standard database; host assignments across all viral reads in (a) are derived from Virus-Host DB (Methods).

Across all samples, ribosomal RNA (rRNA) sequences constitute the largest fraction of reads. The median rRNA fraction is 61.1% (range: 1.2–99.5%) (Figure 3a-c). Among the remaining non-ribosomal reads, sequences that could not be classified by Kraken dominate (median 28.9%, range 0.14–91.9%) followed by bacteria (median 5.7%, range 0.39–48.1%). Viruses (median 1.6%, range 0.00–14.3%), eukaryotes (median 0.08%, range 0.00–3.1%), and archaea (median 0.03%, range 0.00–0.87%) contribute smaller fractions (Figure 3b).

High-level taxonomic composition varies over time, across sites, and between sequencing laboratories (Figure 3b; Supplementary Figure S4), motivating the use of normalization strategies. Metagenomic sequencing data is compositional, meaning that pathogen abundance expressed as a fraction of total reads (total read relative abundance) depends on the abundance of all other taxa present. As a result, fluctuations in highly abundant background components, whether from natural changes in the sewer microbiome or from protocol changes, can shift pathogen total read relative abundance independent of any change in pathogen concentration. The temporary omission of ribodepletion reagent among mid-2025 SB-processed samples is an example of the latter (Figure 3c, 3e; Supplementary Figure S5; Methods). Normalization to fecal indicator viruses, such as human diet-derived tobamoviruses, including pepper mild mottle virus (PMMoV) or tomato brown rugose fruit virus (ToBRFV), can correct for these fluctuations. Here, we normalize time-series pathogen abundance (Figures 4c and 5) by tobamovirus genus counts, which improves concordance with wastewater PCR and clinical trends relative to total read relative abundance (Supplementary Tables S3, S4; Justen, Zulli, et al., 2026). Figure 3e shows tobamovirus as a fraction of total reads, with counts scaled from the shallow taxonomic profiling subsample, in which tobamovirus reads were abundant (median 47,194 per sample, 95% of samples above 1,900; Supplementary Figure S6).

**Figure 4.**
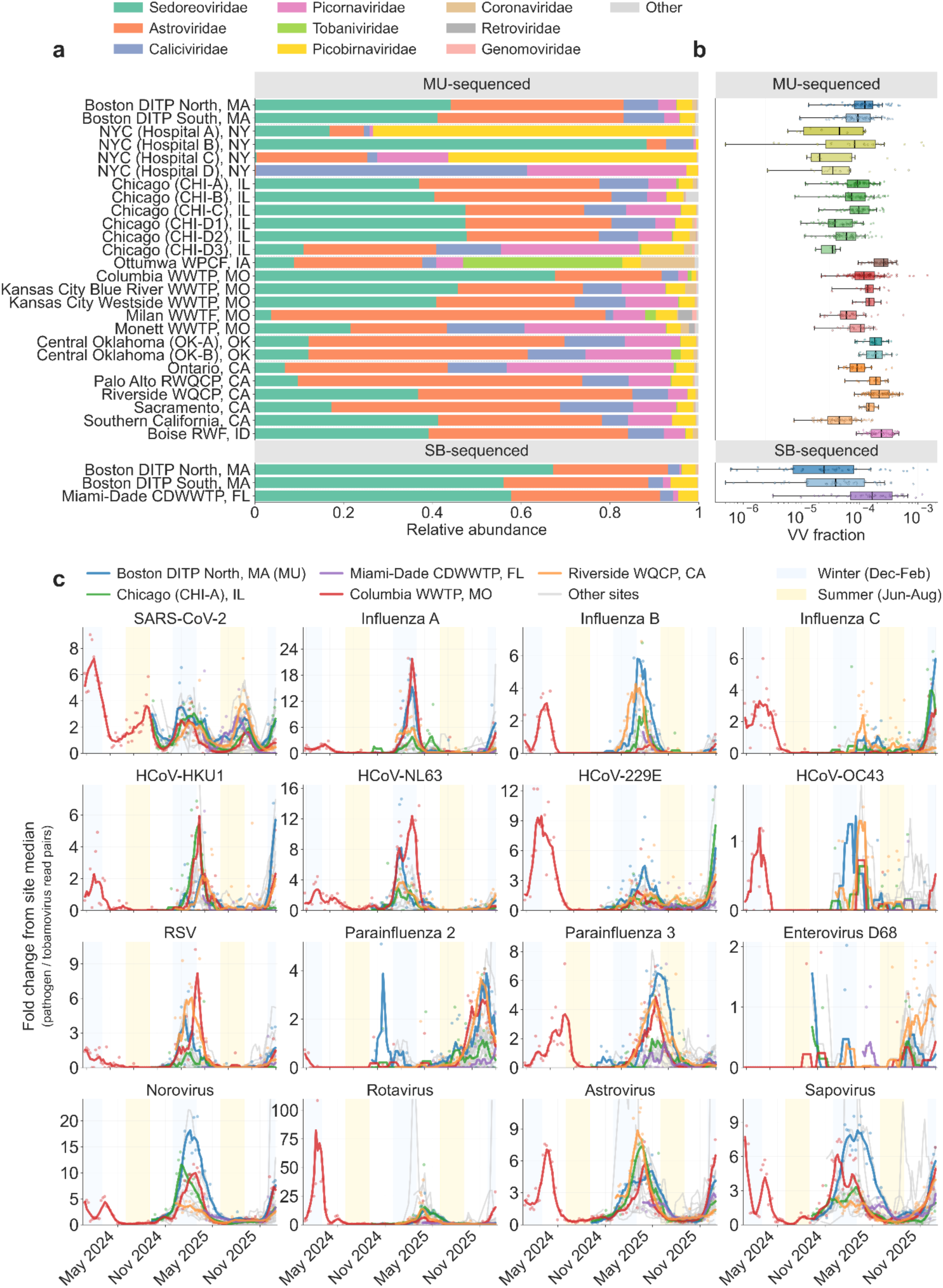
Vertebrate-infecting virus composition and temporal dynamics. (a) Family-level composition of vertebrate-infecting virus (VV) reads by site, stratified by sequencing partner. (b) Total VV read fraction by site. (c) Temporal patterns of select human-infecting viruses across CASPER sites. For each pathogen, abundance is normalized by tobamovirus genus counts from the shallow Kraken2 profiling (Methods) and expressed as fold change relative to the site-specific median of non-zero tobamovirus-normalized values. Respiratory viruses (top three rows) and gastrointestinal viruses (bottom row) are shown over time, with five representative sites highlighted in color and remaining sites shown in gray. For highlighted sites, points show individual MMWR-week observations and lines show a centered 5-week moving average (Methods); for remaining sites, only the smoothed series is shown. Y-axis limits are scaled such that the highest smoothed peak among highlighted sites falls at 75% of the axis height; values exceeding this range are clipped. A log-scale version showing tobamovirus-normalized abundance without fold-change normalization is provided in Supplementary Figure S10.

**Figure 5.**
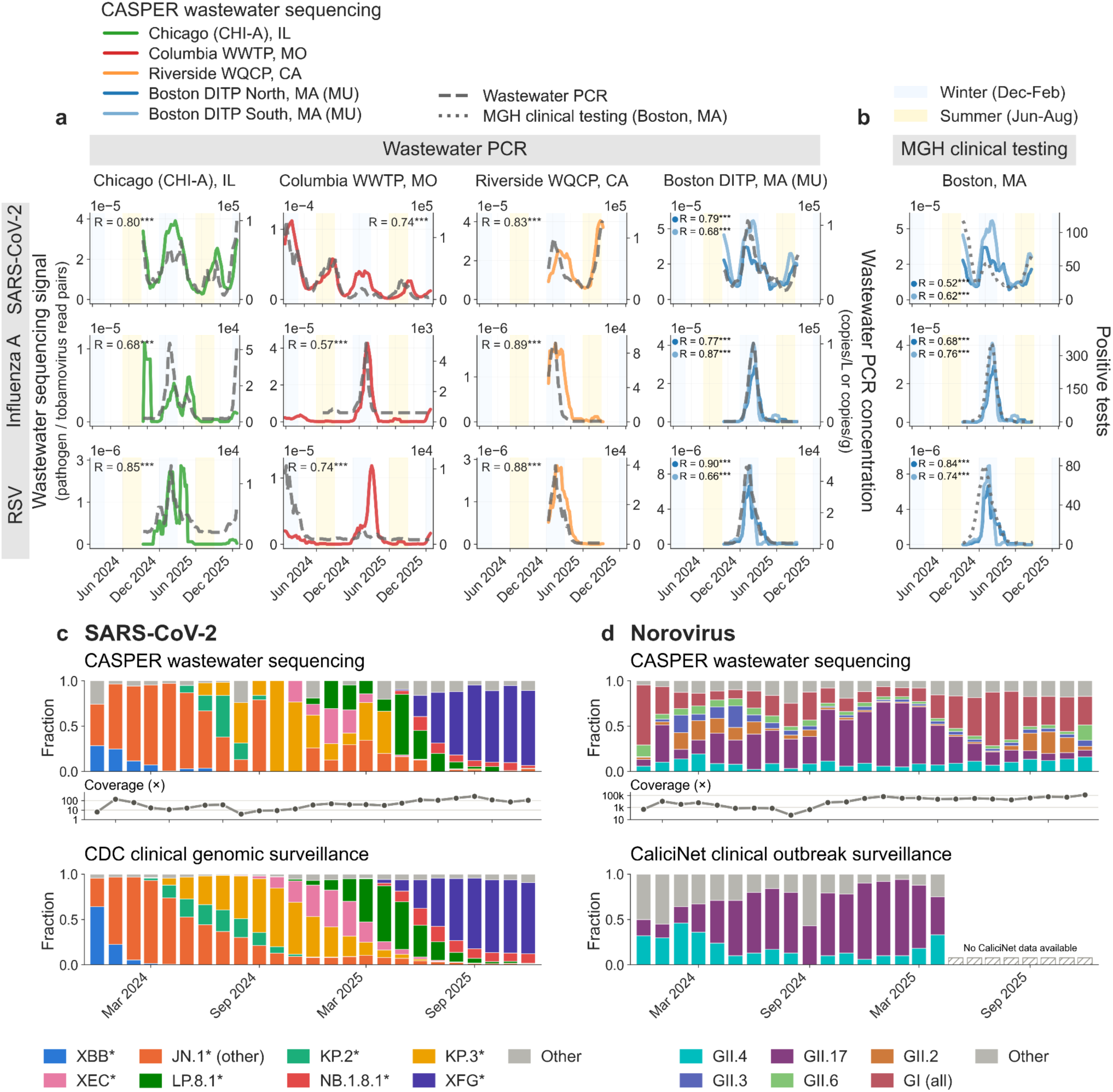
Agreement between wastewater metagenomic sequencing (WW-MGS) and established pathogen monitoring data. (a) WW-MGS abundance compared with digital PCR measurements from the National Wastewater Surveillance System (NWSS) and WastewaterSCAN (Boehm et al., 2026). WW-MGS abundance is normalized by tobamovirus genus counts; PCR measurements are shown as reported concentrations without normalization, in copies/L for influent or copies/g dry weight for settled solids depending on the site (Supplementary Table S10). (b) WW-MGS abundance from Boston Deer Island Treatment Plant (DITP) compared with clinical testing data from Massachusetts General Hospital (MGH). Clinical data represent positive counts from multiplex nucleic acid amplification tests performed on symptomatic patients as part of routine clinical care. For panels (a) and (b), Spearman correlation coefficients (R) were calculated using week-aggregated data with centered 5-week smoothing (Supplementary Tables S3, S4). *p < 0.05, **p < 0.01, ***p < 0.001. (c) SARS-CoV-2 lineage composition from CASPER data with reads pooled monthly across sites (top), mean genome coverage for each monthly pool (middle), and CDC clinical genomic surveillance for the same period (bottom; CDC, 2021). (d) Norovirus capsid genotype composition from CASPER data pooled monthly across sites (top), mean coverage across the VP1 region used for genotyping (middle), and CaliciNet outbreak surveillance (bottom; Barclay & Vinjé, 2025). CaliciNet reports GII.4 and GII.17 against a total-outbreak denominator, so remaining genotypes are shown as a single unresolved category; the published series ends in early 2025. MU: University of Missouri-sequenced.

Among all Kraken-identified viral reads, plant-infecting viruses, many associated with human diet, constitute the majority (58.6%), followed by viruses with unknown or ambiguous host assignment (34.7%), and bacteriophages (6.0%) (Figure 3a). The large unknown fraction likely reflects limited coverage of environmental bacteriophage lineages in Virus-Host DB, and true bacteriophage abundance is likely higher than reported here. Vertebrate-infecting viruses (VV) represent approximately 0.01% of all reads based on the Kraken2 shallow taxonomic profiling.

### Tracking vertebrate-infecting viruses

To sensitively identify vertebrate-infecting viruses (VV) reads across the full dataset, we analyze all sequencing data using SB’s Viral Metagenomics Pipeline (v3.0.1; index version 20250825; github.com/securebio/nao-mgs-workflow; Bradshaw et al., 2026). This Nextflow-based workflow, executed on AWS Batch, is designed to screen large sequencing datasets efficiently for VV sequences. We construct a VV genome database from Virus-Host DB and NCBI GenBank, excluding transgenic, recombinant, and unverified sequences, and masking reference genomes to reduce spurious alignments. We screen raw read pairs against the masked VV genome database using exact k-mer matching and retain any read pair with at least one match. We then adapter-trim and quality-filter candidate reads and screen them against common contaminant genomes, including human, cow, pig, carp, mouse, *E. coli*, and genetic engineering vectors. We align remaining reads to the VV genome database using Bowtie2, filter alignments by a length-normalized score threshold, and assign NCBI taxonomy identifiers using a custom lowest common ancestor (LCA) algorithm (Methods).

To assess pipeline sensitivity and specificity, we benchmarked the workflow on simulated reads from reference genomes of 93 notable VV taxa, drawn both from the pipeline’s reference database and from GenBank sequences deposited after that database was built, and from the most abundant non-VV taxa in CASPER wastewater (Methods). Reads from VV genomes present in the reference database were assigned to the correct genus 97.1% of the time and the correct species 92.0% of the time; for genomes of the same taxa absent from the database, the corresponding rates were 91.4% and 86.3% (Supplementary Figure S7; Supplementary Table S5, S6). Among 94.5 million read pairs simulated from non-VV genomes, 22 were assigned to a vertebrate-infecting virus, a false-positive rate of 2.3 × 10^-7^.

The dedicated VV pipeline identifies an aggregate of 0.025% of total reads across all samples as VV, approximately twice the number detected by the Kraken2 shallow taxonomic profiling. Enteric virus families dominate, including *Sedoreoviridae* (rotaviruses), *Caliciviridae* (noroviruses, sapoviruses), *Astroviridae*, and *Picornaviridae* (enteroviruses, rhinoviruses), consistent with the fecal origin of wastewater (Figure 4a). Respiratory virus families, including *Orthomyxoviridae* (influenza viruses), *Coronaviridae* (coronaviruses), *Paramyxoviridae* (parainfluenza viruses) and *Pneumoviridae* (RSV), occur at lower relative abundance but remain readily identifiable. Among samples, VV reads constitute a median of 0.01% of reads (range: 0.00–1.08%), varying by site and sequencing partner (Figure 4b; Supplementary Figure S8). Among paired samples from Boston DITP, processed by both SB and MU, the MU protocol yields a higher VV fraction (median ratio 1.6× at DITP South and 5.4× at North), although normalized pathogen trends remain correlated (Supplementary Figure S9; Supplementary Table S7)

Hospital sites and sites receiving animal agricultural inputs show distinctive VV composition signatures (Figure 4a). Ottumwa, IA, which receives industrial wastewater from a livestock processing facility, shows elevated *Tobaniviridae* and *Coronaviridae* consistent with porcine viruses (buldecovirus, pedacovirus, torovirus). Hospital samples representing grab collections taken closer to the source, with less dilution from other sewershed inputs, and from populations enriched for sick or immunocompromised individuals (Renfro et al., 2026), also diverge from municipal influent patterns, with NYC Hospital A and C showing elevated *Picobirnaviridae* (human picobirnavirus) and *Picornaviridae* (enterovirus) abundance.

We next examined time-series abundance trends for respiratory and enteric viruses in CASPER data (Figure 4c). Absolute assigned read counts vary by orders of magnitude across targets, with median counts among samples with at least one assigned read pair ranging from 1,728 to 29,170 for the enteric pathogens shown and from 2 to 82 for the respiratory pathogens (Supplementary Figure S11; Supplementary Table S8). Despite the low number of absolute assignments, tobamovirus-normalized abundance trends (Figure 4c) tracked known epidemiological patterns, such as a winter–spring spike for many respiratory pathogens and broad concordance in pathogen trends across U.S. sites, with observable leads and lags between sites likely reflecting regional outbreak timing. Among respiratory viruses, influenza A and B, RSV, and seasonal human coronaviruses display consistent seasonal peaks in winter and spring. Enteric viruses including norovirus, rotavirus, astrovirus, and sapovirus similarly show cross-site concordance, with synchronized winter and spring peaks. Enterovirus D68 shows elevated abundance across California sites in fall 2025, consistent with regional wastewater PCR data (WastewaterSCAN, 2025).

Other pathogens show less consistent year-to-year patterns. At Columbia, MO, one of the few sites with multi-year coverage, parainfluenza 3 shows spring peaks in both 2024 and 2025, whereas parainfluenza 2 is detected in fall 2025 but absent the previous year. Influenza C, which, unlike influenza A and B, is rarely a clinical testing target, was abundant at Columbia, MO, in winter 2023–2024, largely absent the following winter, and detected again across multiple sites in winter 2025–2026.

To assess concordance with established pathogen monitoring methods, we compared CASPER WW-MGS measurements with publicly available wastewater digital PCR data from the CDC NWSS data portal, which aggregates data generated by CDC, state and local health departments, and by WastewaterSCAN (Boehm et al., 2026), at sites with overlapping coverage (Supplementary Table S9). For SARS-CoV-2, influenza A, and RSV, WW-MGS and PCR measurements show weak to strong positive correlations at sites with extended time series (Figure 5a; Spearman R = 0.57–0.90). Correlations, p-values, and number of overlapping weeks for all sites are reported in Supplementary Figure S12 and Supplementary Table S3; wastewater PCR assay metadata is reported in Supplementary Table S10.

For Boston, we compared WW-MGS-derived abundance with clinical case counts from testing performed at the Massachusetts General Hospital (MGH), a large academic medical center whose patient catchment substantially overlaps the DITP sewershed (Figure 5b). WW-MGS measurements from both the DITP North and South systems correlate with MGH positive test counts for all three viruses (R = 0.52–0.84; Supplementary Figure S13, Supplementary Table S4). Together, these comparisons indicate that CASPER data tracks population-level pathogen trends alongside established monitoring methods, while detecting a broader range of pathogens.

CASPER data also supports genotype- and lineage-level analysis. While low coverage of many pathogens, particularly respiratory viruses, often hinders lineage or genotype assignment from a single sample, pooling reads across sites can recover national-scale pathogen variant dynamics. For SARS-CoV-2, CASPER captures the successive transitions from JN.1 to KP.3 and eventually XFG, at approximately the same time as in CDC clinical genomic surveillance (Figure 5c) (CDC, 2021). Composition is noisier than the clinical series, particularly in earlier months when fewer sites were contributing (Figure 2a), and does not resolve circulating lineages at the same granularity. Norovirus is roughly two orders of magnitude more abundant in these data, and genotype composition tracks the rise of GII.17 relative to GII.4 through the 2024–25 season reported by CaliciNet (Figure 5d) (Barclay & Vinjé, 2025). Both genotypes account for a smaller share of the total in the wastewater data than in CaliciNet, most likely because CaliciNet counts recognized outbreaks, which GII.4 and GII.17 dominate through efficient person-to-person transmission, whereas genogroup I is more often implicated in point-source events less likely to be recognized and reported.

## Discussion

Wastewater metagenomic sequencing represents a promising tool for public health and biosecurity. Here, we report the first major release of WW-MGS data generated by the U.S.-based CASPER consortium, covering two years, 13 million people, 1,206 samples, 358 terabases, and 27 sites across nine U.S. states. The release substantially expands the volume of available wastewater sequencing data and demonstrates the feasibility of scalable workflows for deep untargeted wastewater sequencing.

Time-series national-scale data from CASPER enable broad detection and tracking of pathogens. By adopting a pathogen-agnostic approach, CASPER extends monitoring beyond common targets such as SARS-CoV-2, influenza, and RSV to hundreds of viral taxa, including rare or emerging threats. Separate work has reported how CASPER data from Columbia, MO, identified H5N1 influenza in early 2024, before any infected herd was confirmed in Missouri (Rushford et al., 2025), and detected measles in Chicago, IL, wastewater, linked by genomic analysis to a single confirmed infection in a sewershed serving more than one million people (Poretsky et al., 2026).

We anticipate that public CASPER data will have broad utility for pathogen surveillance, including retrospective outbreak investigations; WW-MGS modeling and statistical methods development (Grimm, Kaufman, et al., 2025; Justen, Zulli, et al., 2026); evaluation of alternative pathogen monitoring approaches, such as air sampling (Justen et al., 2025; Minor et al., 2023), blood supply sampling (Justen, Bhasin, et al., 2026; Kean et al., 2026), airport and aircraft wastewater surveillance (Friedman et al., 2025; McLaren et al., 2026; St-Onge et al., 2025), and hybrid-capture sequencing (Kantor & Jiang, 2024; Tisza et al., 2023; Wolfe et al., 2026); and the development of novel detection approaches, such as anomalous growth detection (The Nucleic Acid Observatory Consortium, 2021), genetic engineering detection (Crook et al., 2022; Kaufman, 2024; Tay et al., 2024), AI/ML methods (Liu et al., 2025; Ren et al., 2020), and reference-, marker-, and functional-based approaches (Balaji et al., 2022; Camargo et al., 2024; Luebbert et al., 2025; Wittmann et al., 2025). CASPER data also supports further exploration of microbial dark matter through large-scale metagenomic assembly (Edgar et al., 2022; Fiamenghi et al., 2025; Kantor et al., 2026; Neri et al., 2022; Worp et al., 2025).

A key distinguishing feature of CASPER is its sequencing depth. Most existing untargeted WW-MGS studies have used substantially lower per-sample depth, limiting their ability to resolve rare viral targets. We show that some of these sensitivity limitations can be overcome by sequencing more deeply. Even so, sensitivity constraints remain (Grimm, Kaufman, et al. 2025), particularly for pathogens with low prevalence or low fecal shedding, and genome coverage for most pathogens remains partial, limiting strain-level characterization.

The untargeted WW-MGS deployed by CASPER has other limitations. It requires costly sequencing infrastructure, and the ongoing cost of deep sequencing exceeds that of more targeted platforms. Turnaround time also tends to be longer, driven by longer laboratory workflows and sequencing duration. The protocols used in CASPER, while more untargeted than other current wastewater biosurveillance approaches, are designed primarily to capture RNA, potentially making detection of certain DNA viruses and cellular genomes more challenging, though DNA is retained in libraries prepared without DNase treatment. The scale of data generated also requires robust compute infrastructure for storage and processing. However, continued declines in sequencing cost, optimization and automation of sample processing, and development of efficient bioinformatic workflows are making WW-MGS an increasingly tractable platform for routine pathogen-agnostic detection.

CASPER will continue to pursue these improvements, aiming to serve as a real-time pathogen surveillance resource. Within the consortium, partners share data as results become available, and agreements cover the use and reporting of findings. Bulk data uploads from a growing number of public CASPER sites to the SRA have continued since this release and currently occur on a quarterly schedule.

In sharing this resource, we wish to highlight that interpreting WW-MGS data for public health and biosecurity requires careful consideration of nucleic acid sources beyond active human infection. These sources include sample contamination, laboratory and industrial waste, stormwater intrusion, animal infections, diet-associated contributions, and vaccine shedding. This complexity can make it challenging to determine whether detected sequences originate from human infections or other sources, especially when limited sequencing coverage permits only partial strain characterization (Ameaka et al., 2026). Investigation of flagged sequences, often based on only a small number of unique reads, is therefore nuanced, and determining whether a signal represents an actionable finding requires careful consideration of the environmental context. This perspective guides our communications with stakeholders, and we encourage others to bear it in mind.

We invite researchers, public health agencies, and other interested parties to engage with this growing resource. As the CASPER network continues to expand, we welcome inquiries from potential collaborators at.

## Methods

### Sample processing and sequencing at the University of Missouri

#### Sample collection and transport

##### Composite samples

Influent wastewater was collected from most locations as a 24-hour composite sample. Samples were aliquoted into three 50 mL conicals (150 mL total), shipped overnight with ice packs in insulated containers, and stored at 4°C until processing. Samples were processed in batches, typically within one week of receipt. Chicago composite samples CHI-D1, D2, and D3 represent different catchments served by the same wastewater treatment plant.

##### Moore swab samples

CHI-C samples were collected using a Moore swab deployed for 24 hours in a manhole serving an airport. Wastewater was recovered by manual compression of the swab into a collection bag, yielding approximately 30–40 mL of sample which was transferred to a 50 mL conical for shipment.

##### Grab samples

NYC hospital samples were collected via grab sampling from four facilities, with approximately 100 mL collected per sample and shipped in two 50 mL conicals. Hospital A samples were collected from an indoor service hatch within the facility at approximately 5:00 AM. Hospitals B, C, and D samples were collected from external manholes exclusively serving the hospital facilities: Hospital B at approximately 12:00 PM, Hospital C at approximately 9:00 AM, and Hospital D between 8:00–11:00 AM. The Hospital B manhole also receives stormwater from an adjacent dock area; Hospitals C and D manholes have minimal rainwater input. These samples were initially collected as an exploratory effort, which accounts for extended processing times: samples were stored at 4°C prior to shipping to MU, with a median time from collection to sequencing of 62 days (range: 21–657 days). We expect future turnaround times for these sites to align with other locations in the network.

#### Viral concentration and nucleic acid extraction

Samples were centrifuged at 2000 xg for 5 minutes at 4°C and the supernatant was filtered through 0.22 µm filters (Millipore, SCGP00525). Filtrates (35 mL) were mixed with 12.5 mL of 50% (w/vol) PEG (Research Products International, P48080-1000.0) and 1.2 M NaCl and incubated for 1 h at 4°C. The mixture was then centrifuged at 12,000 xg for 2 h at 4°C to pellet viruses. After decanting the supernatant, nucleic acids were extracted from viral pellets using the QIAamp Viral RNA Mini Kit (Qiagen, 52906) according to the manufacturer’s instructions, omitting carrier RNA and DNase treatment. Extractions prior to 2025-06-04 were performed using a Qiagen QIAcube; subsequent extractions were performed manually in a biosafety cabinet.

A subset of Columbia, MO, samples collected between 2024-01-02 and 2024-03-19 underwent enzymatic pretreatment prior to filtration to reduce bacterial ribosomal RNA. Samples were incubated with 10,000 ng RNase A (ThermoFisher, R1253), 2 units of TURBO DNase (Invitrogen, AM1907), and 4.75 mL of buffer containing 5 mM CaCl₂ and 25 mM MgCl₂. Some samples additionally received 30 mM EDTA to promote dissociation of rRNA from ribosomal complexes. For all nuclease-treated samples, 400 units of RNaseOUT™ Recombinant Ribonuclease Inhibitor (Invitrogen, 10777019) were added prior to RNA extraction. Treatment conditions varied over time: samples from 2024-01-02 through 2024-01-09 were processed in triplicate under three conditions (no treatment, nuclease only, nuclease plus EDTA); samples from 2024-01-16 through 2024-02-20 were processed under two conditions (nuclease only, nuclease plus EDTA); and samples from 2024-02-27 through 2024-03-19 were processed under two conditions (no treatment, nuclease only). These treatments did not substantially affect rRNA content or viral detection (Rushford et al., 2025); all samples collected after 2024-03-19 were therefore processed without enzymatic pretreatment. For samples processed under multiple conditions, sequencing data were merged prior to public release on SRA.

To evaluate algorithms designed to detect engineered sequences, we spiked 1 µL of a previously described NL4-3–derived HIV construct (containing a CMV-driven puromycin resistance gene and lacking *env*, *nef*, *vif*, and *vpr*) into the Columbia, MO, sample collected on 2024-07-09, prior to filtration (Robinson et al., 2022).

#### RT-qPCR quantification of PMMoV

Successful concentration of viral RNA were evaluated prior to library preparation using comparative RT-qPCR targeting PMMoV. RT-qPCR assays utilized PMMoV specific forward (5’-GGCGTAGATCCATTGGTGC-3’) and reverse (5’-CGAACCTTCCTCCTTTGATG-3’) primers, each prepared at a stock concentration of 100 µM, along with a TaqMan probe (VIC-5’ GCTGTGGTTTCAAATGAGAGTGG 3’-QSY) prepared at 100 µM. Each 20 µL RT-qPCR reaction consisted of 5 µL of TaqPath 1-Step RT-qPCR Master Mix GC (Applied Biosystems, A15299), forward and reverse primers at a final concentration of 500 nM each, the TaqMan probe at 125 nM, and 5 µL of the RNA template. Amplification was performed on an Applied Biosystems 7500 Fast Real-Time PCR System using the following cycling conditions: reverse transcription at 48 °C for 15 min, initial denaturation at 95 °C for 10 min, and 40 amplification cycles of 95 °C for 15 s followed by 60 °C for 1 min.

#### Library preparation and sequencing

Total RNA concentrations were measured using the Qubit RNA High Sensitivity Assay prior to library construction. RNA input amounts were normalized based on Qubit quantification, targeting 10–15 ng of RNA in a final volume of 9.5 µL. For samples with RNA concentrations below the detection limit, the full 9.5 µL was used as input. Libraries were prepped using the Illumina Stranded mRNA Prep (Illumina, 20040534) and index adapters sets A–D (Illumina, 20091655, 20091657, 20091659, and 20091661). Sequencing libraries were prepped according to the manufacturer’s protocol with minor modifications listed below.

The mRNA capture step was omitted, and library preparation began directly with RNA fragmentation followed by cDNA synthesis. Each normalized RNA sample was divided into two equal fractions with one fraction undergoing fragmentation to reduce longer RNA molecules to a size range suitable for sequencing, and the second fraction left unfragmented to preserve RNA already within the target size range. Following fragmentation, both fractions were recombined prior to cDNA synthesis.

Final indexed libraries were size-selected during the post-PCR cleanup using a two-step AMPure XP bead protocol (Beckman Coulter, A63881). Initially, 30 µL of AMPure XP beads were added to 50 µL of post-indexing PCR products to bind larger library fragments. After a 5-minute incubation, the supernatant was transferred to a new tube and combined with an additional 20 µL of AMPure XP beads to capture shorter fragments. These shorter bead-bound libraries were washed, eluted in 30 µL of resuspension buffer, and used for final pooling.

Sequencing was performed using Illumina NovaSeq platforms with paired-end 2×150 bp reads. Libraries generated through 2024-04-16 were sequenced on a NovaSeq 6000, while libraries prepared after were sequenced on a NovaSeq X system. The transition to NovaSeq X was associated with improved quality scores and a transient decrease in GC content for Columbia, MO, samples (Supplementary Figures S14, S15), which resolved as the sequencing core optimized performance on the new instrument.

While most libraries were sequenced at 2×150 bp, a small number of samples were sequenced with different read lengths due to pilot runs or inadvertent sequencing core errors (Supplementary Figure S16): (1) Columbia, MO, samples from 2023-12-26, 2024-01-02, and 2024-01-09 include a small fraction of reads at 2×75 bp from initial shallow sequencing, with the majority at 2×150 bp from subsequent deep sequencing; (2) Columbia, MO, samples from 2024-06-11 through 2024-07-02 include approximately 900M reads at 2×100 bp alongside 2×150 bp reads, as part of diagnostic sequencing to investigate the transient GC bias; (3) Columbia, MO, samples from 2024-08-13 through 2024-09-10, Boston DITP North and DITP South from 2024-09-11, and CHI-A, CHI-B, and CHI-D3 from 2024-09-08 were inadvertently sequenced at 2×100 bp.

Libraries from samples collected prior to 2024-09-24 were generated by the University of Missouri Genomics Technology Core; samples collected after that date were processed in the Department of Molecular Microbiology and Immunology at the University of Missouri.

### Sample processing and sequencing at SecureBio

#### Sample collection and transport

Wastewater samples consisting of 1 L 24-hour composite influent from Miami-Dade CDWWTP and Boston DITP North and South were shipped to SecureBio for processing and sequencing. Boston samples were transported via courier in coolers containing ice packs. Miami samples were shipped overnight via FedEx in insulated containers to maintain temperature stability. Upon receipt, all samples were processed either immediately or within 24 hours. A 200 mL aliquot from each bulk sample was used for downstream processing.

#### Viral concentration, nucleic acid extraction, and rRNA depletion

Viral concentration and nucleic acid extraction were performed as described in (Machtinger et al., 2025). To facilitate dissociation of viral particles from solid matter, Tween 20 (Sigma Aldrich, P7949) was added to the 200 mL aliquot to a final concentration of 0.1%. Samples were vortexed at medium speed for approximately 1 minute, followed by sonication in a water bath at 40 kHz for 1 minute.

Following physical dissociation, samples were clarified by centrifugation at 10,000 xg for 5 minutes at 4°C using an Eppendorf 5920R centrifuge with a FA-6×250 fixed-angle rotor. The supernatant was decanted and filtered through a 0.45 µm polyethersulfone (PES) vacuum filtration unit (VWR, 10040-470) to remove remaining bacterial cells and large debris.

Viral concentration was performed using the InnovaPrep Concentrating Pipette Select with an Ultra Concentrating Pipette Tip (CPT; InnovaPrep, CC08004-100), following the manufacturer’s recommended protocol adjustments for wastewater processing (Concentrating Pathogens from Raw and Primary Wastewater Using the InnovaPrep® Concentrating Pipette, Protocol (Revision E), n.d.)). Samples were eluted using InnovaPrep Buffer-Tris fluid (InnovaPrep, HC08005) directly into a collection tube containing 400 µL of DNA/RNA Shield (Zymo Research, R1200-125) to achieve a 1:1 ratio of eluate to shield, resulting in a final concentrate volume of approximately 800 µL.

Total nucleic acids were extracted from the resulting concentrate using the Quick-Viral™ RNA/DNA Miniprep Kit (Zymo Research, D7021) according to the manufacturer’s instructions, scaled for the volume of the concentrated sample. To prepare samples for RNA sequencing, extracts were treated with the TURBO DNA-free™ Kit (Thermo Fisher Scientific, AM2238) to remove genomic DNA. Bacterial ribosomal RNA was subsequently depleted using the Pan-Bacterial RiboPOOLs kit (siTOOLs Biotech, dp-K096-00026). Due to a reagent preparation error, ribodepletion probes were omitted from samples processed between May 13 and July 1, 2025 (inclusive; Supplementary Table S11). Sequencing data from these samples exhibit a higher fraction of ribosomal reads and reduced effective depth for non-ribosomal targets (Supplementary Figure S5).

#### Library preparation and sequencing

Total RNA concentrations were quantified using the Qubit RNA High Sensitivity Assay (ThermoFisher Scientific, Q32855). RNA sequencing libraries were constructed using 5 µL of ribodepleted RNA as input. Library preparation was performed using the xGen RNA Library Prep Kit (Integrated DNA Technologies, 10009814) with IDT Unique Dual Index (UDI) PCR primers (Integrated DNA Technologies, 10005922) according to the manufacturer’s instructions. Final amplified libraries were pooled at equimolar concentrations (2–4 nM) and submitted to the

Broad Clinical Labs (Broad Institute, Cambridge, MA, USA) for sequencing on the NovaSeq X platform (Illumina, San Diego, CA, USA) at 2×150 bp.

### Bioinformatic processing

Sequencing data was analyzed using SecureBio’s Viral Metagenomics Pipeline (v3.0.1; index version 20250825; github.com/securebio/nao-mgs-workflow; Bradshaw et al., 2026), implemented in Nextflow (v25.10.0; Di Tommaso et al., 2017) and executed on AWS Batch. For this manuscript, the pipeline performs two primary analyses: (1) shallow taxonomic profiling on a subset of reads, and (2) deep VV profiling on all reads. Processing requires a median of 18 CPU-hours per sample and approximately 93 GB of peak memory. Compressed sequencing data occupies a median of 140 GB per sample, or approximately 14 GB per 100 million read pairs.

#### Preprocessing and quality control

Total read counts were computed from raw demultiplexed paired-end FASTQ files for each index group. For quality assessment and taxonomic profiling, read pairs were randomly subsampled to approximately one million per index group using Seqtk (v1.5, Li, 2013). For sequencing libraries spanning multiple lanes or deliveries, this results in several million subsampled read pairs per library (e.g., 8 million for a library sequenced across 8 lanes). Subsampled reads were adapter-trimmed and quality-filtered with FASTP (v0.23.4; parameters: ‘--cut_front --cut_tail --correction --detect_adapter_for_pe --trim_poly_x --cut_mean_quality 20 --average_qual 20 --qualified_quality_phred 20 --low_complexity_filter’; Chen et al., 2018). Quality filtering removed a median of 0.5% (range 0.2–25.9%) of subsampled reads for MU-sequenced libraries and 10.1% (range 1.2–44.7%) for SB-sequenced libraries (Supplementary Figure S17). Quality metrics such as mean GC content (Supplementary Figure S14) and quality scores (Supplementary Figure S15) were generated using FASTQC (v0.12.1; Andrews, 2010) and aggregated with MultiQC (v1.21; Ewels et al., 2016).

#### Shallow taxonomic profiling

Subsampled, adapter-trimmed reads were partitioned into ribosomal and non-ribosomal fractions using BBDuk (BBTools v39.01; parameters: ‘k=27 minkmerfraction=0.4’; Bushnell, 2014) against the SILVA ribosomal RNA databases (v138.2; Chuvochina et al., 2025): the SSU (Small Subunit; 16S/18S rRNA) NR99 and LSU (Large Subunit; 23S/28S rRNA) NR99 reference sets. Both fractions were independently classified with Kraken2 (v2.1.3; Wood et al., 2019) using the Standard reference database (k2_standard_20250714), which includes RefSeq complete genomes for bacteria, archaea, and viruses, the human reference genome (GRCh38), and UniVec_Core vector sequences. Ribosomal and non-ribosomal classifications were reported separately.

For aggregate taxonomic composition analysis (Figure 4), we combined ribosomal and non-ribosomal Kraken2 classifications into six categories: (1) Ribosomal—all rRNA reads regardless of taxonomic classification; (2) Bacteria (non-rRNA only); (3) Archaea (non-rRNA reads only); (4) Eukaryota (non-rRNA only); (5) Viruses (non-rRNA only); and (6) Unclassified—non-rRNA reads that were either unclassified by Kraken2 or assigned directly to the root or cellular organisms without more specific classification. Reads assigned to NCBI’s "unclassified entries" or "other entries" were excluded as ambiguous. Site-level compositions were calculated by summing estimated read counts (category fraction × total reads) across all libraries at each site, then normalizing to proportions, effectively weighting each library’s contribution by its sequencing depth.

Taxonomic category fractions and rRNA fractions were computed relative to reads passing quality filtering (the denominator for Kraken2 classification). For the six-category composition analysis (Figure 3a-c), category fractions were normalized to sum to 1. Tobamovirus clade counts use the total subsampled reads (pre-quality filtering) as denominator.

#### Vertebrate-infecting virus identification

All reads from each index group underwent deep profiling for vertebrate-infecting viruses:

1. **K-mer screening.** Raw read pairs were screened against a masked database of VV genomes using BBDuk (parameters: ‘k=24 minkmerhits=1’), retaining any read pair containing at least one 24-mer exact match to any VV genome. This rapid initial filter discards the vast majority of non-viral reads while retaining putative VV reads for more computationally intensive downstream analysis.
2. **Read cleaning.** Putative VV reads were adapter-trimmed and quality-filtered with FASTP (parameters as above), followed by additional adapter trimming with Cutadapt (v5.0; parameters: ‘-b -B -m 20 -e 0.33 --action=trim’), which trims adapters from either end of either read and discards read pairs where either read falls below 20 bp after trimming.
3. **Alignment to VV database.** Cleaned reads were aligned to the VV genome database using Bowtie2 (v2.5.4; parameters: ‘--local --very-sensitive-local --score-min G,0.1,19 -k 10’; Langmead & Salzberg, 2012), retaining up to 10 alignments per read to enable subsequent lowest common ancestor (LCA) analysis.
4. **Contaminant removal.** Reads aligning to the VV database were then screened against common contaminant genomes using Bowtie2 (parameters: ‘--local --very-sensitive-local’), sequentially filtering against human (GRCh38), cow, pig, carp, mouse, *E. coli*, and common genetic engineering vectors. Reads mapping to any contaminant were discarded.
5. **Score filtering.** Surviving alignments were filtered by a length-normalized alignment score threshold (alignmentScore / ln(readLength) ≥ 20) to remove low-confidence alignments.
6. **Taxonomic assignment via LCA.** Reads passing score filtering were assigned NCBI taxids using a custom LCA algorithm. For reads with multiple alignments, the algorithm computes the LCA across all aligned taxa, with special handling for unclassified taxa (those whose names contain "unclassified" or "sp.") and artificial sequences. Unclassified taxa are included in LCA calculation only if they have the highest alignment score and are not tied with any classified taxon. LCA assignments are computed separately for natural sequences, artificial sequences, and all sequences combined; natural-only LCA assignments are used for downstream analysis.

#### Vertebrate-infecting virus genome database construction

The VV genome database was constructed by the pipeline’s INDEX workflow as follows: (1) vertebrate-infecting virus taxids were obtained from Virus-Host DB (daily release, 2025-08-25) and NCBI Taxonomy (downloaded 2025-08-25); (2) this list was expanded to include all descendant taxonomy IDs; (3) all corresponding viral genome sequences were downloaded from NCBI GenBank (downloaded 2025-08-25) using ncbi-genome-download (v0.3.3; Blin, 2023); (4) sequences were filtered to exclude entries with header keywords indicating transgenic, mutant, recombinant, unverified, or draft sequences, as well as specific accessions previously identified as contaminated or erroneous; (5) remaining genomes were masked using BBDuk in two passes to reduce spurious alignments: first, adapter sequences were masked (parameters: ‘k=20 hdist=3 mink=8 hdist2=1 mask=N’) along with low-entropy regions (parameters: ‘entropy=0.5 entropymask=t’); second, homopolymer runs of ≥10 bp were masked (parameters: ‘k=10 hdist=0 mask=N’).

#### Host annotation

Viral taxa were annotated with host infection status using a custom algorithm that propagates host information through the NCBI viral taxonomy based on Virus-Host DB annotations (Mihara et al., 2016). The algorithm assigns each viral taxon an infection status for each host group of interest: MATCH (affirmatively infects the host), INCONSISTENT (does not infect the host), CONSISTENT (likely infects based on related taxa), or UNCLEAR (ambiguous due to conflicting descendant annotations). Taxa designated as "vertebrate-infecting" for inclusion in the VV database are those with MATCH or CONSISTENT status for the vertebrate host group (NCBI taxid 7742). Implementation details are available in the pipeline repository (github.com/securebio/nao-mgs-workflow).

For viral host composition analysis of Kraken2 results (Figure 3), we applied the same algorithm using an extended set of host categories grouped into six display categories: Vertebrate, Invertebrate (Metazoa minus Vertebrata), Bacteria (bacteriophages), Plant (Viridiplantae), Other (archaea, fungi, other eukaryotes, satellite viruses), and Unknown or ambiguous (metagenome-associated sequences, unclassified entries, and viruses not in the annotation database). Virus-Host DB has limited coverage of environmental bacteriophage lineages, likely resulting in underestimation of bacteriophage abundance.

For downstream analysis and figures (Figure 4a, Supplementary Figure S8), we excluded the Dicistroviridae family from vertebrate-infecting virus counts. Although two constituent taxa are annotated as vertebrate-infecting in Virus-Host DB, both appear to represent insect viruses detected in vertebrate feces rather than true vertebrate infections.

#### Benchmarking the vertebrate-infecting virus workflow

Sensitivity and specificity of the VV identification workflow were assessed on simulated reads from three panels of reference genomes. To assess sensitivity, we defined two "positive set" VV panels consisting of reference genomes from 93 notable VV taxa spanning 92 species, 27 families, and including DNA and RNA genomes (Supplementary Table S5-S6). Notable taxa were selected across published pathogen surveillance and priority lists, including from CDC NWSS and WastewaterSCAN targets, nationally notifiable conditions, the World Health Organization (WHO) 2024 pathogen prioritization (World Health Organization, une 2024), and vaccine-preventable viral diseases.

##### In-index positive set

The "in-index" positive set consists of 93 genomes selected from the VV genomes database used in the pipeline (index version 20250825). Each taxon is represented by one complete reference genome, selected as a whole RefSeq-linked assembly. Most taxa contribute a single accession, while segmented genomes (e.g., influenza and rotavirus) contribute one accession per segment, giving 165 accessions in total.

##### Out-of-index positive set

The "out-of-index" positive set consists of VV genomes from GenBank whose accessions were absent from the index. We selected VV genomes deposited on or after 11 March 2025, the day after the most recent sequence in the index 20250825 build (10 March 2025). Qualifying records required a complete genome or segment, excluding records described as unverified, patent, synthetic, vector, recombinant, chimeric, clone, partial, mutant, engineered, expression, plasmid, or nearly complete. Each species contributed one genome: a single record for a non-segmented species, and for a segmented species one complete isolate, with every segment drawn from that same isolate. This yielded 76 genomes across 76 species, comprising 134 accessions. Divergence of the out-of-index set from the VV database was assessed by aligning each accession to the masked VV genome database with minimap2 (v2.30; -x asm20 -c --secondary=no; Li, 2018). Of the 134 accessions, 122 aligned at 83.2% to 100% identity to the nearest database sequence (median 97.9%), while the remaining 12 were too divergent to produce a minimap2 alignment.

##### Negative set

To assess specificity, we defined a "negative set" measuring how often reads from common non-VV wastewater taxa were incorrectly assigned as VV. We used results from the Kraken2 shallow taxonomic profiling to select the 200 most abundant non-VV taxa, which together accounted for 22.7% of profiled reads. We retrieved one reference genome from RefSeq for each taxon, requiring complete assemblies. The final negative set, comprising 213 accessions representing 189 taxa and accounting for 19.9% of profiled reads, is dominated by bacteria (175 taxa) and includes human (GRCh38 assembly), bacteriophage (6 taxa), archaea (2 taxa), and tobamoviruses (5 taxa), including PMMoV and ToBRFV.

##### Simulating reads

We used InSilicoSeq (v2.0.1, Gourlé et al., 2019) to simulate 2 × 151 bp paired-end reads under its empirical NovaSeq error model, drawing fragment lengths from a normal distribution with mean 250 bp and standard deviation 50 bp. Each fragment’s start position was drawn uniformly at random along the source sequence, independently of every other fragment. For the positive sets, we generated 10,000 read pairs per genome, 930,000 pairs in-index and 760,000 out-of-index. For the negative set, we generated 500,000 read pairs per genome, 94,500,000 pairs in total. Genomes spanning several accessions divided their allocation among them in proportion to accession length.

##### Evaluation

Simulated reads were processed through the vertebrate-infecting virus identification workflow, and scored against the genome each read was simulated from. For the positive sets, the true and assigned taxa were projected onto species, genus and family in turn, and each read counted as correct at the finest rank where the two agreed. Reads matching at none of those ranks, and reads absent from the workflow’s output, were counted as incorrect. For the negative set, any read assigned to a vertebrate-infecting virus was a false positive. The rate of correct and incorrect assignments are shown in Supplementary Figure S7 and Supplementary Tables S5 and S6, with the total number of simulated reads in that set representing the denominator.

#### Relative abundance and normalization

We report VV abundances using two complementary metrics depending on the analysis context:

##### Total read relative abundance

For cross-site comparisons of overall VV abundance (Figure 5), we calculated total read relative abundance as VV read counts divided by total read pairs in the sequencing library.

##### Tobamovirus-normalized abundance

For pathogen time-series analysis (Figures 4c and 5), we normalized VV abundances from the VV pipeline by tobamovirus clade counts from the Kraken2 shallow taxonomic profiling. Because sequencing data is compositional, total read relative abundance depends on the abundance of all other taxa in the library, so shifts in background composition alter apparent pathogen abundance independent of pathogen concentration. Normalizing to a fecal indicator addresses this, and additionally accounts for variation in fecal input concentration, viral recovery efficiency, and sample processing across samples and sites. Tobamoviruses, including PMMoV and ToBRFV, are plant viruses shed in human feces at high and relatively stable concentrations. PMMoV in particular is widely used as a fecal strength indicator in wastewater surveillance (Maal-Bared et al., 2023; Symonds et al., 2018; Wolfe et al., 2021). Aggregating at the genus level yields higher counts than any single species and reduces sensitivity to variation in individual indicators, and improved concordance with wastewater PCR and clinical measurements in a retrospective evaluation of normalization approaches for WW-MGS data (Justen, Zulli, et al., 2026). Counts were taken as clade counts for the genus Tobamovirus in the non-rRNA fraction, and scaled to total library read depth (tobamovirus_scaled = tobamovirus_counts × total_read_pairs / n_reads_subsampled) to extrapolate from the subsampled profiling reads to the full library, where n_reads_subsampled is the total subsampled read pairs before quality filtering. Tobamovirus-normalized abundance was then calculated as VV_counts / tobamovirus_scaled.

#### Wastewater PCR data

We compared WW-MGS-derived pathogen abundances with wastewater PCR measurements obtained from the CDC NWSS data portal for sites where matching data was available. The NWSS portal aggregates wastewater PCR data from multiple sources, including state and local health departments, CDC, and WastewaterSCAN (Boehm et al., 2026) (Supplementary Table S10). Data was downloaded via the CDC Socrata API from the public datasets for SARS-CoV-2 (dataset j9g8-acpt; CDC, 2025c), Influenza A (dataset ymmh-divb; CDC, 2025a), and RSV (dataset 45cq-cw4i; CDC, 2025b). NWSS sewershed_id values were matched to CASPER sampling sites with assistance from sampling partners. Population served values reported in the NWSS data were used to inform population estimates in Table 1. NWSS sewershed IDs are shown in Supplementary Table S9. WastewaterSCAN data are shared under a CC BY-NC 4.0 license.

For Boston, NWSS maintains separate sewershed IDs for the DITP North (743) and South System (752); however, current NWSS data are only available for the combined system (sewershed 742), which we used for all Boston comparisons (Figure 5, Supplementary Figure S12-S13). For Boise, WW-MGS samples represent composites from two treatment plants (sewershed 372 and 374), each reporting separately to NWSS. We used data from sewershed 372 for comparison in Supplementary Figure S12, as it reported all three pathogens across the full comparison window.

For each matched site-pathogen combination we used the raw pathogen concentration reported to NWSS as the comparison metric. Concentrations are reported in each source’s native units, with copies per litre of wastewater for sites reporting through state and local health departments and copies per gram of dry sludge for WastewaterSCAN sites. Because comparisons use Spearman correlation, which is invariant to monotonic rescaling, differences in reporting units do not affect the reported coefficients. WW-MGS abundances were tobamovirus-normalized for all wastewater PCR comparisons. Wastewater PCR assays are further detailed in Supplementary Table S10.

#### Clinical respiratory virus diagnostic testing data

We compared WW-MGS-derived, tobamovirus-normalized pathogen abundances from Boston DITP with clinical respiratory virus testing data from the Massachusetts General Hospital and its affiliated outpatient health centers. Clinical testing data comprised aggregated weekly test counts (positive and negative) from nucleic acid amplification test (NAAT) assays performed by the Massachusetts General Hospital Clinical Microbiology Laboratory, covering the period from the start of Boston wastewater sequencing through August 31, 2025. The hospital network’s catchment overlaps geographically with the sewershed served by Boston DITP.

Clinical respiratory virus testing followed a seasonal algorithm in emergency department (ED) and inpatient (IP) settings: during fall and winter months, symptomatic patients received multiplex molecular testing that included at least influenza, SARS-CoV-2, and RSV; during spring and summer months, SARS-CoV-2-only testing was the default, with multiplex molecular testing reserved for special populations (immunocompromised and pediatric patients being admitted to the hospital). In outpatient (OP) settings, a different order set was used where test selection was at clinician discretion, with embedded language within the order set describing appropriate indications for testing. Testing for SARS-CoV-2, influenza A/B, and RSV was available using this order set; testing for additional respiratory viruses required microbiology laboratory approval. All available multiplex molecular assays across all settings were either cleared or authorized for emergency use by the Food and Drug Administration and had been verified for clinical use by the clinical laboratory.

For our comparison, we applied test filtering that mirrored this seasonal testing algorithm. During fall and winter months, we used positive counts from multiplex molecular assays only, excluding SARS-CoV-2-specific assays to avoid repeat testing of known positives. During spring and summer months, we included SARS-CoV-2-specific assays, as this was the default testing strategy for symptomatic patients in ED and IP settings and the recommended approach in OP settings. Influenza and RSV counts used multiplex molecular assays only year-round, as no single-pathogen assays exist for these pathogens. Test volume and positive rate by pathogen are shown in Supplementary Figure S13.

Clinical data was collected under a waiver of consent as non-identifiable aggregate variables with oversight from Massachusetts General Brigham Institutional Review Board (IRB: 2023P001716).

#### Norovirus and SARS-CoV-2 variant assignment and comparison

To assess whether CASPER data support strain-level genomic analysis, we examined norovirus genotype and SARS-CoV-2 lineage composition and compared both with national clinical genomic surveillance.

For SARS-CoV-2, we extracted read pairs assigned to the SARS-CoV-2 clade by the VV pipeline and aligned them to the Wuhan-Hu-1 reference genome (NC_045512.2) with Bowtie2 (parameters: --local --very-sensitive-local). Reads were pooled across all sites within each calendar month, and lineage abundances were estimated with Freyja (v2.0.0; UShER barcodes downloaded 2026-09-01; Levy et al., 2025; Turakhia et al., 2021). For comparison with national clinical data, SARS-CoV-2 variant proportions were obtained from the CDC SARS-CoV-2 Variant Proportions dataset (CDC, 2021, dataset jr58-6ysp), using the national series of four-week empiric estimates.

For norovirus, we extracted read pairs assigned to the Norwalk virus clade by the VV pipeline and aligned them to reference sequences from the CDC Human Calicivirus Typing Tool (HuCaT) (Tatusov et al., 2021), the reference set used by CaliciNet to type outbreak specimens. References were filtered to near-complete VP1 (≥1,400 bp), giving 228 references across 37 genotypes. Reads were aligned to the whole panel simultaneously with Bowtie2 (parameters: --local --very-sensitive-local -k 10 --no-unal), and each read pair was assigned to the genotype whose best-scoring reference exceeded every other genotype by at least 10 alignment-score points. Pairs without a clear winner were counted as ambiguous (1.6% of aligned pairs) and excluded from reported fractions. For comparison, norovirus genotype proportions were taken from monthly CaliciNet outbreak percentages, reported as GII.4 and GII.17 proportions against a total-outbreak denominator (Barclay & Vinjé, 2025), so remaining genotypes are reported as a single unresolved category and are not directly comparable with the measured other-genotype fraction in the wastewater data.

#### Time series smoothing and correlation

To enable comparison across data sources with different sampling frequencies, all smoothed time series plots in this manuscript use a consistent methodology. WW-MGS and wastewater PCR time series were aggregated to MMWR (Morbidity and Mortality Weekly Report) epidemiological weeks using the epiweeks Python package (v2.3.0). Clinical data was already provided as weekly aggregates aligned to MMWR weeks (reported by week start date, Sunday).

Values within MMWR weeks were averaged using arithmetic mean for clinical data and geometric mean for WW-PCR and WW-MGS. We then applied a 5-week moving average, with the geometric mean computed as expm1(mean(log1p(x))) to remain defined for zero-value observations in the window. For visualization, smoothed values were plotted at the MMWR week midpoint (Wednesday).

For time series visualizations, site-specific filtering was applied to avoid visual artifacts from sparse early sampling: the first sample from NYC Hospital B and each MU-processed Boston DITP site was dropped, and NYC Hospital C excludes all samples before October 2025. NYC Hospitals A and D are plotted without filtering. This filtering applies to time series visualisations (Figures 3e, 4c, and 5; Supplementary Figures S9–S13) but not to summary statistics, per-site supplementary plots, or correlation calculations.

Spearman correlation coefficients were calculated between MMWR-week-aligned smoothed time series for each site-pathogen and wastewater PCR versus clinical combination, restricted to the overlapping date range where both data sources had measurements. Spearman correlation was used because the relationship between WW-MGS and wastewater PCR and clinical measurements is expected to be monotonic but not necessarily linear. All statistical tests reported in this study are two-sided.

#### Comparison with public SRA wastewater sequencing data

To examine the CASPER dataset as a fraction of publicly available wastewater sequencing data (Figure 2d, Supplementary Figure S2a), we queried the NCBI SRA metadata using Google BigQuery (table ‘nih-sra-datastore.sra.metadatà). We defined two reference categories for comparison:

1. **All wastewater metagenomic sequencing** (Figure 2d): SRA runs with ‘organism’ IN ("wastewater metagenome", “sludge metagenome”), representing publicly deposited wastewater metagenomic sequencing regardless of sequencing methodology.

2. **Untargeted wastewater sequencing** (Supplementary Figure S2a): A subset of wastewater and sludge metagenome runs filtered to ‘assay_typè IN ("RNA-Seq", "WGS") and ‘librarysourcè IN ("METAGENOMIC", "METATRANSCRIPTOMIC"), representing untargeted shotgun or metatranscriptomic sequencing approaches comparable to the CASPER methodology.

Both filters require ‘organism’ IN ("wastewater metagenome", “sludge metagenome”) and therefore do not capture studies deposited under other organism labels (e.g., "viral metagenome", "metagenomes").

For CASPER data, we queried runs associated with BioProject PRJNA1247874 (this data release) and PRJNA1198001 (the previously published Los Angeles dataset from Grimm, Rothman, et al., 2025). Monthly sequencing volumes were aggregated by sample collection date for each category. SRA metadata queries were performed on 2026-03-31.

To compare CASPER data with prior WW-MGS efforts (Figure 2e, Supplementary Figure S2b), we compiled a non-systematic collection of published studies. For each study with publicly deposited data, we queried the associated NCBI SRA BioProject for sample-level base pair statistics. We reviewed each SRA record and classified samples as untargeted metagenomic sequencing, excluding amplicon or hybrid-capture samples when computing per-sample median sequencing depth and total sequencing effort. For studies without public SRA data, we estimated these metrics from statistics reported in the manuscript text, tables, or supplementary materials, excluding studies where such estimation was not tractable. The full annotated literature review table is provided in Supplementary Table S1.

## Ethical and privacy considerations

Wastewater sequencing captures human nucleic acid alongside microbial sequence. We perform no analysis of human sequence, and human-derived reads are masked using NCBI’s Human Read Removal Tool (Katz et al., 2021) prior to public release.

## Data availability

Sequencing data generated by the CASPER consortium included in this release are available on the NCBI SRA under BioProject PRJNA1247874. Prior to public release, human-derived reads were masked using NCBI’s Human Read Removal Tool. Additional deep sequencing data from Los Angeles, collected under CASPER, are available under BioProject PRJNA1198001 (Grimm, Rothman, et al., 2025).

Near real-time visualization of human-infecting virus trends from CASPER sampling sites is available through public dashboards maintained by the University of Wisconsin-Madison (O’Connor et al., 2025).

Wastewater PCR data used for validation are publicly available through the CDC NWSS data portal: SARS-CoV-2 (dataset j9g8-acpt), Influenza A (dataset ymmh-divb), and RSV (dataset 45cq-cw4i). A subset of these data were generated by WastewaterSCAN (Boehm et al. 2024, 2026) and are shared under a CC BY-NC 4.0 license; see https://data.wastewaterscan.org/about for terms of use. WastewaterSCAN data were collected as part of the WastewaterSCAN / SCAN project, a partnership between Stanford University, Emory University, and Verily funded philanthropically through a gift to Stanford University.

Tabular data underlying manuscript figures, PMMoV RT-qPCR measurements, and clinical respiratory virus testing data from Massachusetts General Hospital are available in the paper’s GitHub repository (github.com/securebio/casper-paper).

## Acknowledgements

We thank the University of Missouri Genomics Technology Core for sequencing services, technical support in weekly sequencing efforts, and optimization of library preparation protocols. We thank the Broad Clinical Labs at the Broad Institute for sequencing support. We thank the staff of the Riverside Water Quality Control Plant; Gabriel Holguin and Rodney Brees at Inland Empire Utilities Agency Regional Water Recycling Plant No. 1; Steve Rhode and the Massachusetts Water Resources Authority Central Laboratory Violet Team; the operations staff at NYC Health + Hospitals; the Sacramento Area Sewer District and its Environmental Laboratory; and Tami Hansen at the Columbia, MO, wastewater treatment plant for assistance with sample collection. We thank Adam Horton and Dolores Sanchez Gonzalez for coordinating sample shipments from Chicago. We thank the National Center for Biotechnology Information (NCBI) team at the National Library of Medicine (NLM) for their support with SRA metadata management. Figure 1b was created with BioRender (https://BioRender.com/1o0o1uk).

Prepared by Lawrence Livermore National Laboratory under Contract DE-AC52-07NA27344.

The conclusions, opinions, or recommendations in this paper are those of the authors and not of the Illinois Department of Public Health, the Chicago Department of Public Health, or the Centers for Disease Control and Prevention.

## Author contributions

**Conceptualization:** Lennart Justen, Clayton Rushford, Jeff Kaufman, Marc Johnson, Rose Kantor. **Methodology:** Lennart Justen, Clayton Rushford, Olivia Hershey, William Bradshaw, Mike McLaren, Jeff Kaufman, Marc Johnson. **Software:** Lennart Justen, Simon Grimm, William Bradshaw, Harmon Bhasin, Daniel Rice, Katherine Stansifer, Jo Faraguna. **Validation:** Lennart Justen, Clayton Rushford, William Bradshaw, Harmon Bhasin. **Formal analysis:** Lennart Justen. **Investigation:** Lennart Justen, Clayton Rushford, Olivia Hershey, Róisín Floyd-O’Sullivan, Alejandro Tovar-Mendez, Emma Copen. **Resources:** Lennart Justen, Kristen Shelton, Ayaaz Amirali, Sherin Kannoly, Sofia Pesantez, Aiden Stanciu, Iñigo Caballero Quiroga, Leopolda Silvera, Nicole Greenwood, Barbra Bongiovi, Austin Walkins, Ryan Love, Scott Lening, Kaylyn Patterson, Theresa Johnston, Sandra Hernandez, Aymara Benitez, Billie Jo McCarley, Samantha Engelage, Suguna Pillay, Cindy Calender, Brent Herring, Carey Robinson, Monett Wastewater Treatment Plant, Columbia Missouri Wastewater Treatment Plant, Gordon Adams, Jillian Paull, Jamie Devlin, Vamsi Thiriveedhi, Sarah Turbett, Jacob E. Lemieux, John Dennehy, Rachel Poretsky, Jason Rothman, Helena Solo-Gabriele, Jason Vogel, Pardis Sabeti, Jeff Kaufman, Marc Johnson. **Data curation:** Lennart Justen, Clayton Rushford, Róisín Floyd-O’Sullivan, Simon Grimm, Daniel Rice, Katherine Stansifer, Jo Faraguna, Alessandro Zulli, Alejandro Tovar-Mendez, Emma Copen, Aiden Stanciu, Gordon Adams, Jillian Paull, Jamie Devlin, Vamsi Thiriveedhi, Sarah Turbett, John Dennehy, Rachel Poretsky, Jason Rothman, Helena Solo-Gabriele, Jason Vogel. **Writing – original draft:** Lennart Justen, Clayton Rushford, Olivia Hershey. **Writing – review & editing:** Lennart Justen, Clayton Rushford, William Bradshaw, Katherine Stansifer, Michael McLaren, Alessandro Zulli, Daniel Cunningham-Bryant, Gordon Adams, Jillian Paull, Sarah Turbett, Rose Kantor, David O’Connor, John Dennehy, Jason Rothman, Helena Solo-Gabriele, Pardis Sabeti, Jeff Kaufman, Marc Johnson. **Visualization:** Lennart Justen. **Supervision:** Olivia Hershey, William Bradshaw, Daniel Cunningham-Bryant, Sarah Turbett, Jacob E. Lemieux, John Dennehy, Rachel Poretsky, Helena Solo-Gabriele, Pardis Sabeti, Jeff Kaufman, Marc Johnson. **Project administration:** Lennart Justen, Clayton Rushford, Jeff Kaufman, Marc Johnson. **Funding acquisition:** David O’Connor, Jason Rothman, Helena Solo-Gabriele, Pardis Sabeti, Jeff Kaufman, Marc Johnson.

## Code availability

The SecureBio Viral Metagenomics Pipeline used to process sequencing data is available at github.com/securebio/nao-mgs-workflow. Code for generating figures and analyses presented in this paper is available at github.com/securebio/casper-paper.

## References

Adalja, A. A., Watson, M., Toner, E. S., Cicero, A., & Inglesby, T. V. (2018). The Characteristics of Pandemic Pathogens. Center for Health Security, Johns Hopkins Bloomberg School of Health. https://www.centerforhealthsecurity.org/our-work/pubs_archive/pubs-pdfs/2018/180510-pandemic-pathogens-report.pdf

Adams, C., Bias, M., Welsh, R. M., Webb, J., Reese, H., Delgado, S., Person, J., West, R., Shin, S., & Kirby, A. (2024). The National Wastewater Surveillance System (NWSS): From inception to widespread coverage, 2020-2022, United States. The Science of the Total Environment, 924(171566), 171566. 10.1016/j.scitotenv.2024.171566

Adams, C., Kirby, A. E., Bias, M., Riser, A., Wong, K. K., Mercante, J. W., & Reese, H. (2024). Detecting mpox cases through wastewater surveillance - United States, august 2022-may 2023. MMWR. Morbidity and Mortality Weekly Report, 73(2), 37–43. 10.15585/mmwr.mm7302a3

Ameaka, F., Cardo, D., Jernigan, D., Rivers, C., & Kelly, A. (2026). Why Metagenomic Surveillance Requires Investment in Decision-making Support, Not Just Technology. Shield Initiative. https://shield.centerforhealthsecurity.org/sites/default/files/2026-05/MetaBridge.pdf

Andrews, S. (2010). FastQC: A Quality Control Tool for High Throughput Sequence Data. https://github.com/s-andrews/FastQC

Balaji, A., Kille, B., Kappell, A. D., Godbold, G. D., Diep, M., Elworth, R. A. L., Qian, Z., Albin, D., Nasko, D. J., Shah, N., Pop, M., Segarra, S., Ternus, K. L., & Treangen, T. J. (2022). SeqScreen: accurate and sensitive functional screening of pathogenic sequences via ensemble learning. Genome Biology, 23(1), 133. 10.1186/s13059-022-02695-x

Barclay, L., & Vinjé, J. (2025). Increasing predominance of Norovirus GII.17 over GII.4, United States, 2022-2025. Emerging Infectious Diseases, 31(7), 1471–1473. 10.3201/eid3107.250524

Blin, K. (2023). ncbi-genome-download. Zenodo. 10.5281/ZENODO.8192486

Boehm, A. B., Wolfe, M. K., Bidwell, A. L., Zulli, A., Chan-Herur, V., White, B. J., Shelden, B., & Duong, D. (2024). Human pathogen nucleic acids in wastewater solids from 191 wastewater treatment plants in the United States. Scientific Data, 11(1), 1141. 10.1038/s41597-024-03969-8

Boehm, A. B., Wolfe, M. K., Bidwell, A. L., Zulli, A., White, B. J., Shelden, B., & Duong, D. (2026). Pathogen nucleic acids data in wastewater solids from 147 treatment plants in the United States: 2024-2025. Data in Brief, 65(112503), 112503. 10.1016/j.dib.2026.112503

Bradshaw, W. J., Bhasin, H., Grimm, S. L., Stansifer, K., Rice, D. P., Kaufman, J., Fields, E., Justen, L., McLaren, M. R., & Faraguna, J. D. (2026). securebio/nao-mgs-workflow: 3.0.1.9. Zenodo. 10.5281/ZENODO.18434553

Bushnell, B. (2014, March 17). BBMap: A Fast, Accurate, Splice-Aware Aligner. 9th Annual Genomics of Energy & Environment Meeting, Walnut Creek, CA. https://www.osti.gov/servlets/purl/1241166

Camargo, A. P., Roux, S., Schulz, F., Babinski, M., Xu, Y., Hu, B., Chain, P. S. G., Nayfach, S., & Kyrpides, N. C. (2024). Identification of mobile genetic elements with geNomad. Nature Biotechnology, 42(8), 1303–1312. 10.1038/s41587-023-01953-y

CDC. (2021). SARS-CoV-2 Variant Proportions [Dataset]. https://data.cdc.gov/Laboratory-Surveillance/SARS-CoV-2-Variant-Proportions/jr58-6ysp

CDC. (2025a). CDC Wastewater Data for Influenza A [Dataset]. https://data.cdc.gov/Public-Health-Surveillance/CDC-Wastewater-Data-for-Influenza-A/ymmh-divb

CDC. (2025b). CDC Wastewater Data for RSV [Dataset]. https://data.cdc.gov/Public-Health-Surveillance/CDC-Wastewater-Data-for-RSV/45cq-cw4i

CDC. (2025c). CDC Wastewater Data for SARS-CoV-2 [Dataset]. https://data.cdc.gov/Public-Health-Surveillance/CDC-Wastewater-Data-for-SARS-CoV-2/j9g8-acpt

Chen, S., Zhou, Y., Chen, Y., & Gu, J. (2018). fastp: an ultra-fast all-in-one FASTQ preprocessor. Bioinformatics (Oxford, England), 34(17), i884–i890. 10.1093/bioinformatics/bty560

Chuvochina, M., Gerken, J., Frentrup, M., Sandikci, Y., Goldmann, R., Freese, H. M., Göker, M., Sikorski, J., Yarza, P., Quast, C., Peplies, J., Glöckner, F. O., & Reimer, L. C. (2025). SILVA in 2026: a global core biodata resource for rRNA within the DSMZ digital diversity. Nucleic Acids Research, gkaf1247, gkaf1247. 10.1093/nar/gkaf1247

Committee on Community Wastewater-based Infectious Disease Surveillance, Water Science and Technology Board, Division on Earth and Life Studies, Board on Population Health and Public Health Practice, Health and Medicine Division, & National Academies of Sciences, Engineering, and Medicine. (2024). Increasing the utility of wastewater-based disease surveillance for public health action: A phase 2 report. National Academies Press. 10.17226/27516

Concentrating Pathogens from Raw and Primary Wastewater Using the InnovaPrep® Concentrating Pipette, Protocol (Revision E). (n.d.). InnovaPrep.

Corrin, T., Rabeenthira, P., Young, K. M., Mathiyalagan, G., Baumeister, A., Pussegoda, K., & Waddell, L. A. (2024). A scoping review of human pathogens detected in untreated human wastewater and sludge. Journal of Water and Health, 22(2), 436–449. 10.2166/wh.2024.326

Crook, O. M., Warmbrod, K. L., Lipstein, G., Chung, C., Bakerlee, C. W., McKelvey, T. G., Jr, Holland, S. R., Swett, J. L., Esvelt, K. M., Alley, E. C., & Bradshaw, W. J. (2022). Analysis of the first genetic engineering attribution challenge. Nature Communications, 13(1), 7374. 10.1038/s41467-022-35032-8

Diamond, M. B., Keshaviah, A., Bento, A. I., Conroy-Ben, O., Driver, E. M., Ensor, K. B., Halden, R. U., Hopkins, L. P., Kuhn, K. G., Moe, C. L., Rouchka, E. C., Smith, T., Stevenson, B. S., Susswein, Z., Vogel, J. R., Wolfe, M. K., Stadler, L. B., & Scarpino, S. V. (2022). Wastewater surveillance of pathogens can inform public health responses. Nature Medicine, 28(10), 1992–1995. 10.1038/s41591-022-01940-x

Di Tommaso, P., Chatzou, M., Floden, E. W., Barja, P. P., Palumbo, E., & Notredame, C. (2017). Nextflow enables reproducible computational workflows. Nature Biotechnology, 35(4), 316–319. 10.1038/nbt.3820

Duvallet, C., Wu, F., McElroy, K. A., Imakaev, M., Endo, N., Xiao, A., Zhang, J., Floyd-O’Sullivan, R., Powell, M. M., Mendola, S., Wilson, S. T., Cruz, F., Melman, T., Sathyanarayana, C. L., Olesen, S. W., Erickson, T. B., Ghaeli, N., Chai, P., Alm, E. J., & Matus, M. (2022). Nationwide trends in COVID-19 cases and SARS-CoV-2 RNA wastewater concentrations in the United States. ACS ES&T Water, 2(11), 1899–1909. 10.1021/acsestwater.1c00434

Edgar, R. C., Taylor, B., Lin, V., Altman, T., Barbera, P., Meleshko, D., Lohr, D., Novakovsky, G., Buchfink, B., Al-Shayeb, B., Banfield, J. F., de la Peña, M., Korobeynikov, A., Chikhi, R., & Babaian, A. (2022). Petabase-scale sequence alignment catalyses viral discovery. Nature, 602(7895), 142–147. 10.1038/s41586-021-04332-2

Ewels, P., Magnusson, M., Lundin, S., & Käller, M. (2016). MultiQC: summarize analysis results for multiple tools and samples in a single report. Bioinformatics (Oxford, England), 32(19), 3047–3048. 10.1093/bioinformatics/btw354

Fiamenghi, M. B., Camargo, A. P., Chasapi, I. N., Baltoumas, F. A., Roux, S., Egorov, A. A., Aplakidou, E., Ndela, E. O., Vasquez, Y. M., Chen, I.-M. A., Palaniappan, K., Reddy, T. B. K., Mukherjee, S., Ivanova, N. N., Schulz, F., Woyke, T., Eloe-Fadrosh, E. A., Pavlopoulos, G. A., & Kyrpides, N. C. (2025). Meta-virus resource (MetaVR): expanding the frontiers of viral diversity with 24 million uncultivated virus genomes. Nucleic Acids Research, gkaf1283, gkaf1283. 10.1093/nar/gkaf1283

Friedman, C. R., Morfino, R. C., & Ernst, E. T. (2025). Leveraging a strategic public-private partnership to launch an airport-based pathogen monitoring program to detect emerging health threats. Emerging Infectious Diseases, 31(13), 35–38. 10.3201/eid3113.241407

Gourlé, H., Karlsson-Lindsjö, O., Hayer, J., & Bongcam-Rudloff, E. (2019). Simulating Illumina metagenomic data with InSilicoSeq. Bioinformatics (Oxford, England), 35(3), 521–522. 10.1093/bioinformatics/bty630

Grassly, N. C., Shaw, A. G., & Owusu, M. (2025). Global wastewater surveillance for pathogens with pandemic potential: opportunities and challenges. The Lancet. Microbe, 6(1), 100939. 10.1016/j.lanmic.2024.07.002

Grimm, S. L., Kaufman, J. T., Rice, D. P., Whittaker, C., Bradshaw, W. J., & McLaren, M. R. (2025). Inferring the sensitivity of wastewater metagenomic sequencing for early detection of viruses: a statistical modelling study. The Lancet. Microbe, 101187. 10.1016/j.lanmic.2025.101187

Grimm, S. L., Rothman, J. A., Bradshaw, W. J., Langlois, K., Steele, J. A., Griffith, J. F., Kaufman, J. T., & Whiteson, K. L. (2025). Deep metatranscriptomic sequencing data of wastewater from Los Angeles, USA, 2023-2024. Scientific Data, 13(1), 158. 10.1038/s41597-025-06475-7

Hellmér, M., Paxéus, N., Magnius, L., Enache, L., Arnholm, B., Johansson, A., Bergström, T., & Norder, H. (2014). Detection of pathogenic viruses in sewage provided early warnings of hepatitis A virus and norovirus outbreaks. Applied and Environmental Microbiology, 80(21), 6771–6781. 10.1128/AEM.01981-14

Justen, L. J., Bhasin, H., Whittaker, C., Cunningham-Bryant, D., Lanteri, M. C., Busch, M. P., Esvelt, K. M., & Sabeti, P. C. (2026). Leveraging the U.S. blood supply to detect emerging viral threats. medRxiv, 2026.02.06.26345722. 10.64898/2026.02.06.26345722

Justen, L. J., Grimm, S. L., Esvelt, K. M., & Bradshaw, W. J. (2025). Indoor air sampling for detection of viral nucleic acids. Journal of Aerosol Science, 106549, 106549. 10.1016/j.jaerosci.2025.106549

Justen, L. J., Zulli, A., Kantor, R. S., Linfield, R. Y., Moskatel, L. S., Cunningham-Bryant, D., Kaufman, J., Johnson, M. C., McLaren, M. R., & Sabeti, P. (2026). Establishing wastewater metagenomics as a quantitative pathogen monitoring tool with normalization. In medRxiv (p. 2026.07.14.26356442). medRxiv. 10.64898/2026.07.14.26356442

Kantor, R. S., & Jiang, M. (2024). Considerations and opportunities for probe capture enrichment sequencing of emerging viruses from wastewater. Environmental Science & Technology, 58(19), 8161–8168. 10.1021/acs.est.4c02638

Kantor, R. S., Shakya, M., Ruth, N., Rothman, J. A., Rushford, C., Gregory, D. A., Epstein, A., Kaufman, J. T., Allen, J. E., Chain, P. S. G., O’Connor, D. H., & Johnson, M. C. (2026). A genome-resolved view of the wastewater RNA virome. In medRxiv (p. 2026.05.19.26353600). medRxiv. 10.64898/2026.05.19.26353600

Karthikeyan, S., Levy, J. I., De Hoff, P., Humphrey, G., Birmingham, A., Jepsen, K., Farmer, S., Tubb, H. M., Valles, T., Tribelhorn, C. E., Tsai, R., Aigner, S., Sathe, S., Moshiri, N., Henson, B., Mark, A. M., Hakim, A., Baer, N. A., Barber, T.,… Knight, R. (2022). Wastewater sequencing reveals early cryptic SARS-CoV-2 variant transmission. Nature, 609(7925), 101–108. 10.1038/s41586-022-05049-6

Katz, K. S., Shutov, O., Lapoint, R., Kimelman, M., Brister, J. R., & O’Sullivan, C. (2021). STAT: a fast, scalable, MinHash-based k-mer tool to assess Sequence Read Archive next-generation sequence submissions. Genome Biology, 22(1), 270. 10.1186/s13059-021-02490-0

Kaufman, J. (2024, June 27). Detecting Genetically Engineered Viruses With Metagenomic Sequencing. https://securebio.org/blog/detecting-genetically-engineered-viruses/

Kaufman, J. (2025, June 25). How Much Data From a Sequencing Run? SecureBio. https://data.securebio.org/jefftk-notebook/how-much-data-from-a-sequencing-run

Kean, K., Mayne, R. M., Reid, K., Secret, S., Singleton, B. K., Rockett, R., Rajendra, P., Harvala, H., Breuer, J., Ansari, M. A., Lythgoe, K., Simmonds, P., & Golubchik, T. (2026). A snapshot of the UK blood donor plasma virome: a retrospective cross-sectional cohort study. In medRxiv (p. 2026.05.20.26353651). medRxiv. 10.64898/2026.05.20.26353651

Kidd, M., Richter, A., Best, A., Cumley, N., Mirza, J., Percival, B., Mayhew, M., Megram, O., Ashford, F., White, T., Moles-Garcia, E., Crawford, L., Bosworth, A., Atabani, S. F., Plant, T., & McNally, A. (2021). S-variant SARS-CoV-2 lineage B1.1.7 is associated with significantly higher viral load in samples tested by TaqPath polymerase chain reaction. The Journal of Infectious Diseases, 223(10), 1666–1670. 10.1093/infdis/jiab082

Kirby, A. E., Welsh, R. M., Marsh, Z. A., Yu, A. T., Vugia, D. J., Boehm, A. B., Wolfe, M. K., White, B. J., Matzinger, S. R., Wheeler, A., Bankers, L., Andresen, K., Salatas, C., New York City Department of Environmental Protection, Gregory, D. A., Johnson, M. C., Trujillo, M., Kannoly, S., Smyth, D. S.,… Hopkins, L. (2022). Notes from the field: Early evidence of the SARS-CoV-2 B.1.1.529 (omicron) variant in community wastewater - United States, November-December 2021. MMWR. Morbidity and Mortality Weekly Report, 71(3), 103–105. 10.15585/mmwr.mm7103a5

Lambisia, A. W., Mohammed, K. S., Makori, T. O., Ndwiga, L., Mburu, M. W., Morobe, J. M., Moraa, E. O., Musyoki, J., Murunga, N., Mwangi, J. N., Nokes, D. J., Agoti, C. N., Ochola-Oyier, L. I., & Githinji, G. (2022). Optimization of the SARS-CoV-2 ARTIC Network V4 primers and whole genome sequencing protocol. Frontiers in Medicine, 9, 836728. 10.3389/fmed.2022.836728

Langmead, B., & Salzberg, S. L. (2012). Fast gapped-read alignment with Bowtie 2. Nature Methods, 9(4), 357–359. 10.1038/nmeth.1923

Levy, J. I., Gangavarapu, P., Pilz, D. A., Jesvaghane, M. A., Steedman, A., Zeller, M., Gangavarapu, K., McDonald, D., Libuit, K., Kapsak, C., Young, E. L., Agrawal, S., Orschler, L., Hölzer, M., Stachler, E., Park, D. J., Hinrichs, A. S., Laurent, L. C., Knight, R., & Andersen, K. G. (2025). Real-time, multi-pathogen wastewater genomic surveillance with Freyja 2. In medRxiv (p. 2025.07.26.25332245). 10.1101/2025.07.26.25332245

Li, H. (2013). seqtk. https://github.com/lh3/seqtk

Li, H. (2018). Minimap2: pairwise alignment for nucleotide sequences. Bioinformatics (Oxford, England), 34(18), 3094–3100. 10.1093/bioinformatics/bty191

Liu, O., Jaghouar, S., Hagemann, J., Wang, S., Wiemels, J., Kaufman, J., & Neiswanger, W. (2025). METAGENE-1: Metagenomic foundation model for pandemic monitoring. In arXiv [q-bio.GN]. arXiv. http://arxiv.org/abs/2501.02045

Luebbert, L., Sullivan, D. K., Carilli, M., Eldjárn Hjörleifsson, K., Viloria Winnett, A., Chari, T., & Pachter, L. (2025). Detection of viral sequences at single-cell resolution identifies novel viruses associated with host gene expression changes. Nature Biotechnology. 10.1038/s41587-025-02614-y

Maal-Bared, R., Qiu, Y., Li, Q., Gao, T., Hrudey, S. E., Bhavanam, S., Ruecker, N. J., Ellehoj, E., Lee, B. E., & Pang, X. (2023). Does normalization of SARS-CoV-2 concentrations by Pepper Mild Mottle Virus improve correlations and lead time between wastewater surveillance and clinical data in Alberta (Canada): comparing twelve SARS-CoV-2 normalization approaches. The Science of the Total Environment, 856(Pt 1), 158964. 10.1016/j.scitotenv.2022.158964

Machtinger, A. N., Hershey, O. S., Bradshaw, W. J., Rice, D. P., & McLaren, M. R. (2025, May 26). Concentration and nucleic acid extraction of viruses from wastewater influent. Protocols.io. https://www.protocols.io/view/concentration-and-nucleic-acid-extraction-of-virus-gzzhbx737

McLaren, M. R., Hershey, O. S., Machtinger, A. N., Rice, D. P., Simas, A. M., Friedman, C. R., Gratalo, D., Philipson, C. W., & Bradshaw, W. J. (2026). Metagenomic sequencing of composite airplane wastewater for surveillance of emerging viruses. In medRxiv (p. 2026.01.29.26343714). medRxiv. 10.64898/2026.01.29.26343714

Mihara, T., Nishimura, Y., Shimizu, Y., Nishiyama, H., Yoshikawa, G., Uehara, H., Hingamp, P., Goto, S., & Ogata, H. (2016). Linking virus genomes with host taxonomy. Viruses, 8(3), 66. 10.3390/v8030066

Minor, N. R., Ramuta, M. D., Stauss, M. R., Harwood, O. E., Brakefield, S. F., Alberts, A., Vuyk, W. C., Bobholz, M. J., Rosinski, J. R., Wolf, S., Lund, M., Mussa, M., Beversdorf, L. J., Aliota, M. T., O’Connor, S. L., & O’Connor, D. H. (2023). Metagenomic sequencing detects human respiratory and enteric viruses in air samples collected from congregate settings. Scientific Reports, 13(1), 21398. 10.1038/s41598-023-48352-6

Morfino, R. C., Bart, S. M., Franklin, A., Rome, B. H., Rothstein, A. P., Aichele, T. W. S., Li, S. L., Bivins, A., Ernst, E. T., & Friedman, C. R. (2023). Notes from the Field: Aircraft Wastewater Surveillance for Early Detection of SARS-CoV-2 Variants - John F. Kennedy International Airport, New York City, August-September 2022. MMWR. Morbidity and Mortality Weekly Report, 72(8), 210–211. 10.15585/mmwr.mm7208a3

Neri, U., Wolf, Y. I., Roux, S., Camargo, A. P., Lee, B., Kazlauskas, D., Chen, I. M., Ivanova, N., Zeigler Allen, L., Paez-Espino, D., Bryant, D. A., Bhaya, D., RNA Virus Discovery Consortium, Krupovic, M., Dolja, V. V., Kyrpides, N. C., Koonin, E. V., & Gophna, U. (2022). Expansion of the global RNA virome reveals diverse clades of bacteriophages. Cell, 185(21), 4023–4037.e18. 10.1016/j.cell.2022.08.023

O’Connor, D., Minor, N., Gardner, W., & Johnson, M. (2025). Wastewater Surveillance Dashboards. https://dholab.github.io/public_viz/001-make-by-city-and-by-virus-dashboards/index.html

Peccia, J., Zulli, A., Brackney, D. E., Grubaugh, N. D., Kaplan, E. H., Casanovas-Massana, A., Ko, A. I., Malik, A. A., Wang, D., Wang, M., Warren, J. L., Weinberger, D. M., Arnold, W., & Omer, S. B. (2020). Measurement of SARS-CoV-2 RNA in wastewater tracks community infection dynamics. Nature Biotechnology, 38(10), 1164–1167. 10.1038/s41587-020-0684-z

Poretsky, R. S., Dhiman, V. K., Hendricks, D. L., Lin, C.-Y., Sanchez Gonzalez, D., Greenwald, S., Owens, S. M., Williams, C. H., Leslie, M. T., Bemis, K., Frias, M., Kaufman, J. T., O’Connor, D. H., & Johnson, M. C. (2026). Detection of a single measles infection using untargeted ultra-deep metagenomic sequencing of wastewater in Cook County, Illinois. NEJM Evidence, EVIDpha2600079. 10.1056/EVIDpha2600079

Pray, I., Roguet, A., Dutcher, H. A., Schauer, S., Deraedt, M., Rasmussen, M., Westergaard, R., Antkiewicz, D., Henning, E., Everett, D., & Shafer, M. (2026). Genotype-specific detection of measles virus using wastewater surveillance - Wisconsin, February 2026. MMWR. Morbidity and Mortality Weekly Report, 75(27), 344–348. 10.15585/mmwr.mm7527a1

Public Health England. (2020). Investigation of novel SARS-COV-2 variant: Variant of Concern 202012/01. https://assets.publishing.service.gov.uk/government/uploads/system/uploads/attachment_data/file/959438/Technical_Briefing_VOC_SH_NJL2_SH2.pdf

Rebecca Falender, D., Melissa Sutton, M., Cieslak, P., Liko, J., Mickle, D., Kelly, C., & Radniecki, T. (2026). Notes from the field: Retrospective analysis of wild-type measles virus in wastewater during a measles outbreak — Oregon, march 24–September 22, 2024. MMWR. Morbidity and Mortality Weekly Report, 75. 10.15585/mmwr.mm7502a1

Renfro, Z. T., Zulli, A., Parsonnet, J., Boehm, A., & Bennett, C. L. (2026). Flush with data: harnessing emergency department wastewater as an innovative approach for surveillance of infectious diseases. American Journal of Epidemiology, kwag019, kwag019. 10.1093/aje/kwag019

Ren, J., Song, K., Deng, C., Ahlgren, N. A., Fuhrman, J. A., Li, Y., Xie, X., Poplin, R., & Sun, F. (2020). Identifying viruses from metagenomic data using deep learning. Quantitative Biology (Beijing, China), 8(1), 64–77. 10.1007/s40484-019-0187-4

Robinson, C. A., Hsieh, H.-Y., Hsu, S.-Y., Wang, Y., Salcedo, B. T., Belenchia, A., Klutts, J., Zemmer, S., Reynolds, M., Semkiw, E., Foley, T., Wan, X., Wieberg, C. G., Wenzel, J., Lin, C.-H., & Johnson, M. C. (2022). Defining biological and biophysical properties of SARS-CoV-2 genetic material in wastewater. The Science of the Total Environment, 807(Pt 1), 150786. 10.1016/j.scitotenv.2021.150786

Rothman, J. A., Loveless, T. B., Kapcia, J., 3rd, Adams, E. D., Steele, J. A., Zimmer-Faust, A. G., Langlois, K., Wanless, D., Griffith, M., Mao, L., Chokry, J., Griffith, J. F., & Whiteson, K. L. (2021). RNA viromics of Southern California wastewater and detection of SARS-CoV-2 single-nucleotide variants. Applied and Environmental Microbiology, 87(23), e0144821. 10.1128/AEM.01448-21

Rushford, C., Gregory, D. A., Copen, E. E., Naydenov, J., Tovar-Mendez, A., Li, P.-E., Kantor, R. S., Shakya, M., Ruth, N., Chain, P. S. G., Hsieh, H.-Y., Hunter, T., Kome, J., Frank, M., Lyddon, T. D., Kaufman, J., O’Connor, D. H., & Johnson, M. C. (2025). Untargeted longitudinal ultra deep metagenomic sequencing of wastewater provides a comprehensive readout of expected and unexpected viral pathogens. In medRxiv (p. 2025.10.27.25338874). 10.1101/2025.10.27.25338874

Schuele, L., Masirika, L. M., Udahemuka, J. C., Siangoli, F. B., Mbiribindi, J. B., Ndishimye, P., Aarestrup, F. M., Koopmans, M., Oude Munnink, B. B., Molenkamp, R., GREATLIFE MPOX group, & Collaborators. (2024). Real-time PCR assay to detect the novel Clade Ib monkeypox virus, September 2023 to May 2024. Euro Surveillance : Bulletin Europeen Sur Les Maladies Transmissibles [Euro Surveillance : European Communicable Disease Bulletin], 29(32), 2400486. 10.2807/1560-7917.ES.2024.29.32.2400486

Sikorski, M. J., & Levine, M. M. (2020). Reviving the “Moore swab”: A classic environmental surveillance tool involving filtration of flowing surface water and sewage water to recover typhoidal Salmonella bacteria. Applied and Environmental Microbiology, 86(13), e00060–20. 10.1128/AEM.00060-20

Spurbeck, R. R., Catlin, L. A., Mukherjee, C., Smith, A. K., & Minard-Smith, A. (2023). Analysis of metatranscriptomic methods to enable wastewater-based biosurveillance of all infectious diseases. Frontiers in Public Health, 11. 10.3389/fpubh.2023.114527510.3389/fpubh.2023.1145275.s001

St-Onge, G., Davis, J. T., Hébert-Dufresne, L., Allard, A., Urbinati, A., Scarpino, S. V., Chinazzi, M., & Vespignani, A. (2025). Pandemic monitoring with global aircraft-based wastewater surveillance networks. Nature Medicine, 31(3), 788–796. 10.1038/s41591-025-03501-4

Symonds, E. M., Nguyen, K. H., Harwood, V. J., & Breitbart, M. (2018). Pepper mild mottle virus: A plant pathogen with a greater purpose in (waste)water treatment development and public health management. Water Research, 144, 1–12. 10.1016/j.watres.2018.06.066

Tatusov, R. L., Chhabra, P., Diez-Valcarce, M., Barclay, L., Cannon, J. L., & Vinjé, J. (2021). Human Calicivirus Typing tool: A web-based tool for genotyping human norovirus and sapovirus sequences. Journal of Clinical Virology: The Official Publication of the Pan American Society for Clinical Virology, 134(104718), 104718. 10.1016/j.jcv.2020.104718

Tay, A. P., Didi, K., Wickramarachchi, A., Bauer, D. C., Wilson, L. O. W., & Maselko, M. (2024). Synsor: a tool for alignment-free detection of engineered DNA sequences. Frontiers in Bioengineering and Biotechnology, 12, 1375626. 10.3389/fbioe.2024.1375626

The Nucleic Acid Observatory Consortium. (2021). A Global Nucleic Acid Observatory for Biodefense and Planetary Health. In arXiv [q-bio.GN]. arXiv. http://arxiv.org/abs/2108.02678

Tisza, M., Javornik Cregeen, S., Avadhanula, V., Zhang, P., Ayvaz, T., Feliz, K., Hoffman, K. L., Clark, J. R., Terwilliger, A., Ross, M. C., Cormier, J., Moreno, H., Wang, L., Payne, K., Henke, D., Troisi, C., Wu, F., Rios, J., Deegan, J.,… Maresso, A. W. (2023). Wastewater sequencing reveals community and variant dynamics of the collective human virome. Nature Communications, 14(1), 6878. 10.1038/s41467-023-42064-1

Turakhia, Y., Thornlow, B., Hinrichs, A. S., De Maio, N., Gozashti, L., Lanfear, R., Haussler, D., & Corbett-Detig, R. (2021). Ultrafast Sample placement on Existing tRees (UShER) enables real-time phylogenetics for the SARS-CoV-2 pandemic. Nature Genetics, 53(6), 809–816. 10.1038/s41588-021-00862-7

WastewaterSCAN. (2025). EVD68, California. WastewaterSCAN Dashboard. https://data.wastewaterscan.org/tracker?charts=CvYDEAAgAUgAUgY3NGQwMjZSBjA3Y2VkN1IGYjcyM2FlUgYzMDMyYzhSBjI0ZDAzMFIGMTEwN2I2UgY2NTdlYTVSBjY0NzNjMFIGN2NhOTA1UgYxZDYxNDlSBjMyNDllZlIGNWFhNjk5UgYzYTNmYTFSBjc0YjM5YVIGZmEyZDYzUgYwMzI1ZGRSBmI5MjVlN1IGMWE0M2E3UgZhMGZiY2RSBjI1NDgxOVIGNTc1NzM4UgY4YTliNGJSBjI3ODQ3MFIGY2RjZWFkUgYwMmQyNDJSBjcxYTJmNFIGYmNiYjg1UgZiYzc5ZjlSBmU5ZTg3ZVIGZWVjMmMyUgYyZTA3ZTZSBjU3OWRhM1IGYTEwODZmUgY3YWExMzZSBmMyMjE5N1IGYjljMDJkUgY2MjkyMmZSBjM3MzcwMlIGZGRlODhmUgYzMTlhYTZSBjg0ZDQ4OVIGMGM4MDkxUgZhZDg2YTlSBmM4ZDM1N1IGMzE3NDU0UgZlZDlhMWZSBmE4MmNlOVIGY2IwZTFjUgYzNzQzMGFSBjY3YzJlYlIGZGQzNmZiUgZmZWQ5N2ZSBjE4OTFlMFIGZWRlZmI0UgYyOTNiMjVSBjYwOWZjOFIGYzk1ZTY0WgVFVkQ2OHIKMjAyNS0wNy0yNXIKMjAyNS0xMi0yOIoBBjI0Y2RmMA%3D%3D&selectedChartId=24cdf0

Wetterstrand, K. A. (2019, March 13). DNA Sequencing Costs: Data from the NHGRI Genome Sequencing Program (GSP). Genome.gov; NHGRI. https://www.genome.gov/about-genomics/fact-sheets/DNA-Sequencing-Costs-Data

Wittmann, B. J., Alexanian, T., Bartling, C., Beal, J., Clore, A., Diggans, J., Flyangolts, K., Gemler, B. T., Mitchell, T., Murphy, S. T., Wheeler, N. E., & Horvitz, E. (2025). Strengthening nucleic acid biosecurity screening against generative protein design tools. Science (New York, N.Y.), 390(6768), 82–87. 10.1126/science.adu8578

Wolfe, M. K., Duong, D., Hughes, B., Chan-Herur, V., White, B. J., & Boehm, A. B. (2022). Detection of monkeypox viral DNA in a routine wastewater monitoring program. In medRxiv (p. 2022.07.25.22278043). medRxiv. 10.1101/2022.07.25.22278043

Wolfe, M. K., North, D., Jaffe, A. L., Zulli, A., Duong, D., Shelden, B., Goldman, M., Richardson, M., Thana, P., Chan-Herur, V., Kheradpour, P., Bidwell, A. L., Hilton, S. P., Conforti, S., Paulos, A. P., & Boehm, A. B. (2026). Prospective metagenomic sequencing of wastewater across the United States yields robust viral enrichment and concordance with digital PCR measurements. Applied and Environmental Microbiology, e01020–26. 10.1128/aem.01020-26

Wolfe, M. K., Topol, A., Knudson, A., Simpson, A., White, B., Vugia, D. J., Yu, A. T., Li, L., Balliet, M., Stoddard, P., Han, G. S., Wigginton, K. R., & Boehm, A. B. (2021). High-frequency, high-throughput quantification of SARS-CoV-2 RNA in wastewater settled solids at eight publicly owned treatment works in northern California shows strong association with COVID-19 incidence. mSystems, 6(5), e0082921. 10.1128/mSystems.00829-21

Wongprommoon, A., Chomkatekaew, C., & Chewapreecha, C. (2024). Monitoring pathogens in wastewater. Nature Reviews. Microbiology, 22(5), 261. 10.1038/s41579-024-01033-1

Wood, D. E., Lu, J., & Langmead, B. (2019). Improved metagenomic analysis with Kraken 2. Genome Biology, 20(1), 257. 10.1186/s13059-019-1891-0

World Health Organization. (une 2024). Pathogens prioritization: a scientific framework for epidemic and pandemic research preparedness. https://www.who.int/publications/m/item/pathogens-prioritization-a-scientific-framework-for-epidemic-and-pandemic-research-preparedness

Worp, N., Nieuwenhuijse, D. F., Izquierdo-Lara, R. W., Schapendonk, C. M. E., Brinch, C., Jensen, E. E. B., Munk, P., Hendriksen, R. S., Global Sewage Surveillance Consortium, Aarestrup, F., Oude Munnink, B. B., Koopmans, M. P. G., & de Graaf, M. (2025). Unveiling the global urban virome through wastewater metagenomics. Nature Communications, 16(1), 10707. 10.1038/s41467-025-65208-x

Wu, F., Oghuan, J., Gitter, A., Mena, K. D., & Brown, E. L. (2023). Wide mismatches in the sequences of primers and probes for monkeypox virus diagnostic assays. Journal of Medical Virology, 95(1), e28395. 10.1002/jmv.28395

Zulli, A., Bowie, K. R., Ruedaflores, M., Sahly, J., Angel, D., Rohatgi, K., Malik, W. S., Hao, R. M., Shepherd, J., & Peccia, J. (2025). Probe-based enrichment sequencing applied to wastewater surveillance accurately tracks multiple viral respiratory outbreaks. ACS ES&T Water, 5(8), 4575–4583. 10.1021/acsestwater.5c00262

