## Supplementary material for "Deep untargeted wastewater metagenomic sequencing from sewersheds across the United States": si

**Affiliations**
[1]Massachusetts Institute of Technology, Media Lab, Cambridge, MA, USA
[2]The Charles Stark Draper Laboratory, Inc., Draper Scholar, Cambridge, MA, USA
[3]Broad Institute of MIT and Harvard, Cambridge, MA, USA
[4]University of Missouri, Department of Veterinary Pathobiology, Columbia, MO, USA
[5]SecureBio, Cambridge, MA, USA
[6]University of Missouri, Department of Molecular Microbiology and Immunology, Columbia, MO, USA
[7]University of Oklahoma, School of Civil Engineering and Environmental Science, Norman, OK, USA
[8]University of Miami, Department of Chemical, Environmental, and Materials Engineering, Miami, FL, USA
[9]Queens College of The City University of New York, Biology Department, Flushing, NY, USA
[10]NYC Health + Hospitals, New York, NY, USA
[11]Riverside Water Quality Control Plant, Riverside, CA, USA
[12]City of Boise, Boise, ID, USA
[13]Inland Empire Utilities Agency, Chino, CA, USA
[14]Metropolitan Water Reclamation District of Greater Chicago, Chicago, IL, USA
[15]Miami-Dade Water and Sewer Department, Miami, FL, USA
[16]City of Palo Alto, Palo Alto, CA, USA
[17]City of Kansas City Missouri, KC Health, Communicable Disease and Prevention Division, Kansas City, MO
[18]City of Kansas City Missouri, KC Water, Water and Wastewater Operations, Kansas City, MO
[19]City of Kansas City Missouri, KC Water, Wastewater Treatment Division, Kansas City, MO

[20]The Charles Stark Draper Laboratory, Inc., Cambridge, MA, USA
[21]Massachusetts General Hospital, Division of Infectious Diseases, Boston, MA, USA
[22]Howard Hughes Medical Institute, Chevy Chase, MD, USA
[23]Massachusetts General Hospital, Department of Pathology, Boston, MA, USA
[24]Harvard Medical School, Boston, MA, USA
[25]Lawrence Livermore National Laboratory, Physical and Life Sciences Directorate, Livermore, CA, USA
[26]University of Wisconsin-Madison, Department of Pathology and Laboratory Medicine, Madison, WI, USA
[27]The Graduate Center of The City University of New York, New York, NY, USA
[28]University of Illinois Chicago, Department of Biological Sciences, Chicago, IL, USA
[29]University of California, Riverside, Department of Microbiology and Plant Pathology, Riverside, CA, USA
[30]Harvard University, Harvard T.H. Chan School of Public Health, Department of Immunology and Infectious Diseases, Boston, MA, USA
[31]Harvard University, Faculty of Arts and Sciences, Department of Organismic and Evolutionary Biology, Cambridge, MA, USA

#### Table of contents

##### Figures


| | | |
|---|---|---|
| Supplementary Figure S1. | Sequencing depth (total read pairs) over time by site and sequencing partner. | 4 |
| Supplementary Figure S2. | CASPER contribution to untargeted wastewater metagenomic sequencing (WW-MGS) data on the NCBI Sequence Read Archive (SRA) | 5 |
| Supplementary Figure S3. | Sample turnaround time from collection to sequencing data delivery. | 6 |
| Supplementary Figure S4. | Taxonomic composition over time by site. | 7 |
| Supplementary Figure S5. | Effect of ribodepletion on vertebrate-infecting virus (VV) read fraction. | 8 |
| Supplementary Figure S6. | Size of the shallow taxonomic profiling subsample and tobamovirus counts. | 9 |
| Supplementary Figure S7. | Classification accuracy of the vertebrate-infecting virus (VV) workflow on simulated reads. | 10 |
| Supplementary Figure S8. | Vertebrate-infecting virus family composition over time by site. | 11 |
| Supplementary Figure S9. | Paired comparison of the University of Missouri (MU) and SecureBio (SB) protocols at Boston Deer Island Treatment Plant (DITP). | 12 |
| Supplementary Figure S10. | Temporal abundance of select viruses across CASPER sites (log scale). | 13 |
| Supplementary Figure S11. | Read pairs assigned to select pathogen clades over time across CASPER sites (log scale). | 14 |
| Supplementary Figure S12. | Agreement between wastewater metagenomic sequencing and wastewater PCR measurements across all paired sites. | 15 |
| Supplementary Figure S13. | Clinical respiratory virus testing from Massachusetts General Hospital. | 16 |
| Supplementary Figure S14. | GC content over time by site. | 17 |
| Supplementary Figure S15. | Sequencing quality scores over time by site. | 18 |
| Supplementary Figure S16. | Read length over time by site. | 19 |
| Supplementary Figure S17. | Quality control (QC) pass rate over time by site. | 20 |


##### Tables


| | | |
|---|---|---|
| Supplementary Table S1. | Summary of sequencing depth across published wastewater metagenomic sequencing (WW-MGS) studies. | 21 |
| Supplementary Table S2. | Historical cost of sequencing a typical CASPER sample. | 22 |
| Supplementary Table S3. | Correlation between wastewater metagenomic sequencing and wastewater PCR data. | 23 |
| Supplementary Table S4. | Correlation between wastewater metagenomic sequencing and clinical testing data. | 24 |
| Supplementary Table S5. | Composition and classification accuracy of the benchmarking panels. | 25 |
| Supplementary Table S6. | Classification accuracy of the vertebrate-infecting virus (VV) workflow by viral family. | 26 |
| Supplementary Table S7. | Agreement between the University of Missouri (MU) and SecureBio (SB) protocols on paired samples from Boston Deer Island Treatment Plant (DITP). | 27 |
| Supplementary Table S8. | Read pairs assigned to the pathogens shown in Figure 4c. | 28 |
| Supplementary Table S9. | National Wastewater Surveillance System (NWSS) sewershed identifiers for CASPER sites. | 29 |
| Supplementary Table S10. | Wastewater PCR assay details and data source designations for CASPER comparison sites. | 30 |
| Supplementary Table S11. | SecureBio (SB) samples prepared with ribodepletion reagent omission. | 31 |

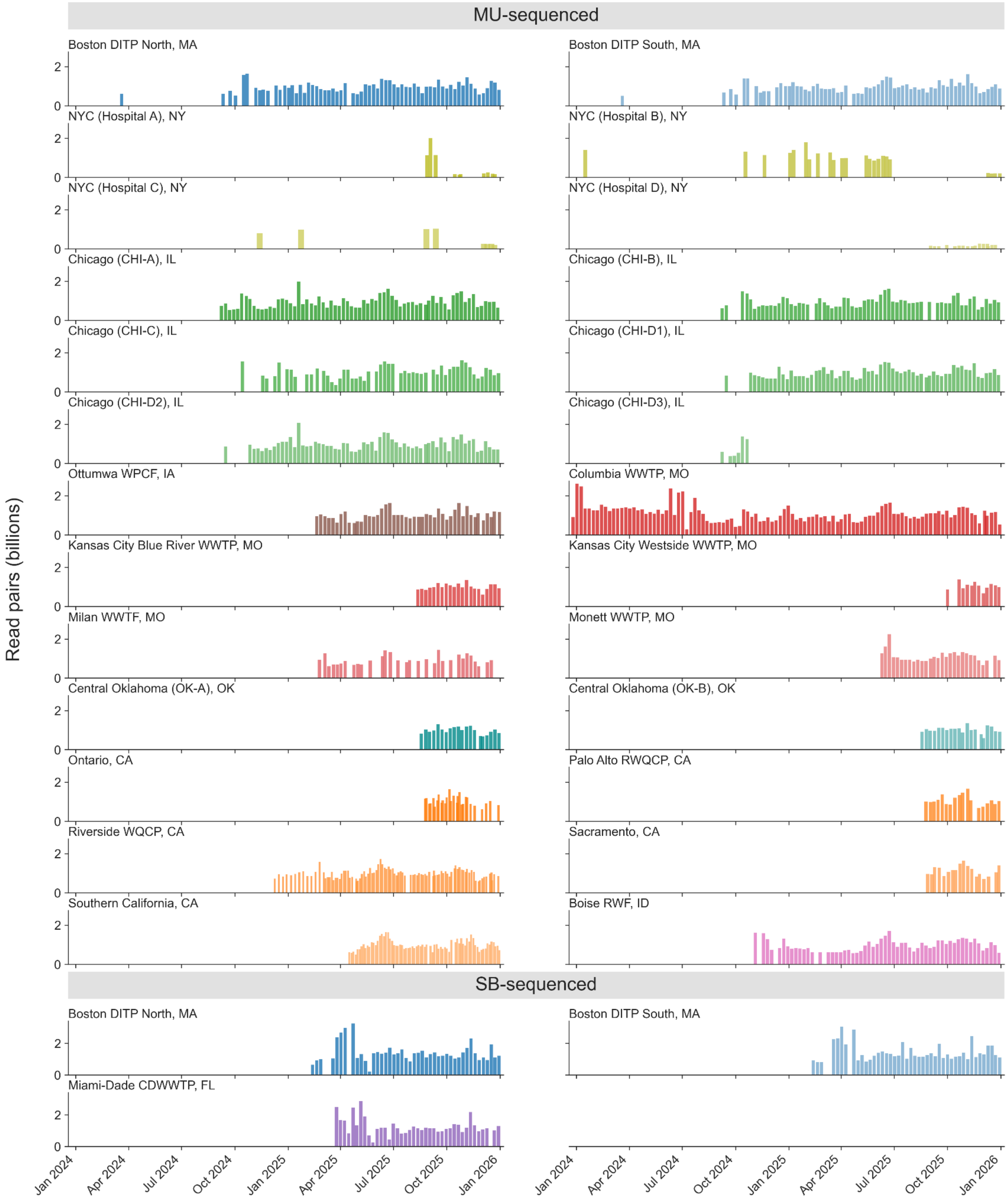


**Supplementary Figure S1. Sequencing depth (total read pairs) over time by site and sequencing partner.**

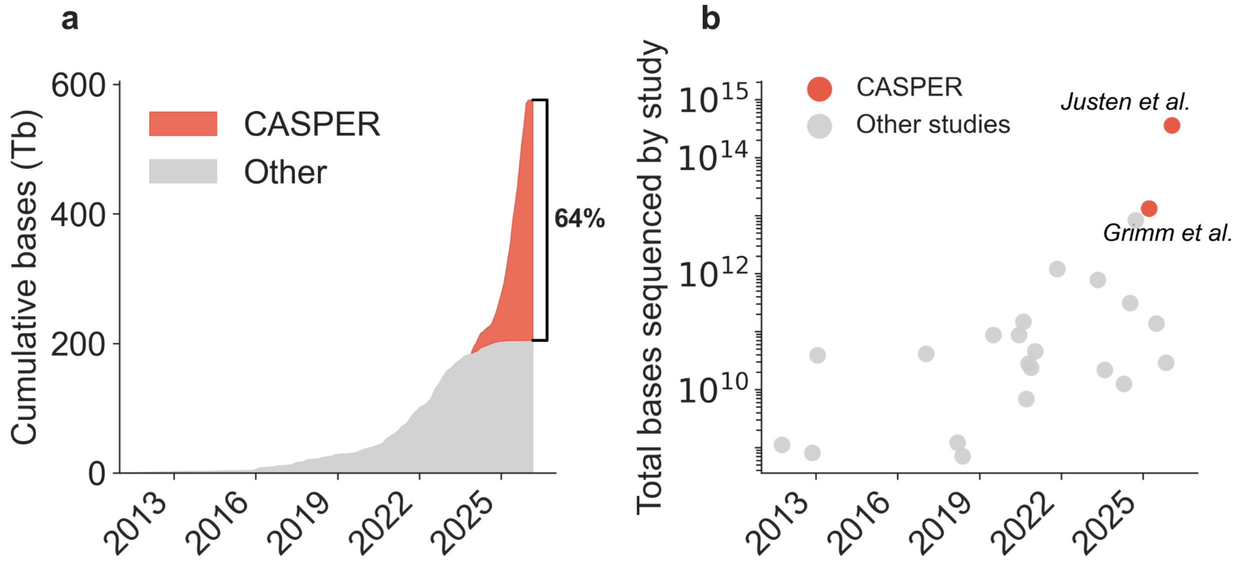


**Supplementary Figure S2. CASPER contribution to untargeted wastewater metagenomic sequencing (WW-MGS) data on the NCBI Sequence Read Archive (SRA).** (a) CASPER data from this release (PRJNA1247874; 358 Tb) and (Grimm et al., 2025) (PRJNA1198001; 13 Tb) as a fraction of the cumulative untargeted WW-MGS bases deposited on the NCBI SRA over time. (b) Total untargeted WW-MGS bases by study for publicly available WW-MGS datasets identified in our literature survey (Supplementary Table S1).

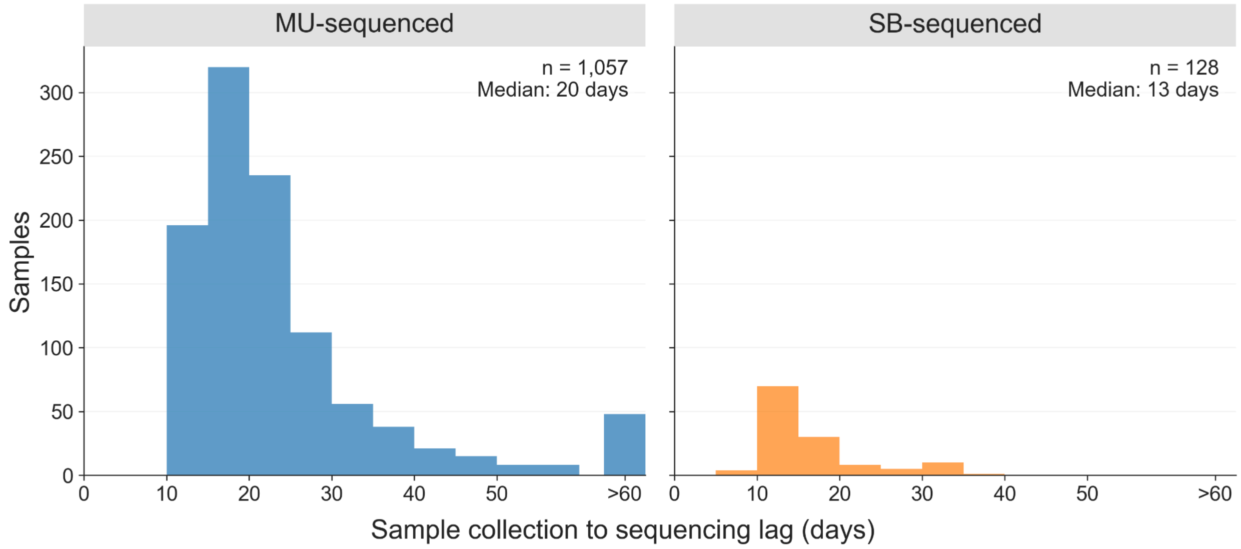


**Supplementary Figure S3. Sample turnaround time from collection to sequencing data delivery.** Distribution of turnaround time (days) by site and sequencing laboratory. The upper range reflects NYC hospital grab samples that were stored at 4°C for several months prior to extraction during a pilot phase.

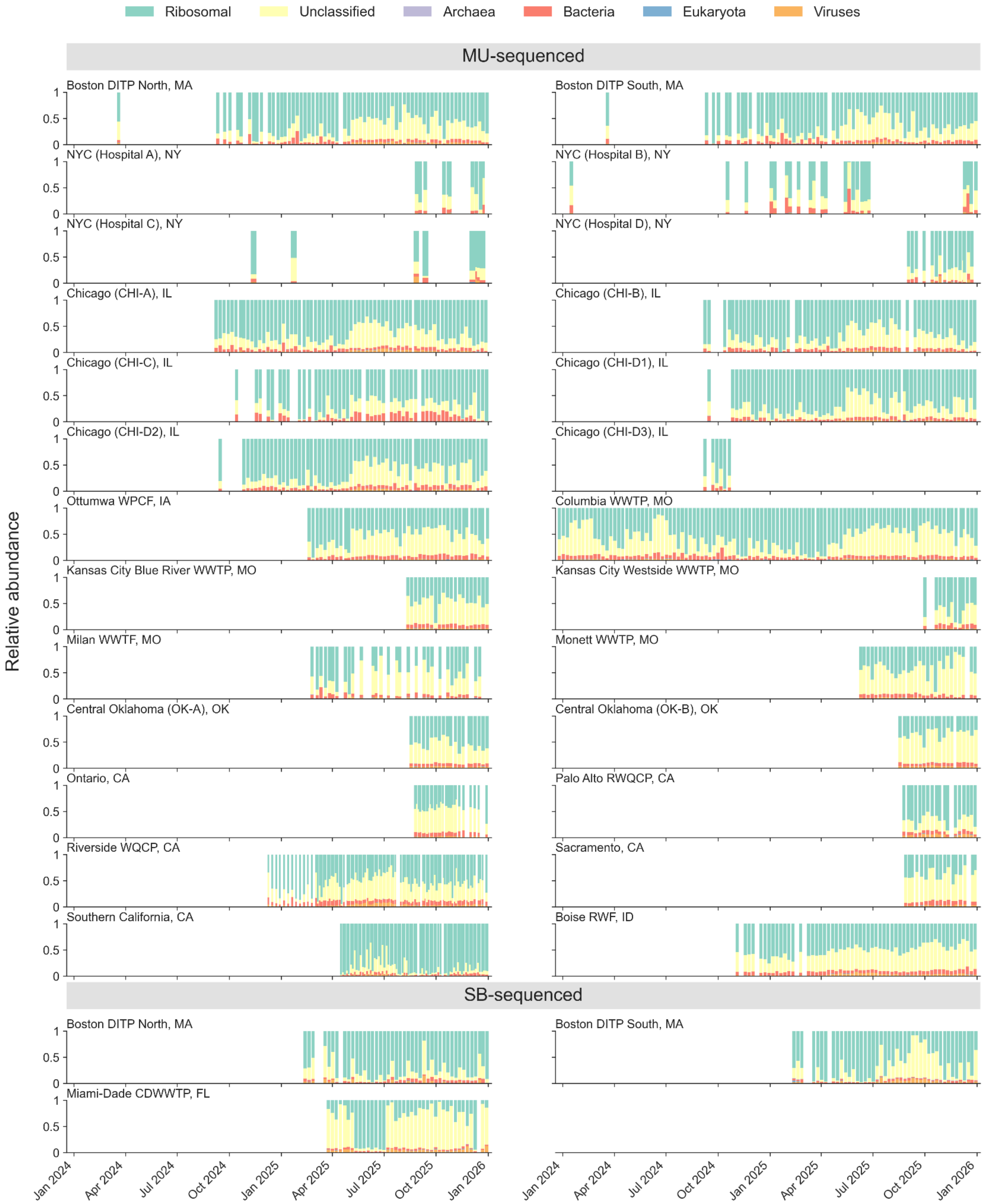


**Supplementary Figure S4. Taxonomic composition over time by site.** Proportion of reads assigned to major taxonomic categories for each sampling location, grouped by sequencing partner.

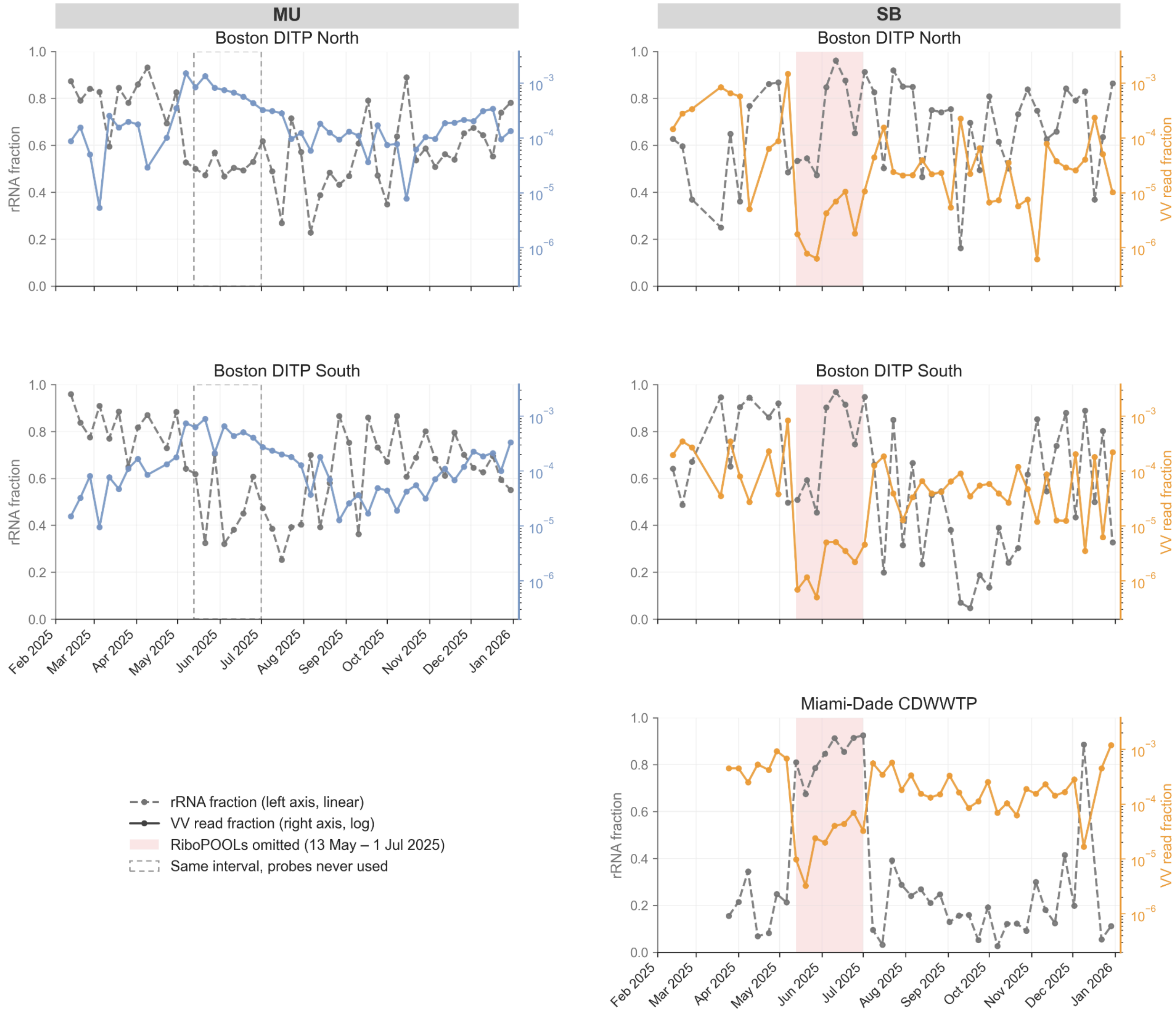


**Supplementary Figure S5. Effect of ribodepletion on vertebrate-infecting virus (VV) read fraction.** Ribosomal RNA (rRNA) fraction (grey, left axis) and VV read fraction (coloured, right axis, log scale) over time for paired Boston Deer Island Treatment Plant (DITP) North and South samples processed by the University of Missouri (MU, left column) and SecureBio (SB, right column), and for Miami-Dade Central District Wastewater Treatment Plant (CDWWTP), FL, processed by SB. The MU protocol does not include a ribodepletion step; the SB protocol applies RiboPOOLs probes, which were omitted from SB library preparation between 13 May and 1 July 2025 (shaded). Dashed boxes mark the same interval in MU samples, where no probes were used at any point. Affected SB accessions are listed in Supplementary Table S11.

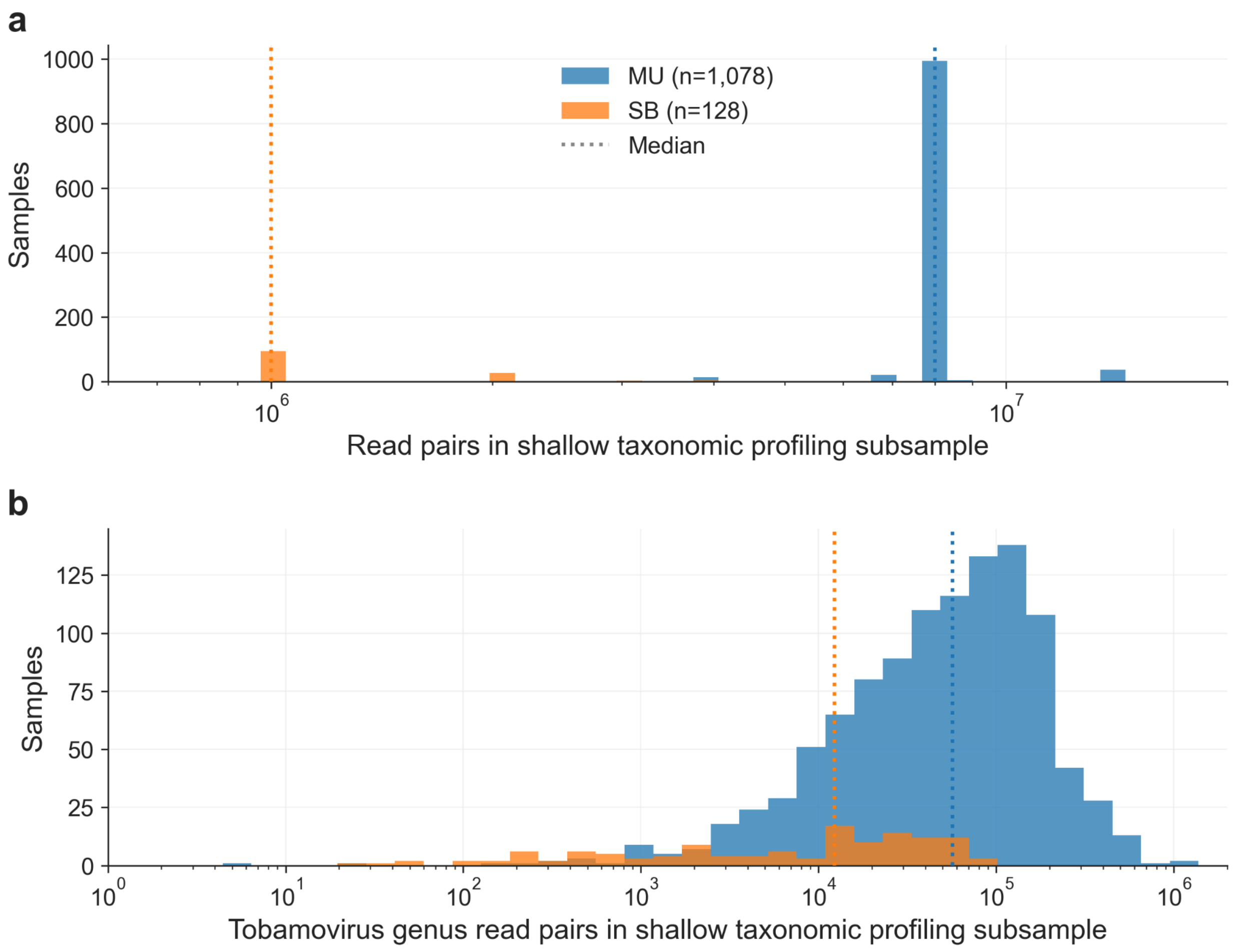


**Supplementary Figure S6. Size of the shallow taxonomic profiling subsample and tobamovirus counts.** (a) Read pairs subsampled by the shallow taxonomic profiling workflow (Methods). The pipeline subsamples 1M read pairs per sequencing lane, typically yielding 8M read pairs for samples processed at the University of Missouri (MU) and 1M for samples processed by SecureBio (SB). (b) Read pairs assigned to the tobamovirus clade by Kraken2 against the Standard database in the shallow taxonomic profiling subsample.

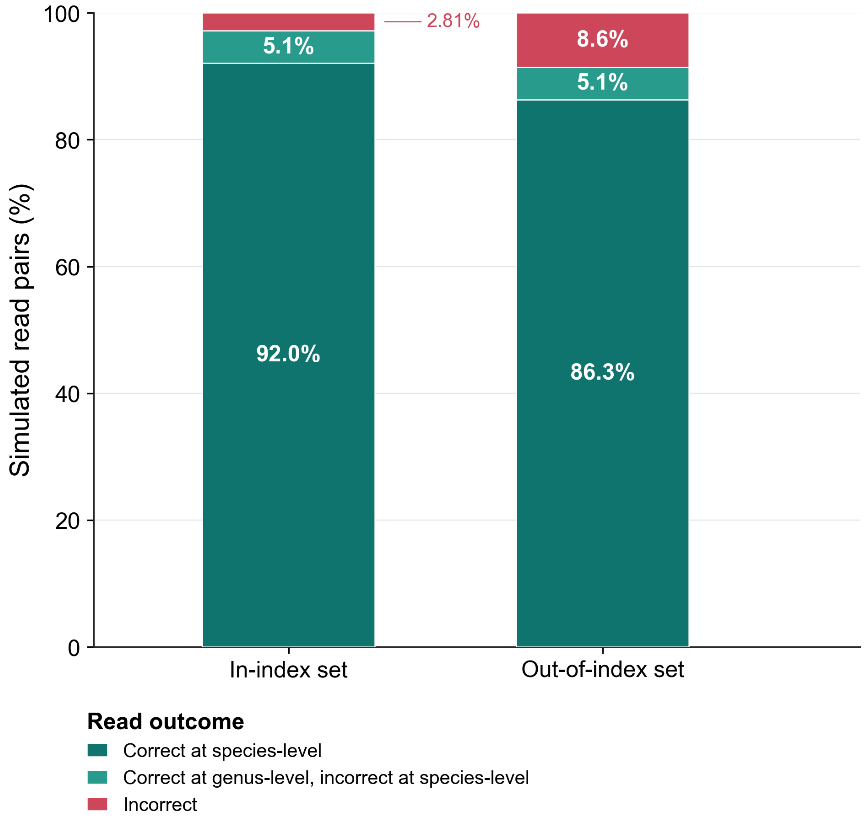


**Supplementary Figure S7. Classification accuracy of the vertebrate-infecting virus (VV) workflow on simulated reads.** Paired-end reads were simulated from reference genomes of 93 notable vertebrate-infecting virus taxa, processed through the VV identification workflow, and scored against the genome each read was simulated from. The in-index set comprises genomes drawn from the workflow's reference database (93 genomes, 92 species); the out-of-index set comprises GenBank genomes absent from that database, deposited after its build date (76 genomes, 76 species). Read pairs are scored as correct at species level, correct at genus level but incorrect at species level, or incorrect at both. Panel construction, read simulation, and scoring are described in Methods.

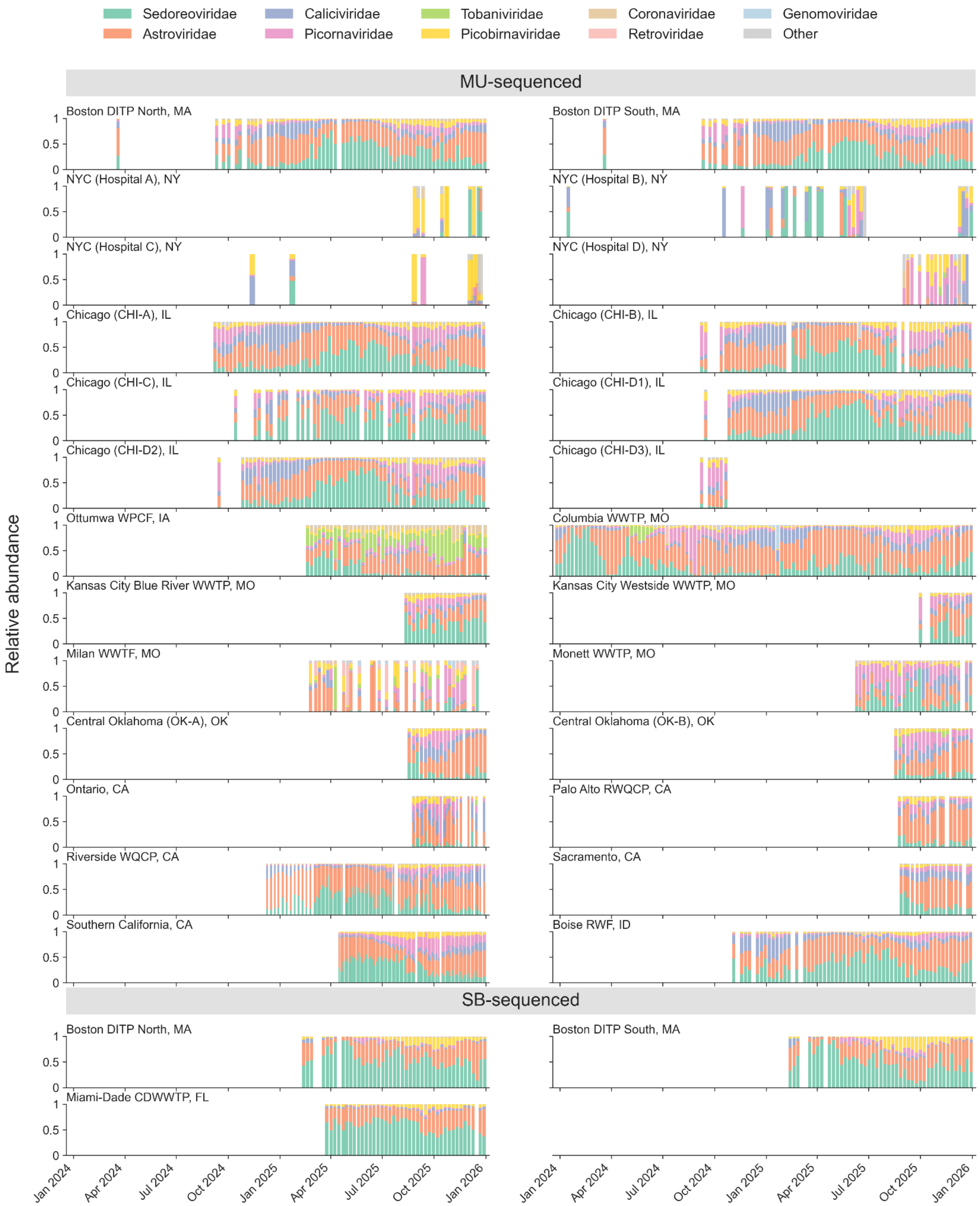


**Supplementary Figure S8. Vertebrate-infecting virus family composition over time by site.** Relative abundance of vertebrate-infecting virus (VV) reads by viral family for each sampling location, grouped by sequencing partner.

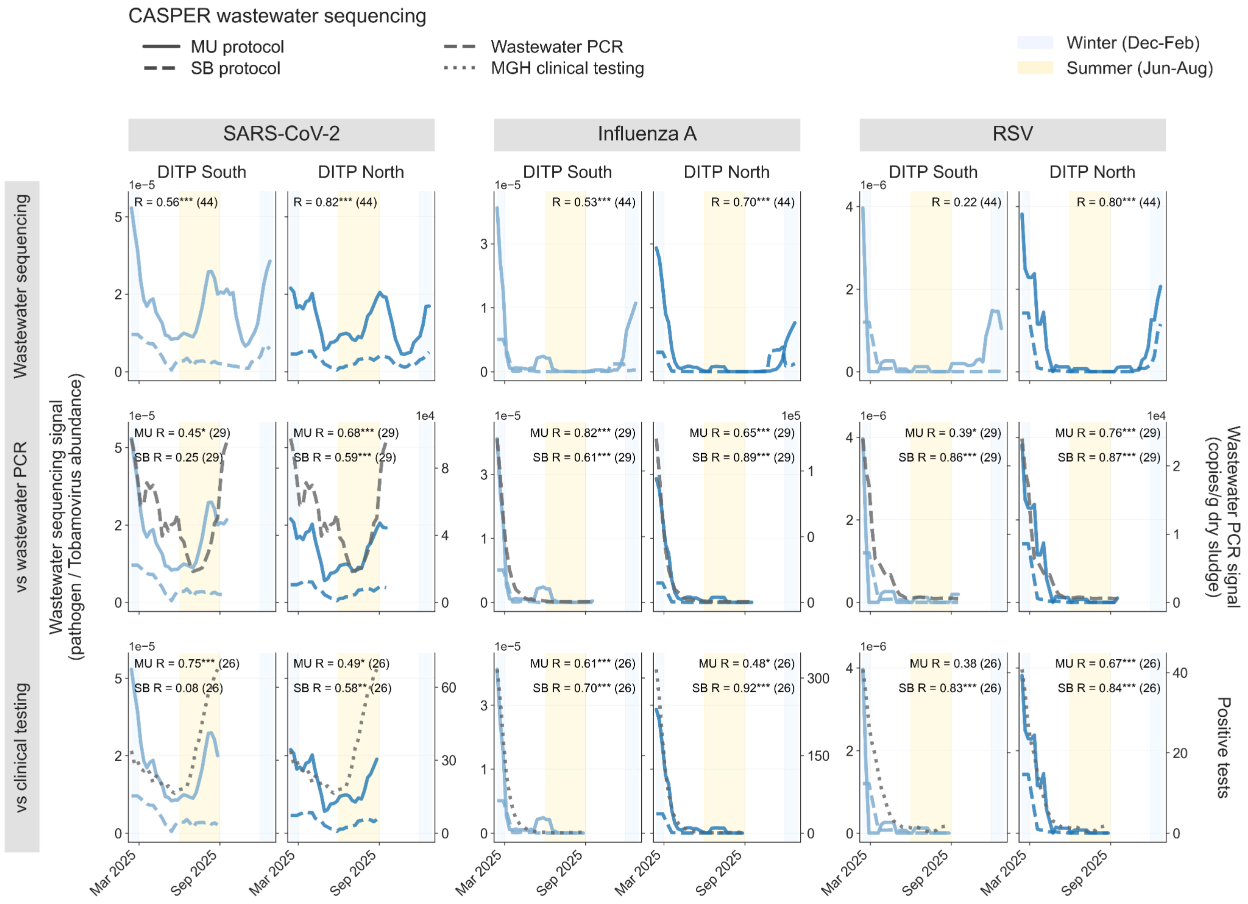


**Supplementary Figure S9. Paired comparison of the University of Missouri (MU) and SecureBio (SB) protocols at Boston Deer Island Treatment Plant (DITP).** DITP North and South samples were split between the two laboratories, giving 44 collection dates per system (February–December 2025) on which both protocols processed the same wastewater. Rows show the two protocols against each other, each protocol against wastewater PCR reported to the National Wastewater Surveillance System by WastewaterSCAN (copies per gram of dry sludge), and each protocol against Massachusetts General Hospital (MGH) clinical testing (weekly positive tests). Spearman correlation coefficients (R) were calculated on MMWR-week series smoothed with a centered five-week moving average, restricted to weeks covered by both laboratories, with the number of contributing weeks given in parentheses. *p < 0.05, **p < 0.01, ***p < 0.001. Values are tabulated in Supplementary Table S7.

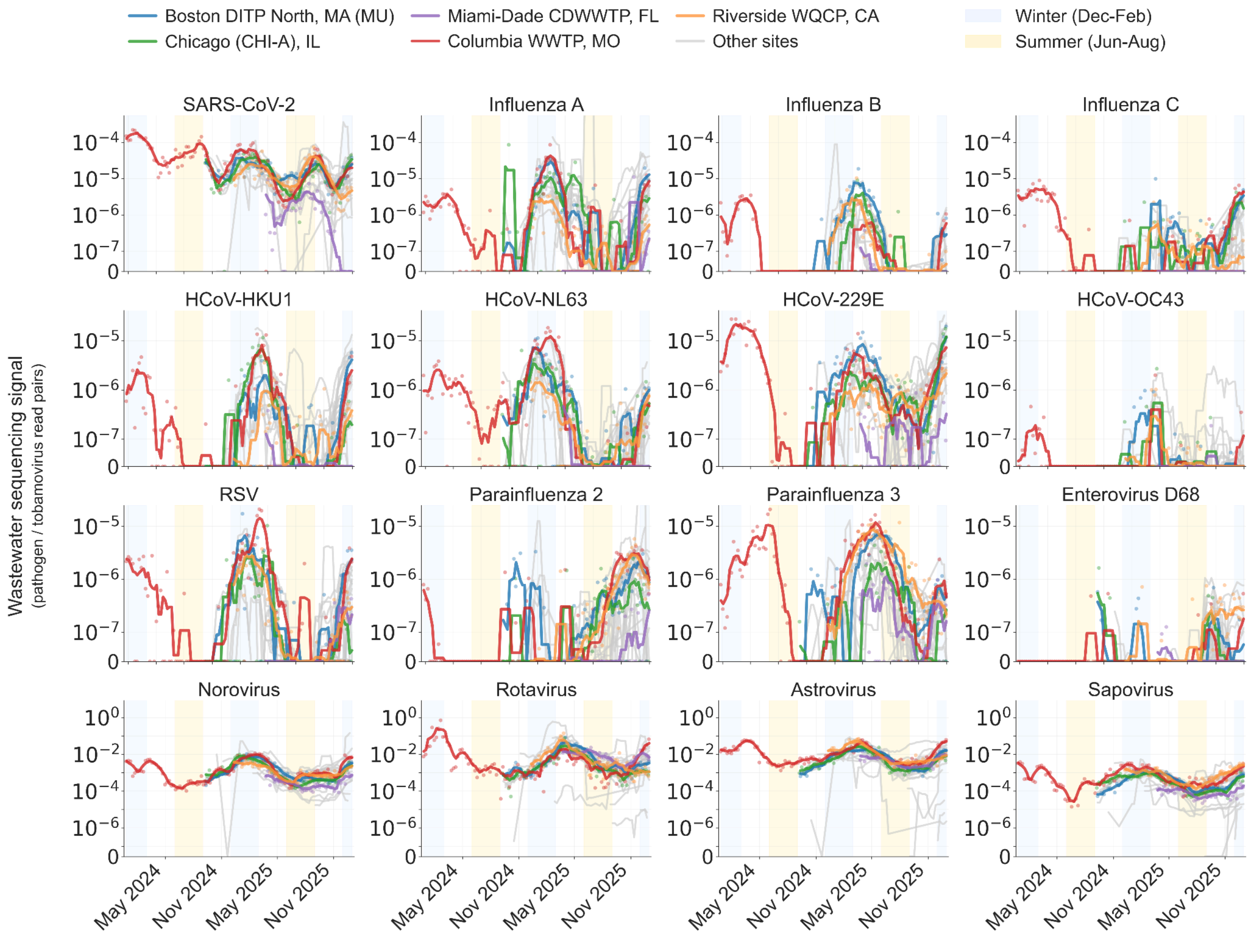


**Supplementary Figure S10. Temporal abundance of select viruses across CASPER sites (log scale).** Tobamovirus-normalized abundance of human-infecting respiratory viruses (top rows) and gastrointestinal viruses (bottom row) over time.

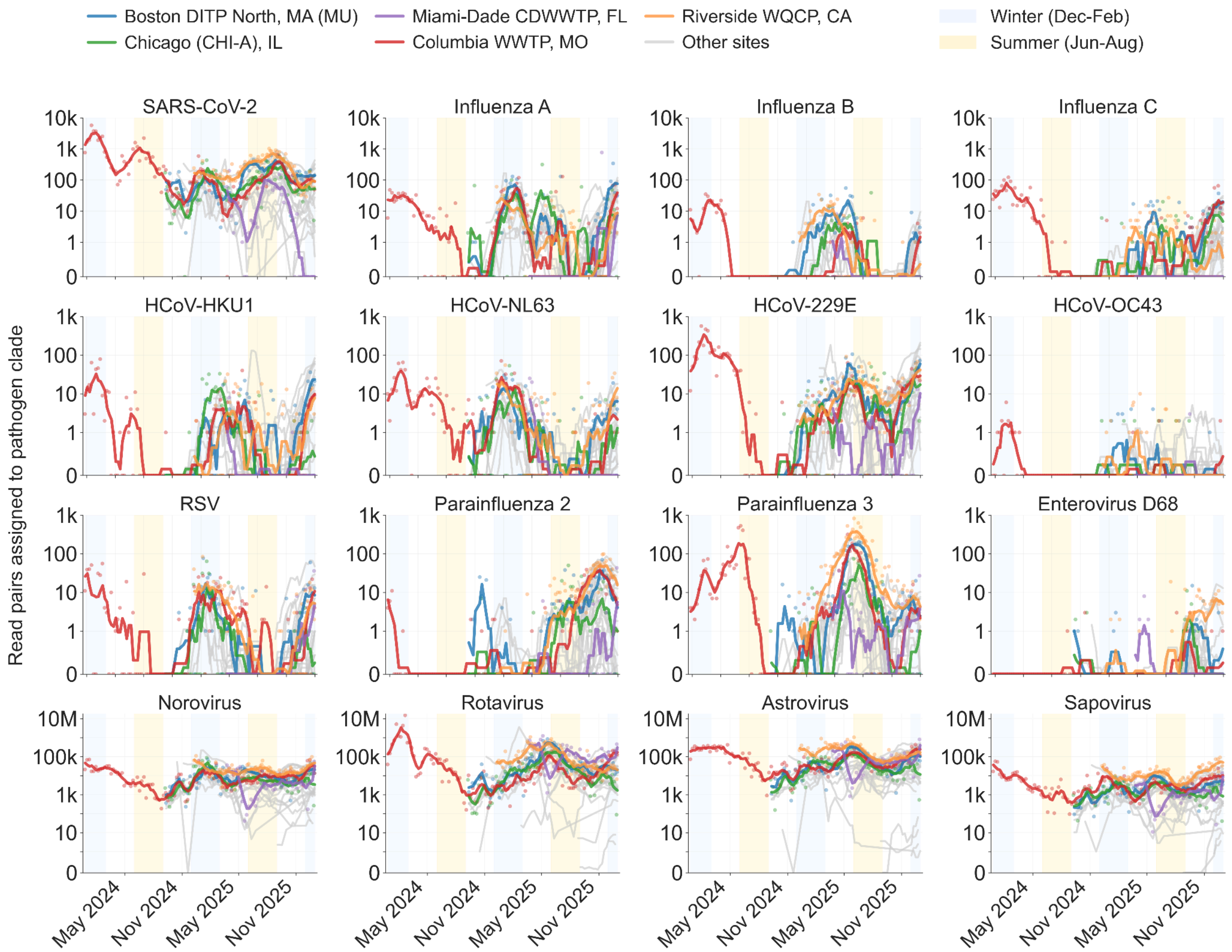


**Supplementary Figure S11. Read pairs assigned to select pathogen clades over time across CASPER sites (log scale).** Reads were assigned using the vertebrate-infecting virus identification pipeline (Methods). Sequencing depth targets 1 billion reads per sample but varies across samples and sites (Supplementary Figure S1).

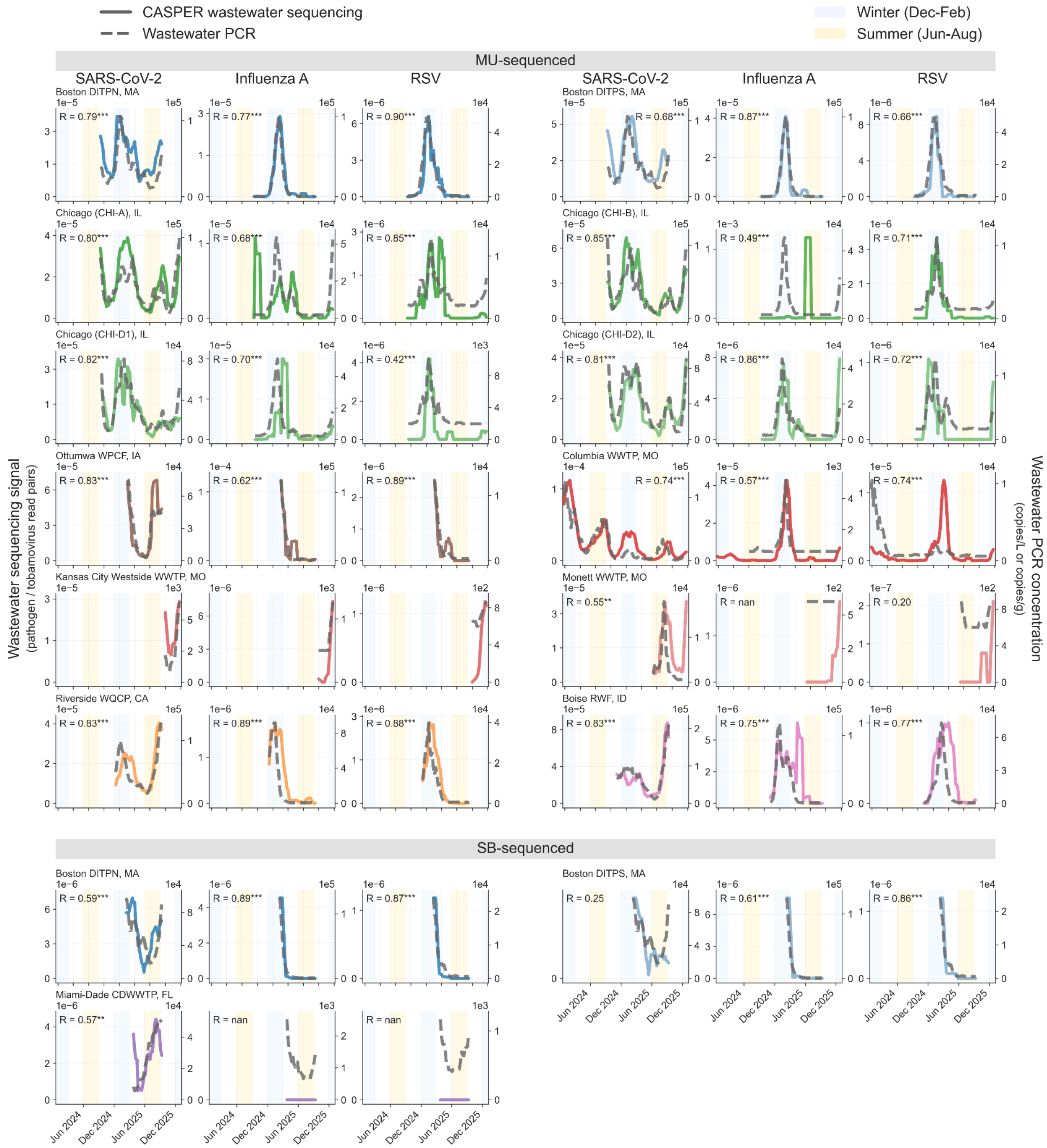


**Supplementary Figure S12. Agreement between wastewater metagenomic sequencing and wastewater PCR measurements across all paired sites.** Tobamovirus-normalized wastewater metagenomic sequencing abundance compared with wastewater PCR concentrations from the CDC National Wastewater Surveillance System data portal. PCR concentrations are shown in each source's reported units (Methods). Time series data are aggregated to MMWR epidemiological weeks and smoothed with a 5-week centered moving average. Spearman correlation coefficients (R) calculated on week-aligned smoothed values. *p < 0.05, **p < 0.01, ***p < 0.001. MU: University of Missouri; SB: SecureBio.

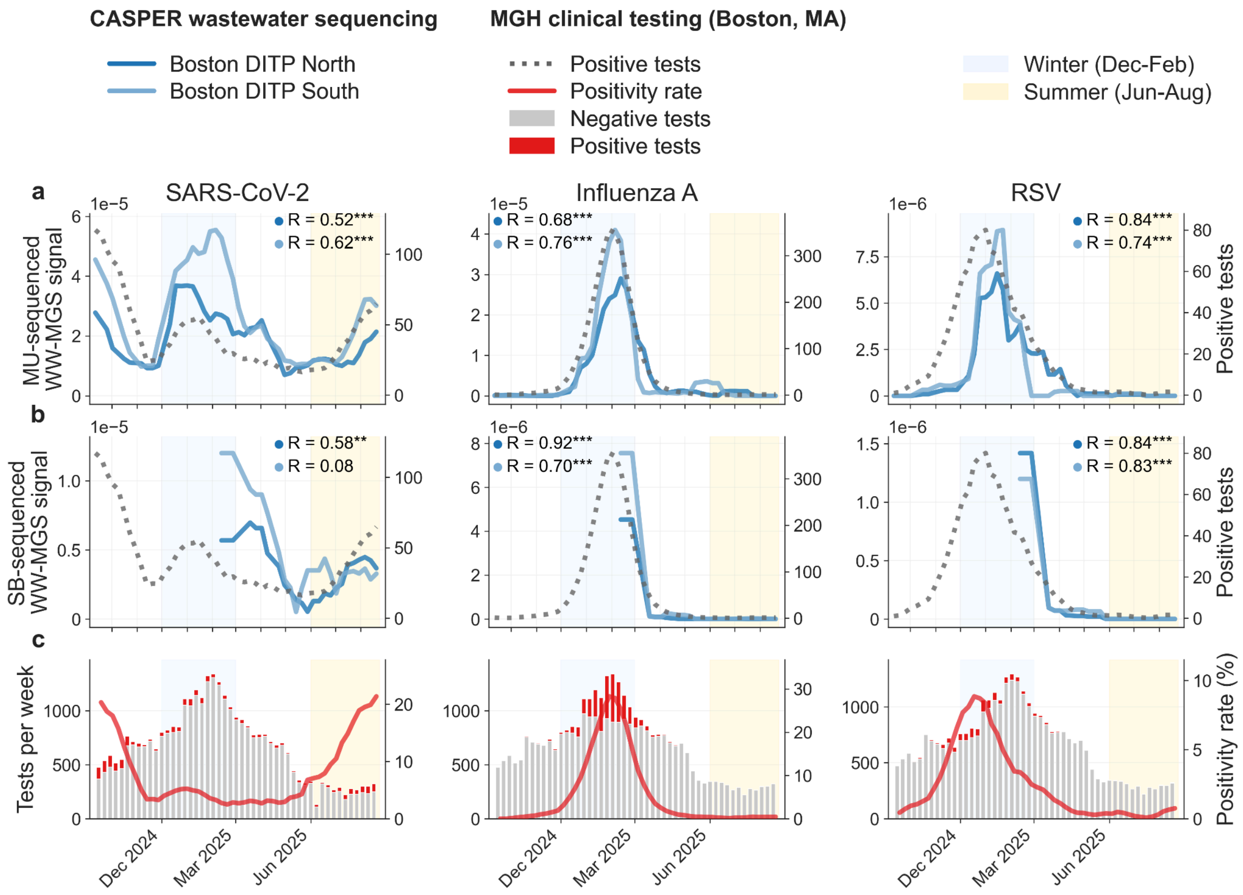


**Supplementary Figure S13. Clinical respiratory virus testing from Massachusetts General Hospital.** (a) University of Missouri (MU)-sequenced tobamovirus-normalized wastewater metagenomic sequencing abundance from Boston Deer Island Treatment Plant (DITP) North and South systems compared with weekly positive test counts from Massachusetts General Hospital (MGH) testing. (b) SecureBio (SB)-sequenced tobamovirus-normalized wastewater metagenomic sequencing abundance compared with the same clinical data. (c) Weekly testing volume with negative and positive results, along with positivity rate. Clinical testing followed a seasonal algorithm with expanded multiplex molecular panels during fall and winter months (see Methods). Spearman correlation coefficients (R) shown with significance: *$p < 0.05$, **$p < 0.01$, ***$p < 0.001$.

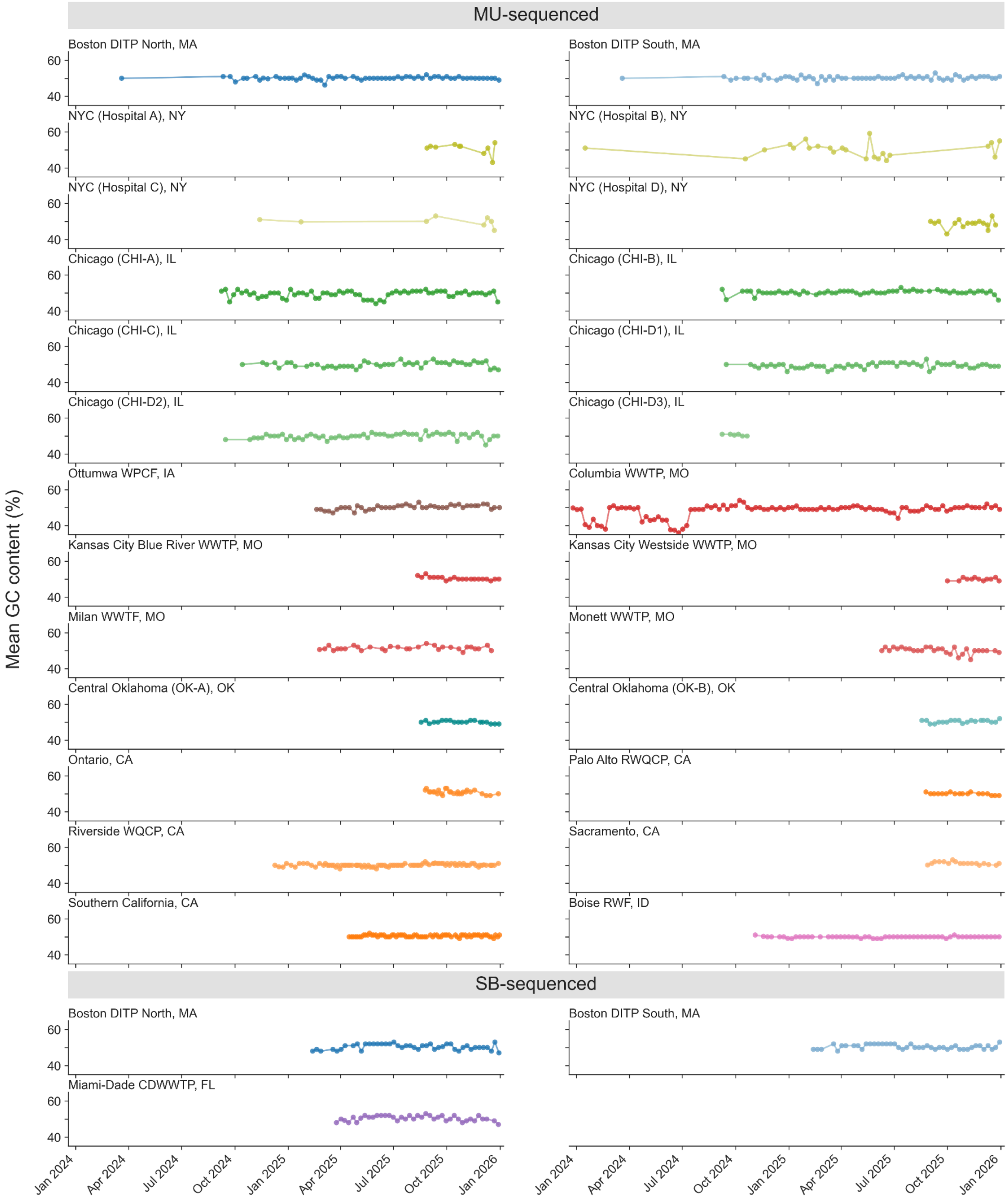


**Supplementary Figure S14. GC content over time by site.** Mean GC content (%) for each sampling location, grouped by sequencing partner. Values were calculated from subsampled reads after adapter trimming and quality filtering (see Methods). The transient decrease for Columbia, MO in mid-2024 coincides with the transition from NovaSeq 6000 to NovaSeq X.

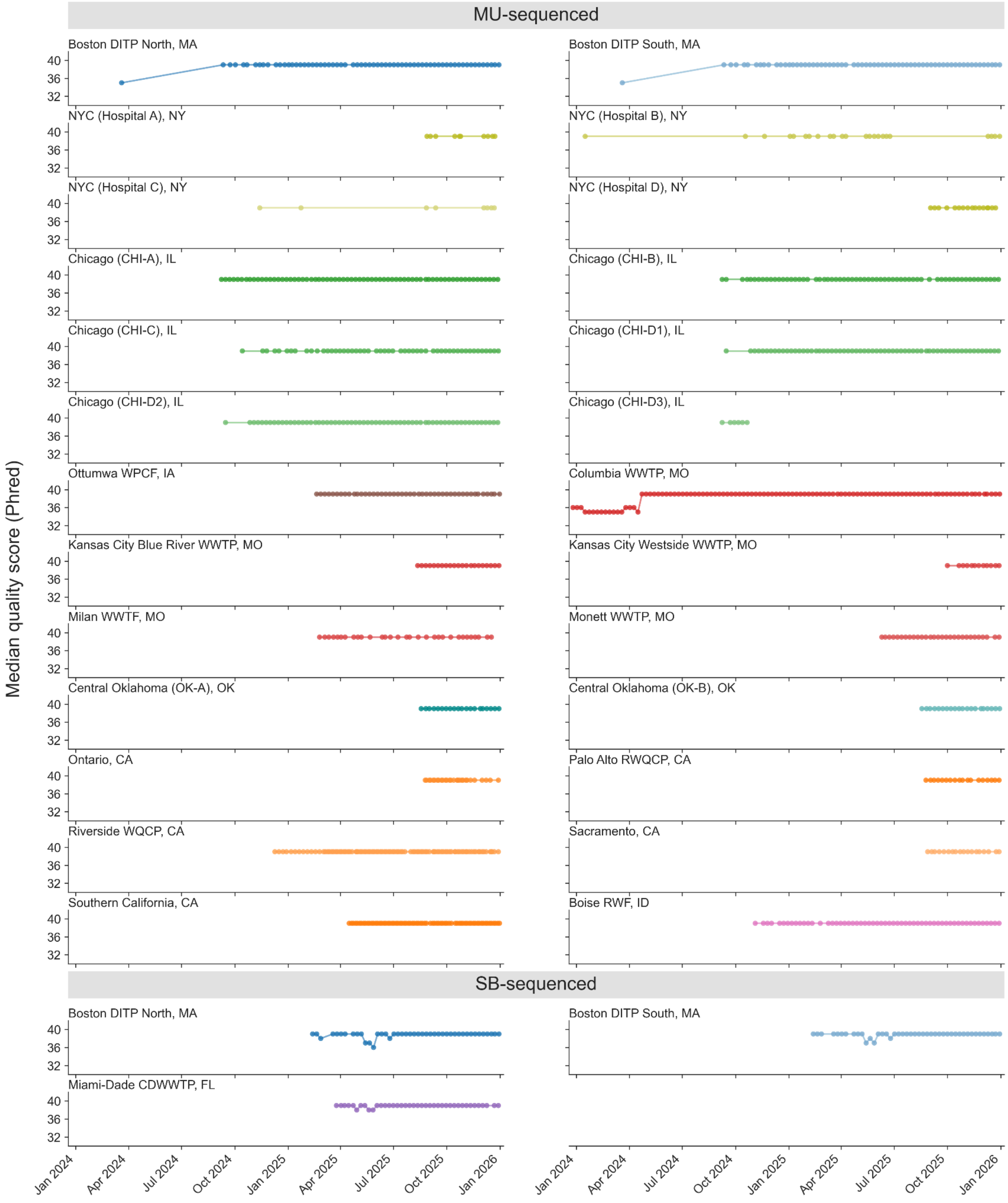


**Supplementary Figure S15. Sequencing quality scores over time by site.** Median raw read Phred quality score for each sampling location, grouped by sequencing partner. The increase for Columbia, MO and Boston, MA in mid-2024 coincides with the transition from NovaSeq 6000 to NovaSeq X.

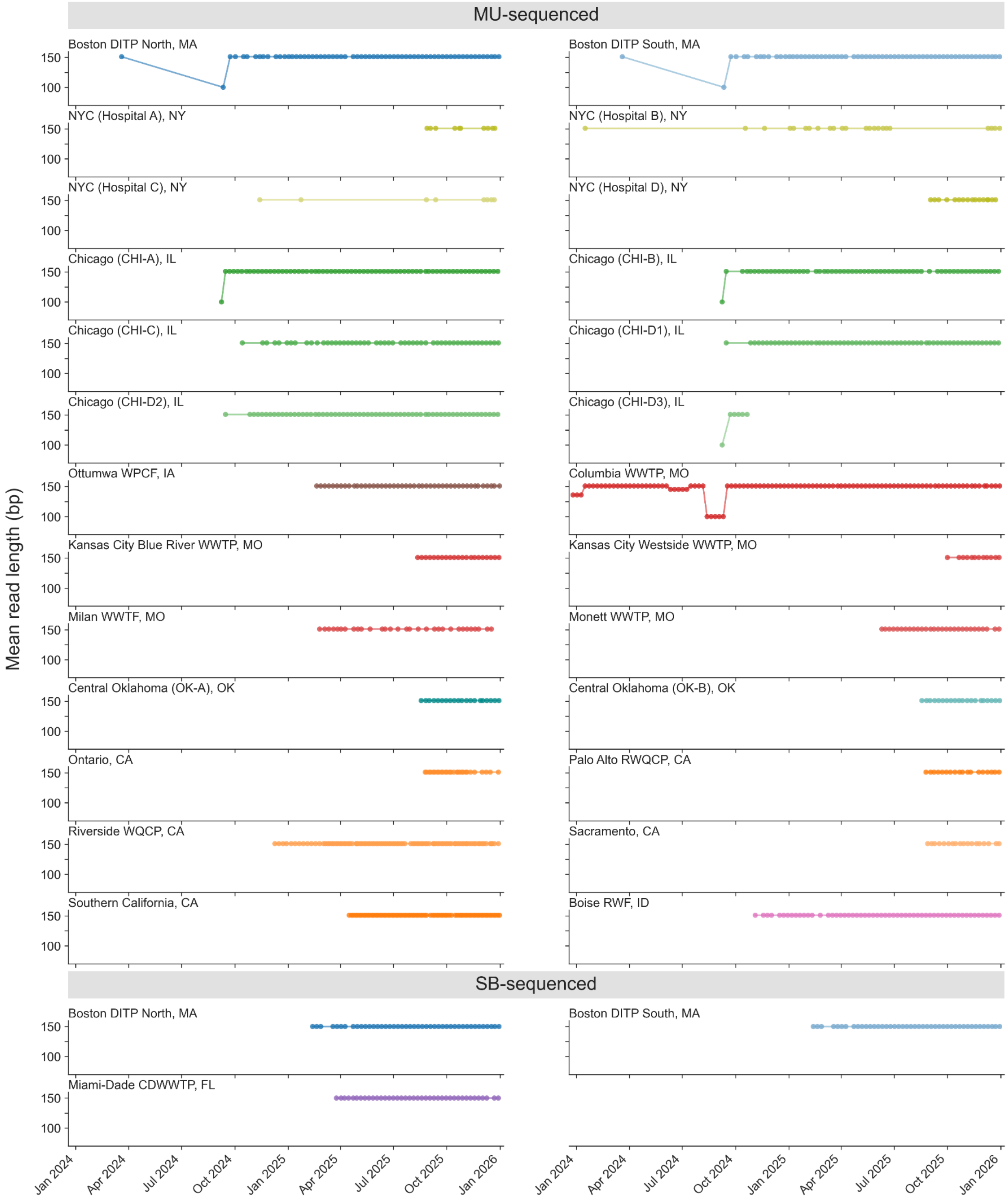


**Supplementary Figure S16. Read length over time by site.** Mean raw read length (bp) for each sampling location, grouped by sequencing laboratory. Most libraries were sequenced at 2x150 bp; 17 MU-processed libraries were sequenced at 2x100 bp due to diagnostic runs or sequencing configuration errors (see Methods).

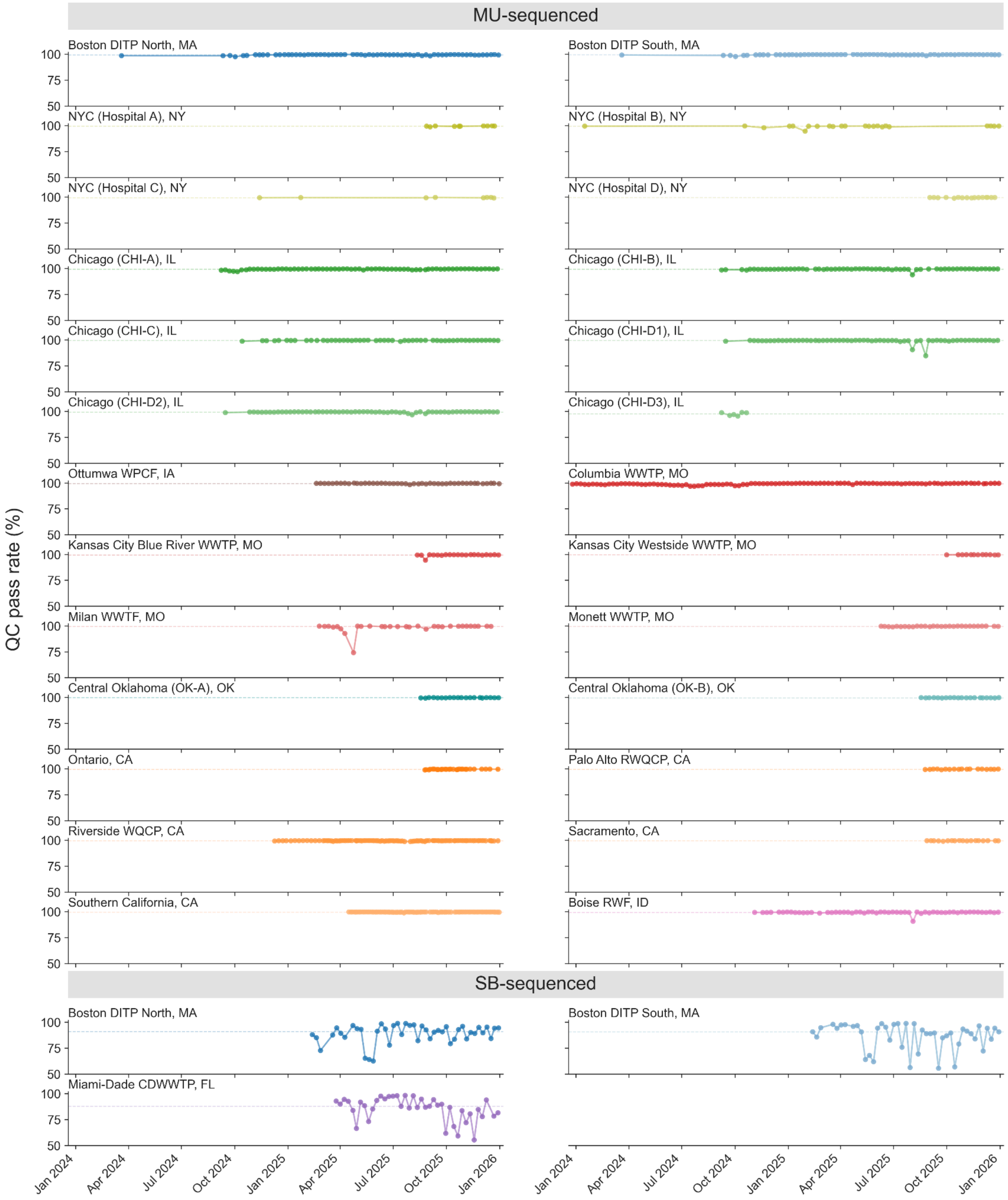


**Supplementary Figure S17. Quality control (QC) pass rate over time by site.** Fraction of reads passing adapter trimming and quality filtering by FASTP (see Methods).

**Supplementary Table S1. Summary of sequencing depth across published wastewater metagenomic sequencing (WW-MGS) studies.** Median and total gigabases (Gb) are reported for untargeted WW-MGS samples only, excluding amplicon or hybrid-capture samples where applicable. Base pair statistics were obtained either directly from the NCBI Sequence Read Archive (SRA) or estimated from values reported in the manuscript text, tables, or supplementary materials. Studies where per-sample sequencing depth could not be determined from public data or manuscript reporting are noted. CASPER data comprise two BioProjects: the Los Angeles pilot dataset (PRJNA1198001; Grimm et al., 2025) and the multi-site release described in this paper (PRJNA1247874; Justen et al., 2026).

| Study | Bioproject | Median Gb (WW-MGS samples) | Total Gb (WW-MGS samples) | Base stats method |
|---|---|---|---|---|
| (Cantalupo et al., 2011) | PRJNA70623 | 0.4 | 1.1 | SRA |
| (Ng et al., 2012) | PRJNA169010 | 0.2 | 0.8 | SRA |
| (Bibby & Peccia, 2013) | | 3.2 | 38.7 | estimated |
| (Hjelmsø et al., 2017) | PRJEB15242 | 0.9 | 40.9 | estimated |
| (Fernandez-Cassi et al., 2018) | | 0.3 | 1.2 | estimated |
| (Adriaenssens et al., 2018) | PRJNA421889 | 0.1 | 0.7 | SRA |
| (Maritz et al., 2019) | PRJEB28033 | 5.1 | 86.4 | SRA |
| (Martínez-Puchol et al., 2020) | – | Could not determine | Could not determine | – |
| (Gulino et al., 2020) | PRJEB28033 | 5.1 | 86.4 | SRA |
| (Nieuwenhuijse et al., 2020) | PRJEB23496 | 1.4 | 146.4 | SRA |
| (Guajardo-Leiva et al., 2020) | PRJNA648644 | 6.8 | 6.8 | SRA |
| (Brinch et al., 2020) | PRJEB34633 | 4.1 | 27.6 | SRA |
| (Wang et al., 2020) | – | 1.8 | 23.6 | estimated |
| (Crits-Christoph et al., 2021) | PRJNA661613 | 6.3 | 45.2 | SRA |
| (Rothman et al., 2021) | PRJNA729801 | 3.9 | 1189.4 | SRA |
| (Spurbeck et al., 2023) | PRJNA924011 | Could not determine | Could not determine | – |
| (Stockdale et al., 2023) | PRJNA842541 | 5.6 | 771.8 | SRA |
| (McCall et al., 2023) | PRJNA943189 | 10.8 | 21.6 | SRA |
| (Child et al., 2023) | PRJEB62830 | Could not determine | Could not determine | – |
| (Fernandez-Cassi & Kohn, 2024) | PRJNA948647 | 0.8 | 12.4 | SRA |
| (Wyler et al., 2024) | PRJNA948850 | 2.5 | 307.5 | estimated |
| (Tierney et al., 2024) | PRJNA946141 | 6.0 | 8235.1 | SRA |
| (Grimm et al., 2025) (CASPER) | PRJNA1198001 | 327.3 | 13132.6 | SRA |
| (Guajardo-Leiva et al., 2025) | PRJNA1196808 | 10.1 | 136.3 | SRA |
| (Worp et al., 2025) | PRJEB87273 | 0.1 | 28.7 | estimated |
| (Bellekom et al., 2026) | PRJNA1261831 | Could not determine | Could not determine | – |
| Justen et al., 2026 (CASPER) | PRJNA1247874 | 289.9 | 357869 | SRA |

**Supplementary Table S2. Historical sequencing cost of a typical CASPER sample.** Cost per megabase (Mb) is derived from the NHGRI Genome Sequencing Costs dataset through February 2022, with subsequent values estimated based on observed sequencing output and pricing (Kaufman, 2025; Wetterstrand, 2019). Per-sample cost is calculated using the median CASPER untargeted sequencing depth in this release of 290 Gb per sample.

| Date | Cost per Mb | Cost per median CASPER sample (290 Gb) | Source |
|---|---|---|---|
| Jan 2007 | $522.71 | $151.43M | NHGRI |
| Jan 2008 | $102.13 | $29.59M | NHGRI |
| Jan 2009 | $2.59 | $750.32k | NHGRI |
| Jan 2010 | $0.52 | $150.64k | NHGRI |
| Jan 2011 | $0.23 | $66.63k | NHGRI |
| Jan 2012 | $0.09 | $26.07k | NHGRI |
| Jan 2013 | $0.06 | $17.38k | NHGRI |
| Jan 2014 | $0.04 | $11.59k | NHGRI |
| Jan 2015 | $0.04 | $11.59k | NHGRI |
| May 2016 | $0.013 | $3.77k | NHGRI |
| Feb 2017 | $0.011 | $3.19k | NHGRI |
| Feb 2018 | $0.014 | $4.06k | NHGRI |
| Feb 2019 | $0.011 | $3.19k | NHGRI |
| Feb 2020 | $0.007 | $2.03k | NHGRI |
| Feb 2021 | $0.009 | $2.61k | NHGRI |
| Feb 2022 | $0.006 | $1.74k | NHGRI |
| Jul 2023 | $0.005 | $1.45k | estimated |
| Jul 2024 | $0.003 | $869.10 | estimated |
| Jan 2025 | $0.002 | $579.40 | estimated |
| Jan 2026 | $0.002 | $579.40 | estimated |

**Supplementary Table S3. Correlation between wastewater metagenomic sequencing and wastewater PCR data.** Spearman correlation coefficients (R) comparing wastewater metagenomic sequencing (WW-MGS) abundance with wastewater PCR measurements from the CDC National Wastewater Surveillance System data portal, with p values in parentheses. Results are shown for WW-MGS abundance expressed as total read relative abundance and as tobamovirus-normalized abundance. PCR pathogen concentrations are reported without normalization, in each source's native units (Methods). Correlations were calculated using MMWR week-aggregated data with centered 5-week smoothing. MU, University of Missouri; SB, SecureBio.

| | | Spearman correlation coefficient (R) and significance (p) | | | | | |
|---|---|---|---|---|---|---|---|
| | | Total read relative abundance | | | Tovamovirus-normalized | | |
| **Sampling site** | **N (weeks)** | **SARS-CoV-2** | **Influenza A** | **RSV** | **SARS-CoV-2** | **Influenza A** | **RSV** |
| **MU-sequenced** | | | | | | | |
| Boston DITP North, MA | 50 | -0.22 (0.125) | 0.64 (5.9e-07) | 0.85 (7.6e-15) | 0.79 (7.8e-12) | 0.77 (4.3e-11) | 0.90 (8.1e-19) |
| Boston DITP South, MA | 50 | -0.20 (0.158) | 0.76 (1.1e-10) | 0.62 (1.7e-06) | 0.68 (7.4e-08) | 0.87 (1.5e-16) | 0.66 (1.6e-07) |
| Chicago (CHI-A), IL | 69 | 0.23 (0.061) | 0.67 (2.2e-10) | 0.85 (9.0e-21) | 0.80 (2.8e-16) | 0.68 (1.6e-10) | 0.85 (3.6e-20) |
| Chicago (CHI-B), IL | 63 | 0.11 (0.411) | 0.42 (6.9e-04) | 0.70 (1.5e-10) | 0.85 (1.6e-18) | 0.49 (4.9e-05) | 0.71 (9.7e-11) |
| Chicago (CHI-D1), IL | 63 | 0.11 (0.391) | 0.74 (6.4e-12) | 0.36 (0.004) | 0.82 (9.7e-17) | 0.70 (1.5e-10) | 0.42 (6.2e-04) |
| Chicago (CHI-D2), IL | 63 | -0.22 (0.084) | 0.83 (5.5e-17) | 0.74 (4.2e-12) | 0.81 (1.1e-15) | 0.86 (2.0e-19) | 0.72 (3.9e-11) |
| Ottumwa WPCF, IA | 31 | 0.50 (0.005) | 0.32 (0.077) | 0.77 (3.3e-07) | 0.83 (5.3e-09) | 0.62 (2.2e-04) | 0.89 (2.4e-11) |
| Columbia WWTP, MO | 101 | 0.80 (8.9e-24) | 0.51 (4.0e-06) | 0.69 (1.7e-15) | 0.74 (6.1e-19) | 0.57 (1.7e-07) | 0.74 (2.0e-18) |
| Monett WWTP, MO | 24 | 0.83 (4.6e-07) | nan | 0.14 (0.522) | 0.55 (0.005) | nan | 0.20 (0.347) |
| Riverside WQCP, CA | 41 | 0.17 (0.294) | 0.85 (3.4e-12) | 0.93 (3.9e-18) | 0.83 (1.6e-11) | 0.89 (9.1e-15) | 0.88 (3.9e-14) |
| Boise RWF, ID | 42 | -0.02 (0.887) | 0.56 (1.1e-04) | 0.56 (9.8e-05) | 0.83 (1.5e-11) | 0.75 (9.7e-09) | 0.77 (3.3e-09) |
| **SB-sequenced** | | | | | | | |
| Boston DITP North, MA | 29 | 0.69 (3.7e-05) | 0.89 (5.9e-11) | 0.86 (3.1e-09) | 0.59 (7.9e-04) | 0.89 (5.9e-11) | 0.87 (1.1e-09) |
| Boston DITP South, MA | 29 | 0.30 (0.111) | 0.39 (0.034) | 0.81 (1.3e-07) | 0.25 (0.194) | 0.61 (4.2e-04) | 0.86 (2.5e-09) |
| Miami-Dade CDWWTP, FL | 26 | 0.60 (0.001) | nan | nan | 0.57 (0.003) | nan | nan |

**Supplementary Table S4. Correlation between wastewater metagenomic sequencing and clinical testing data.** Spearman correlation coefficients (R) comparing wastewater metagenomic sequencing (WW-MGS) abundance from Boston Deer Island Treatment Plant (DITP) with positive test counts from Massachusetts General Hospital (MGH) clinical diagnostic testing. P values are in parentheses. Clinical data represent aggregated weekly positive test counts from multiplex nucleic acid amplification tests. Results are shown for WW-MGS abundance expressed as total read relative abundance and as tobamovirus-normalized abundance. Correlations were calculated using MMWR week-aggregated data with centered 5-week smoothing. MU, University of Missouri. SB, SecureBio.

| | | Spearman correlation coefficient (R) and significance (p) | | | | | |
|---|---|---|---|---|---|---|---|
| | | Total read relative abundance | | | Tobamovirus-normalized | | |
| **Sampling site** | **N (weeks)** | **SARS-CoV-2** | **Influenza A** | **RSV** | **SARS-CoV-2** | **Influenza A** | **RSV** |
| **MU-sequenced** | | | | | | | |
| Boston DITP North, MA | 47 | 0.18 (0.228) | 0.51 (2.6e-04) | 0.69 (6.4e-08) | 0.52 (1.8e-04) | 0.68 (1.2e-07) | 0.84 (2.0e-13) |
| Boston DITP South, MA | 47 | 0.19 (0.196) | 0.63 (2.1e-06) | 0.65 (7.2e-07) | 0.62 (2.8e-06) | 0.76 (3.9e-10) | 0.74 (3.5e-09) |
| **SB-sequenced** | | | | | | | |
| Boston DITP North, MA | 26 | 0.33 (0.102) | 0.92 (3.2e-11) | 0.82 (3.0e-07) | 0.58 (0.002) | 0.92 (3.2e-11) | 0.84 (7.9e-08) |
| Boston DITP South, MA | 26 | 0.85 (4.1e-08) | 0.49 (0.011) | 0.73 (1.9e-05) | 0.08 (0.713) | 0.70 (7.0e-05) | 0.83 (1.7e-07) |

**Supplementary Table S5. Composition and classification accuracy of the benchmarking panels.** Paired-end reads were simulated from each panel and processed through the vertebrate-infecting virus (VV) identification workflow (Methods). The in-index panel comprises reference genomes drawn from the workflow's VV genome database (index version 20250825); the out-of-index panel comprises GenBank genomes of the same taxa absent from that database and deposited after its build date; the negative panel comprises the most abundant non-VV taxa in CASPER wastewater. Positive-panel read pairs are scored as correct at species level, correct at genus but not species level, or incorrect. For the negative panel, any read pair assigned to a vertebrate-infecting virus is a false positive.

| | In-index positive set | Out-of-index positive set | Negative set |
|---|---|---|---|
| **Composition** | | | |
| Taxa | 93 | 76 | 189 |
| Genomes | 93 | 76 | 213 |
| Read pairs simulated | 930,000 | 760,000 | 94,500,000 |
| **Positive set performance** | | | |
| Correct at species (%) | 92.0 | 86.3 | – |
| Correct at genus (%) | 97.2 | 91.4 | – |
| Incorrect | 2.8 | 8.6 | – |
| **Negative set performance** | | | |
| Incorrectly assigned as VV (%) | – | – | $2.3 \times 10^{-5}$ |

**Supplementary Table S6. Classification accuracy of the vertebrate-infecting virus (VV) workflow by viral family.** Paired-end reads were simulated from each panel's reference genomes and processed through the VV identification workflow (Methods). The in-index panel comprises genomes drawn from the workflow's VV genome database (index version 20250825); the out-of-index panel comprises GenBank genomes absent from that database and deposited after its build date. Percentages are of simulated read pairs, averaged across the species within each family. "Correct at genus" is cumulative and includes reads correct at species; "incorrect" denotes reads assigned outside the correct genus or not captured by the VV pipeline.

| | In-index positive set | | | | Out-of-index positive set | | | |
|---|---|---|---|---|---|---|---|---|
| **Family** | Taxa | Correct at species (%) | Correct at genus (%) | Incorrect (%) | Taxa | Correct at species (%) | Correct at genus (%) | Incorrect (%) |
| Adenoviridae | 5 | 96.83 | 97.31 | 2.69 | 5 | 95.24 | 97.56 | 2.44 |
| Arenaviridae | 6 | 92.92 | 93.9 | 6.1 | 3 | 65.95 | 67.53 | 32.47 |
| Astroviridae | 2 | 97.56 | 98.06 | 1.94 | 2 | 98.44 | 99.04 | 0.96 |
| Caliciviridae | 2 | 98.35 | 98.35 | 1.65 | 2 | 96.76 | 96.76 | 3.24 |
| Coronaviridae | 6 | 87.76 | 98.68 | 1.32 | 6 | 84.86 | 97.72 | 2.28 |
| Filoviridae | 3 | 98.66 | 98.66 | 1.34 | 2 | 98.94 | 98.94 | 1.06 |
| Flaviviridae | 10 | 97.33 | 97.37 | 2.63 | 8 | 78.6 | 80.67 | 19.33 |
| Hantaviridae | 3 | 87.64 | 98.14 | 1.86 | 2 | 83.29 | 96.07 | 3.93 |
| Hepadnaviridae | 1 | 97.14 | 98.38 | 1.62 | 1 | 99.04 | 99.34 | 0.66 |
| Kolmioviridae | 1 | 56.52 | 95.88 | 4.12 | 1 | 66.62 | 99.51 | 0.49 |
| Matonaviridae | 1 | 99.16 | 99.17 | 0.83 | 1 | 99.22 | 99.24 | 0.76 |
| Nairoviridae | 1 | 97.59 | 97.76 | 2.24 | 1 | 65.95 | 66.02 | 33.98 |
| Orthoherpesviridae | 9 | 76.22 | 95.51 | 4.49 | 6 | 79.04 | 95.01 | 4.99 |
| Orthomyxoviridae | 3 | 90.31 | 90.31 | 9.69 | 2 | 97.18 | 97.18 | 2.82 |
| Papillomaviridae | 1 | 93.66 | 93.66 | 6.34 | 1 | 99.03 | 99.04 | 0.96 |
| Paramyxoviridae | 8 | 98.14 | 98.15 | 1.85 | 7 | 98.03 | 98.03 | 1.97 |
| Parvoviridae | 2 | 88.15 | 98.17 | 1.83 | 2 | 86.78 | 98.16 | 1.84 |
| Peribunyaviridae | 3 | 86.91 | 98.82 | 1.18 | 2 | 80.55 | 93.81 | 6.19 |
| Phenuiviridae | 1 | 97.52 | 97.97 | 2.03 | 1 | 97.85 | 97.85 | 2.15 |
| Picornaviridae | 9 | 98.09 | 98.2 | 1.8 | 9 | 84.29 | 85.39 | 14.61 |
| Pneumoviridae | 2 | 98.01 | 98.01 | 1.99 | 2 | 97.33 | 97.33 | 2.67 |
| Polyomaviridae | 1 | 42.63 | 99.49 | 0.51 | 1 | 42.87 | 99.08 | 0.92 |
| Poxviridae | 3 | 85.8 | 96.49 | 3.51 | 1 | 91.49 | 97.62 | 2.38 |
| Retroviridae | 2 | 98.02 | 98.06 | 1.94 | 2 | 98.0 | 98.02 | 1.98 |
| Rhabdoviridae | 1 | 98.11 | 98.12 | 1.88 | 1 | 98.05 | 98.08 | 1.92 |
| Sedoreoviridae | 3 | 98.46 | 98.46 | 1.54 | 3 | 66.26 | 66.26 | 33.74 |
| Togaviridae | 3 | 97.12 | 97.15 | 2.85 | 2 | 97.64 | 97.66 | 2.34 |

**Supplementary Table S7. Agreement between the University of Missouri (MU) and SecureBio (SB) protocols on paired samples from Boston Deer Island Treatment Plant (DITP).** DITP North and South samples were split between the two laboratories, giving 44 collection dates per system (2025-02-12 to 2025-12-30) on which both protocols processed the same wastewater. Correlation rows report Spearman coefficients (R) with p values in parentheses, calculated on MMWR-week series smoothed with a centered five-week moving average, as plotted in Supplementary Figure S9. Between-protocol correlations are reported for both metrics: total read relative abundance (clade-assigned read pairs per total read pairs) and tobamovirus-normalized abundance. Ratio rows report the median MU/SB abundance ratio across matched collection dates on which both protocols detected the target (≥ 1 assigned read pair), with the number of such dates given as n. N is the number of overlapping MMWR weeks, restricted throughout to weeks covered by both laboratories. Wastewater PCR is reported to the National Wastewater Surveillance System by WastewaterSCAN in copies per gram of dry sludge; clinical data are weekly positive test counts from Massachusetts General Hospital.

| | | Spearman correlation coefficient (R) and significance (p) | | | | | |
|---|---|---|---|---|---|---|---|
| | | Boston DITP South | | | Boston DITP North | | |
| **Comparison** | **N** | **SARS-CoV-2** | **Influenza A** | **RSV** | **SARS-CoV-2** | **Influenza A** | **RSV** |
| **Between protocols** | | | | | | | |
| Correlation (total read relative abundance) | 44 | 0.29 (0.055) | 0.31 (0.040) | −0.00 (0.997) | −0.21 (0.176) | 0.62 (5.9e-06) | 0.78 (5.8e-10) |
| Correlation (tobamovirus normalized) | 44 | 0.56 (8.2e-05) | 0.53 (2.5e-04) | 0.22 (0.143) | 0.82 (8.0e-12) | 0.70 (9.3e-08) | 0.80 (6.1e-11) |
| Median MU/SB ratio (total read relative abundance) | – | 3.0 (n = 37) | 0.7 (n = 8) | 36.6 (n = 1) | 7.5 (n = 37) | 5.3 (n = 12) | 0.7 (n = 9) |
| Median MU/SB ratio (tobamovirus normalized) | – | 4.8 (n = 37) | 9.6 (n = 8) | 87.2 (n = 1) | 3.6 (n = 37) | 6.3 (n = 12) | 5.2 (n = 9) |
| **vs wastewater PCR** | | | | | | | |
| MU correlation | 29 | 0.45 (0.014) | 0.82 (7.2e-08) | 0.39 (0.037) | 0.68 (4.3e-05) | 0.65 (1.2e-04) | 0.76 (1.8e-06) |
| SB correlation | 29 | 0.25 (0.194) | 0.61 (4.2e-04) | 0.86 (2.5e-09) | 0.59 (7.9e-04) | 0.89 (5.9e-11) | 0.87 (1.1e-09) |
| **vs clinical testing** | | | | | | | |
| MU correlation | 26 | 0.75 (8.8e-06) | 0.61 (9.2e-04) | 0.38 (0.054) | 0.49 (0.010) | 0.48 (0.013) | 0.67 (1.7e-04) |
| SB correlation | 26 | 0.08 (0.713) | 0.70 (7.0e-05) | 0.83 (1.7e-07) | 0.58 (0.002) | 0.92 (3.2e-11) | 0.84 (7.9e-08) |

**Supplementary Table S8. Read pairs assigned to the pathogens shown in Figure 4c.** Assignments were made using the vertebrate-infecting virus (VV) pipeline (Methods) and are not adjusted for sequencing depth. Detections are samples with at least one assigned read pair; non-detects are samples with none. Distributions are reported over all 1,206 samples in this release and over the subset with a detection. IQR, interquartile range.

| Pathogen | Non-detects | All samples | | | Samples with a detection | |
|---|---|---|---|---|---|---|
| | | Median | IQR | Range | Median | IQR |
| SARS-CoV-2 | 120 / 1,206 (10.0%) | 67 | 15-176 | 0-5,727 | 82 | 27-192 |
| Influenza A | 704 / 1,206 (58.4%) | 0 | 0-7 | 0-172,176 | 9 | 3-31 |
| Influenza B | 991 / 1,206 (82.2%) | 0 | 0-0 | 0-453 | 4 | 2-10 |
| RSV | 872 / 1,206 (72.3%) | 0 | 0-1 | 0-171 | 6 | 3-14 |
| HCoV-HKU1 | 822 / 1,206 (68.2%) | 0 | 0-2 | 0-265 | 5 | 2-13 |
| HCoV-NL63 | 811 / 1,206 (67.2%) | 0 | 0-2 | 0-71 | 5 | 2-11 |
| HCoV-229E | 503 / 1,206 (41.7%) | 2 | 0-14 | 0-566 | 11 | 5-26 |
| HCoV-OC43 | 1,099 / 1,206 (91.1%) | 0 | 0-0 | 0-28 | 2 | 1-4 |
| Parainfluenza 2 | 761 / 1,206 (63.1%) | 0 | 0-6 | 0-141 | 10 | 4-26 |
| Parainfluenza 3 | 657 / 1,206 (54.5%) | 0 | 0-8 | 0-821 | 9 | 4-37 |
| Metapneumovirus | 1,031 / 1,206 (85.5%) | 0 | 0-0 | 0-30 | 4 | 2-7 |
| Enterovirus D68 | 1,052 / 1,206 (87.2%) | 0 | 0-0 | 0-26 | 3 | 1-6 |
| Norovirus | 7 / 1,206 (0.6%) | 5,596 | 1,572-15,621 | 0-551,492 | 5,707 | 1,610-15,687 |
| Rotavirus | 21 / 1,206 (1.7%) | 16,298 | 3,508-63,608 | 0-14,975,409 | 17,246 | 3,812-64,863 |
| Astrovirus | 21 / 1,206 (1.7%) | 28,140 | 6,928-93,339 | 0-2,629,454 | 29,170 | 7,832-93,994 |
| Sapovirus | 32 / 1,206 (2.7%) | 1,620 | 425-5,035 | 0-91,510 | 1,728 | 482-5,140 |

**Supplementary Table S9. National Wastewater Surveillance System (NWSS) sewershed identifiers for CASPER sites.** Identifiers enable linkage to publicly available NWSS PCR data. Sites marked "n/a" lack site-specific NWSS data; sites marked "–" either have no NWSS match or declined to share this information.

| Sewershed | NWSS sewershed_id |
|---|---|
| Boston DITP North, MA | 752, 742 |
| Boston DITP South, MA | 743, 742 |
| New York (Hospital A), NY | n/a |
| New York (Hospital B), NY | n/a |
| New York (Hospital C), NY | n/a |
| New York (Hospital D), NY | n/a |
| Chicago (CHI-A), IL | 419 |
| Chicago (CHI-B), IL | 413 |
| Chicago (CHI-C), IL | n/a |
| Chicago (CHI-D1), IL | 423 |
| Chicago (CHI-D2), IL | 429 |
| Chicago (CHI-D3), IL | n/a |
| Ottumwa WPCF, IA | 541 |
| Columbia WWTP, MO | 1045 |
| Kansas City Blue River WWTP, MO | 1074 |
| Kansas City Westside WWTP, MO | 1075 |
| Milan WWTF, MO | n/a |
| Monett WWTP, MO | 1041 |
| Miami-Dade CDWWTP, FL | 309 |
| Central Oklahoma (OK-A), OK | – |
| Central Oklahoma (OK-B), OK | – |
| Ontario, CA | 143 |
| Palo Alto RWQCP, CA | 175 |
| Riverside WQCP, CA | 138 |
| Sacramento, CA | 140 |
| Southern California, CA | – |
| Boise RWF, ID | 372, 374 |

**Supplementary Table S10. Wastewater PCR assay details and data source designations for CASPER comparison sites.** Data were obtained from the CDC National Wastewater Surveillance System (NWSS) data portal; source designations indicate whether data were generated by state and local health departments or by WastewaterSCAN (Boehm et al. 2026).

| Sampling site | Source | PCR type | PCR targets |
|---|---|---|---|
| Boston DITP North, MA (742) | | | |
| SARS-CoV-2 | WastewaterSCAN | ddPCR | N |
| Influenza A | WastewaterSCAN | ddPCR | INFA1, INFA1+INFA2 |
| RSV | WastewaterSCAN | ddPCR | RSV-A+RSV-B |
| Boston DITP South, MA (742) | | | |
| SARS-CoV-2 | WastewaterSCAN | ddPCR | N |
| Influenza A | WastewaterSCAN | ddPCR | INFA1, INFA1+INFA2 |
| RSV | WastewaterSCAN | ddPCR | RSV-A+RSV-B |
| Chicago (CHI-A), IL (419) | | | |
| SARS-CoV-2 | State/local health dept. | dPCR | N1 |
| Influenza A | State/local health dept. | dPCR | INFA1+INFA2 |
| RSV | State/local health dept. | dPCR | RSV-A+RSV-B |
| Chicago (CHI-B), IL (413) | | | |
| SARS-CoV-2 | State/local health dept. | dPCR | N1 |
| Influenza A | State/local health dept. | dPCR | INFA1+INFA2 |
| RSV | State/local health dept. | dPCR | RSV-A+RSV-B |
| Chicago (CHI-D1), IL (423) | | | |
| SARS-CoV-2 | State/local health dept. | dPCR | N1 |
| Influenza A | State/local health dept. | dPCR | INFA1+INFA2 |
| RSV | State/local health dept. | dPCR | RSV-A+RSV-B |
| Chicago (CHI-D2), IL (429) | | | |
| SARS-CoV-2 | State/local health dept. | dPCR | N1 |
| Influenza A | State/local health dept. | dPCR | INFA1+INFA2 |
| RSV | State/local health dept. | dPCR | RSV-A+RSV-B |
| Ottumwa WPCF, IA (541) | | | |
| SARS-CoV-2 | WastewaterSCAN | ddPCR | N |
| Influenza A | WastewaterSCAN | ddPCR | INFA1+INFA2 |
| RSV | WastewaterSCAN | ddPCR | RSV-A+RSV-B |
| Columbia WWTP, MO (1045) | | | |
| SARS-CoV-2 | State/local health dept. | dPCR | N2+N1 |
| Influenza A | State/local health dept. | dPCR | M |
| RSV | State/local health dept. | dPCR | M+N2 |
| Kansas City Westside WWTP, MO (1075) | | | |
| SARS-CoV-2 | State/local health dept. | dPCR | N2+N1 |
| Influenza A | State/local health dept. | dPCR | M |
| RSV | State/local health dept. | dPCR | M+N2 |
| Monett WWTP, MO (1041) | | | |
| SARS-CoV-2 | State/local health dept. | dPCR | N2+N1 |
| Influenza A | State/local health dept. | dPCR | M |
| RSV | State/local health dept. | dPCR | M+N2 |
| Miami-Dade CDWWTP, FL (309) | | | |
| SARS-CoV-2 | WastewaterSCAN | ddPCR | N |
| Influenza A | WastewaterSCAN | ddPCR | INFA1, INFA1+INFA2 |
| RSV | WastewaterSCAN | ddPCR | RSV-A+RSV-B |
| Riverside WQCP, CA (138) | | | |
| SARS-CoV-2 | WastewaterSCAN | ddPCR | N |
| Influenza A | WastewaterSCAN | ddPCR | INFA1+INFA2 |
| RSV | WastewaterSCAN | ddPCR | RSV-A+RSV-B |
| Boise RWF, ID (372) | | | |
| SARS-CoV-2 | WastewaterSCAN | ddPCR | N |
| Influenza A | WastewaterSCAN | ddPCR | INFA1, INFA1+INFA2 |
| RSV | WastewaterSCAN | ddPCR | RSV-A+RSV-B |

**Supplementary Table S11. SecureBio (SB) samples prepared with ribodepletion reagent omission.** RiboPOOLs probes were omitted from SB library preparation between 13 May and 1 July 2025. Samples affected are listed with their BioSample accession; ribodepletion status for all samples in this release is recorded in the analysis repository (Code availability). The University of Missouri protocol does not include a ribodepletion step.

| Site | Collection date | BioSample accession |
|---|---|---|
| Boston DITP North, MA | 2025-05-14 | SAMN55951608 |
| Boston DITP North, MA | 2025-05-21 | SAMN55951610 |
| Boston DITP North, MA | 2025-05-28 | SAMN55951612 |
| Boston DITP North, MA | 2025-06-04 | SAMN55951614 |
| Boston DITP North, MA | 2025-06-11 | SAMN55951616 |
| Boston DITP North, MA | 2025-06-18 | SAMN55951618 |
| Boston DITP North, MA | 2025-06-25 | SAMN55951620 |
| Boston DITP South, MA | 2025-05-14 | SAMN55951609 |
| Boston DITP South, MA | 2025-05-21 | SAMN55951611 |
| Boston DITP South, MA | 2025-05-28 | SAMN55951613 |
| Boston DITP South, MA | 2025-06-04 | SAMN55951615 |
| Boston DITP South, MA | 2025-06-11 | SAMN55951617 |
| Boston DITP South, MA | 2025-06-18 | SAMN55951619 |
| Boston DITP South, MA | 2025-06-25 | SAMN55951621 |
| Miami-Dade CDWWTP, FL | 2025-05-13 | SAMN54911859 |
| Miami-Dade CDWWTP, FL | 2025-05-20 | SAMN54911860 |
| Miami-Dade CDWWTP, FL | 2025-05-27 | SAMN54911861 |
| Miami-Dade CDWWTP, FL | 2025-06-03 | SAMN54911862 |
| Miami-Dade CDWWTP, FL | 2025-06-10 | SAMN54911863 |
| Miami-Dade CDWWTP, FL | 2025-06-17 | SAMN54911864 |
| Miami-Dade CDWWTP, FL | 2025-06-24 | SAMN54911865 |
| Miami-Dade CDWWTP, FL | 2025-07-01 | SAMN54911866 |

### References


Adriaenssens, E. M., Farkas, K., Harrison, C., Jones, D. L., Allison, H. E., & McCarthy, A. J. (2018). Viromic analysis of wastewater input to a river catchment reveals a diverse assemblage of RNA viruses. *mSystems*, *3*(3). https://doi.org/10.1128/mSystems.00025-18

Bellekom, B., Troman, C., Fitz, S., Akello, J. O., Grassly, N. C., & Shaw, A. G. (2026). Comparison of the sensitivity of targeted and untargeted (metagenomic) methods for the detection of viral pathogens in wastewater. *The Science of the Total Environment*, *1013*(181333), 181333.

Bibby, K., & Peccia, J. (2013). Identification of viral pathogen diversity in sewage sludge by metagenome analysis. *Environmental Science & Technology*, *47*(4), 1945–1951.

Boehm, A. B., Wolfe, M. K., Bidwell, A. L., Zulli, A., Chan-Herur, V., White, B. J., Shelden, B., & Duong, D. (2024). Human pathogen nucleic acids in wastewater solids from 191 wastewater treatment plants in the United States. *Scientific Data*, *11*(1), 1141.

Boehm, A. B., Wolfe, M. K., Bidwell, A. L., Zulli, A., White, B. J., Shelden, B., & Duong, D. (2026). Pathogen nucleic acids data in wastewater solids from 147 treatment plants in the United States: 2024-2025. *Data in Brief*, *65*(112503), 112503.

Brinch, C., Leekitcharoenphon, P., Duarte, A. S. R., Svendsen, C. A., Jensen, J. D., & Aarestrup, F. M. (2020). Long-term temporal stability of the resistome in sewage from Copenhagen. *mSystems*, *5*(5). https://doi.org/10.1128/mSystems.00841-20

Cantalupo, P. G., Calgua, B., Zhao, G., Hundesa, A., Wier, A. D., Katz, J. P., Grabe, M., Hendrix, R. W., Girones, R., Wang, D., & Pipas, J. M. (2011). Raw sewage harbors diverse viral populations. *mBio*, *2*(5). https://doi.org/10.1128/mBio.00180-11

Child, H. T., Airey, G., Maloney, D. M., Parker, A., Wild, J., McGinley, S., Evens, N., Porter, J., Templeton, K., Paterson, S., van Aerle, R., Wade, M. J., Jeffries, A. R., & Bassano, I. (2023). Comparison of metagenomic and targeted methods for sequencing human pathogenic viruses from wastewater. *mBio*, *14*(6), e0146823.

Crits-Christoph, A., Kantor, R. S., Olm, M. R., Whitney, O. N., Al-Shayeb, B., Lou, Y. C., Flamholz, A., Kennedy, L. C., Greenwald, H., Hinkle, A., Hetzel, J., Spitzer, S., Koble, J., Tan, A., Hyde, F., Schroth, G., Kuersten, S., Banfield, J. F., & Nelson, K. L. (2021). Genome sequencing of sewage detects regionally prevalent SARS-CoV-2 variants. *mBio*, *12*(1). https://doi.org/10.1128/mBio.02703-20

Fernandez-Cassi, X., & Kohn, T. (2024). Comparison of three viral nucleic acid preamplification pipelines for sewage viral metagenomics. *Food and Environmental Virology*, *16*(3), 1–22.

Fernandez-Cassi, X., Timoneda, N., Martínez-Puchol, S., Rusiñol, M., Rodriguez-Manzano, J., Figuerola, N., Bofill-Mas, S., Abril, J. F., & Girones, R. (2018). Metagenomics for the study of viruses in urban sewage as a tool for public health surveillance. *The Science of the Total Environment*, *618*, 870–880.

Grimm, S. L., Rothman, J. A., Bradshaw, W. J., Langlois, K., Steele, J. A., Griffith, J. F., Kaufman, J. T., & Whiteson, K. L. (2025). Deep metatranscriptomic sequencing data of wastewater from Los Angeles, USA, 2023-2024. *Scientific Data*, *13*(1), 158.

Guajardo-Leiva, S., Chnaiderman, J., Gaggero, A., & Díez, B. (2020). Metagenomic insights into the sewage RNA virosphere of a large city. *Viruses*, *12*(9), 1050.

Guajardo-Leiva, S., Díez, B., Rojas-Fuentes, C., Chnaiderman, J., Castro-Nallar, E., Catril, V., Ampuero, M., & Gaggero, A. (2025). From sewage to genomes: Expanding our understanding of the urban and semi-urban wastewater RNA virome. *Environmental Research*, *276*(121509), 121509.

Gulino, K., Rahman, J., Badri, M., Morton, J., Bonneau, R., & Ghedin, E. (2020). Initial mapping of the New York City wastewater virome. *mSystems*, *5*(3). https://doi.org/10.1128/mSystems.00876-19

Hjelmsø, M. H., Hellmér, M., Fernandez-Cassi, X., Timoneda, N., Lukjancenko, O., Seidel, M., Elsässer, D., Aarestrup, F. M., Löfström, C., Bofill-Mas, S., Abril, J. F., Girones, R., & Schultz, A. C. (2017). Evaluation of methods for the concentration and extraction of viruses from sewage in the context of metagenomic sequencing. *PloS One*, *12*(1), e0170199.

Kaufman, J. (2025, June 25). *How Much Data From a Sequencing Run?* SecureBio. https://data.securebio.org/jefftk-notebook/how-much-data-from-a-sequencing-run

Maritz, J. M., Ten Eyck, T. A., Elizabeth Alter, S., & Carlton, J. M. (2019). Patterns of protist diversity associated with raw sewage in New York City. *The ISME Journal*, *13*(11), 2750–2763.

Martínez-Puchol, S., Rusiñol, M., Fernández-Cassi, X., Timoneda, N., Itarte, M., Andrés, C., Antón, A., Abril, J. F., Girones, R., & Bofill-Mas, S. (2020). Characterisation of the sewage virome: comparison of NGS tools and occurrence of significant pathogens. *The Science of the Total Environment*, *713*(136604), 136604.

McCall, C., Leo Elworth, R. A., Wylie, K. M., Wylie, T. N., Dyson, K., Doughty, R., Treangen, T. J., Hopkins, L., Ensor, K., & Stadler, L. B. (2023). Targeted metagenomic sequencing for detection of vertebrate viruses in wastewater for public health surveillance. *ACS ES&T Water*. https://doi.org/10.1021/acsestwater.3c00183

Ng, T. F. F., Marine, R., Wang, C., Simmonds, P., Kapusinszky, B., Bodhidatta, L., Oderinde, B. S., Wommack, K. E., & Delwart, E. (2012). High variety of known and new RNA and DNA viruses of diverse origins in untreated sewage. *Journal of Virology*, *86*(22), 12161–12175.

Nieuwenhuijse, D. F., Oude Munnink, B. B., Phan, M. V. T., Global Sewage Surveillance project consortium, Munk, P., Venkatakrishnan, S., Aarestrup, F. M., Cotten, M., & Koopmans, M. P. G. (2020). Setting a baseline for global urban virome surveillance in sewage. *Scientific Reports*, *10*(1), 13748.

Rothman, J. A., Loveless, T. B., Kapcia, J., 3rd, Adams, E. D., Steele, J. A., Zimmer-Faust, A. G., Langlois, K., Wanless, D., Griffith, M., Mao, L., Chokry, J., Griffith, J. F., & Whiteson, K. L. (2021). RNA viromics of Southern California wastewater and detection of SARS-CoV-2 single-nucleotide variants. *Applied and Environmental Microbiology*, *87*(23), e0144821.

Spurbeck, R. R., Catlin, L. A., Mukherjee, C., Smith, A. K., & Minard-Smith, A. (2023). Analysis of metatranscriptomic methods to enable wastewater-based biosurveillance of all infectious diseases. *Frontiers in Public Health*, *11*. https://doi.org/10.3389/fpubh.2023.114527510.3389/fpubh.2023.1145275.s001

Stockdale, S. R., Blanchard, A. M., Nayak, A., Husain, A., Nashine, R., Dudani, H., McClure, C. P., Tarr, A. W., Nag, A., Meena, E., Sinha, V., Shrivastava, S. K., Hill, C., Singer, A. C., Gomes, R. L., Acheampong, E., Chidambaram, S. B., Bhatnagar, T., Vetrivel, U., … Monaghan, T. M. (2023). RNA-Seq of untreated wastewater to assess COVID-19 and emerging and endemic viruses for public health surveillance. *The Lancet Regional Health. Southeast Asia*, *14*(100205), 100205.

Tierney, B. T., Foox, J., Ryon, K. A., Butler, D., Damle, N., Young, B. G., Mozsary, C., Babler, K. M., Yin, X., Carattini, Y., Andrews, D., Lucaci, A. G., Solle, N. S., Kumar, N., Shukla, B., Vidović, D., Currall, B., Williams, S. L., Schürer, S. C., … Mason, C. E. (2024). Towards geospatially-resolved public-health surveillance via wastewater sequencing. *Nature Communications*, *15*(1), 8386.

Wang, H., Neyvaldt, J., Enache, L., Sikora, P., Mattsson, A., Johansson, A., Lindh, M., Bergstedt, O., & Norder, H. (2020). Variations among viruses in influent water and effluent water at a wastewater plant over one year as assessed by quantitative PCR and metagenomics. *Applied and Environmental Microbiology*, *86*(24). https://doi.org/10.1128/AEM.02073-20

Wetterstrand, K. A. (2019, March 13). *DNA Sequencing Costs: Data from the NHGRI Genome Sequencing Program (GSP)*. Genome.gov; NHGRI. https://www.genome.gov/about-genomics/fact-sheets/DNA-Sequencing-Costs-Data

Worp, N., Nieuwenhuijse, D. F., Izquierdo-Lara, R. W., Schapendonk, C. M. E., Brinch, C., Jensen, E. E. B., Munk, P., Hendriksen, R. S., Global Sewage Surveillance Consortium, Aarestrup, F., Oude Munnink, B. B., Koopmans, M. P. G., & de Graaf, M. (2025). Unveiling the global urban virome through wastewater metagenomics. *Nature Communications*, *16*(1), 10707.

Wyler, E., Lauber, C., Manukyan, A., Deter, A., Quedenau, C., Teixeira Alves, L. G., Wylezich,

C., Borodina, T., Seitz, S., Altmüller, J., & Landthaler, M. (2024). Pathogen dynamics and discovery of novel viruses and enzymes by deep nucleic acid sequencing of wastewater. *Environment International*, *190*(108875), 108875.
